# Preoperative risk prediction tools that predict morbidity risk in adults undergoing surgery: An Evidence Review

**DOI:** 10.1101/2025.06.27.25330118

**Authors:** Alesha Wale, Toby Ayres, Salina Khatoon, Amy Fox-McNally, Claire Morgan, Helen Morgan, Hannah Shaw, Jacob Davies, Rhiannon Tudor Edwards, Claire Dunstan, Adrian Edwards, Alison Cooper, Ruth Lewis

## Abstract

Risk prediction tools play a critical role in preoperative care by estimating the likelihood of adverse outcomes, including mortality, morbidity, and postoperative complications. In low-risk surgical settings such as surgical hubs, accurate risk prediction is particularly valuable. The aim of this review was to identify and map the evidence for 14 validated pre-operative surgical risk prediction tools currently used in Wales within any elective, or non-emergency surgical setting, and to provide a more in-depth look at the findings for a selection of tools deemed to be the most applicable on a population level to the context of surgical hubs.

Included studies were published between 1999 and 2024. No evidence was found for two of the risk prediction tools however, a total of 118 studies were identified across 12 risk prediction tools. None of the evidence found was looking at the predictive ability of risk prediction tools for selecting patients suitable for surgical hubs. The tools were used across a range of surgical specialties and measured composite complications, individual complications, and healthcare utilisation and recovery measures. No risk prediction tool adequately predicted complications across all surgical specialties. Among the included studies, there was considerable heterogeneity in which surgical specialties the risk prediction tools were used for, how complications were defined, and which measures were used to determine a tool’s predictive ability. This makes direct comparisons very challenging.

Four tools were selected as being potentially the most impactful at a population level for a more in-depth look at the findings: ACS NSQIP, P-POSSUM, RCRI, ASA classification system. A total of 76 studies were identified across these 4 tools. Key findings for the four risk prediction tools of interest are described. Overall, no one tool was identified that adequately predicted complications across all surgical specialties. The predictive ability of the tools varied across different surgical specialties.

Further research using consistent methods is needed to better understand the predictive ability of risk prediction tools and allow a robust evaluation. Given no single risk prediction tool adequately predicted complications across all surgical specialties, it may be likely that some tools are better suited for specific surgery types or that a combination of risk prediction tools may be needed to adequately assess an individual’s level of risk.

**Funding statement:** The authors and their Institutions were funded for this work by the Health and Care Research Wales Evidence Centre, itself funded by Health and Care Research Wales on behalf of Welsh Government.

## 1. BACKGROUND

### 1.1 Who is this Evidence Review for?

This Evidence Review was conducted as part of the Health and Care Research Wales Evidence Centre Work Programme. The review question was suggested by Planned Care Wales.

### 1.2 Background and purpose of this Evidence Review

Risk prediction tools play a critical role in preoperative care by estimating the likelihood of adverse outcomes, including mortality, morbidity, and postoperative complications (The National Institute for Health and Care Excellence [NICE], 2020). These tools support clinicians in making informed decisions about a patient’s overall suitability for surgery and identifying the need for enhanced postoperative care. However, despite their widespread use, there is significant variation in how these tools are applied across different disciplines and surgical settings, with no standardised approach being adopted (Pradhan et al. 2022).

In low-risk surgical settings such as surgical hubs, which typically focus on high-volume, low-complexity procedures, accurate risk prediction is particularly valuable. It helps ensure that patients selected for these settings can safely benefit from treatment while maintaining the efficiency and safety standards required for such facilities. A wide range of risk prediction tools are used for selecting patients who can safely be treated at a surgical hub. However, while the tools have been found to be accurate and reliable, given that there is no standardised approach to selecting risk prediction tools, their selection may not be evidence based and it can be unclear which tools should be used. Some of the tools used are not designed for use in surgery and others may not be best-suited for the low-risk surgical settings.

To ensure a risk prediction tool will be effective it is essential to refer to studies that evaluate its performance using new datasets, and not the dataset that was used to develop the model (referred to as *external validation studies*) (Collins et al., 2014). Assessing the performance of a risk prediction tool in other datasets allows evaluation of the transferability of the tool across different cohorts to examine how well it performs (Collins et al., 2014). External validation studies evaluate both *discrimination* and *calibration* to determine a tool’s performance (how accurately it predicts a risk) (Collins et al., 2014). Discrimination measures how well the tool differentiates between patients who do and do not experience an event, quantified using the area under the receiver operating characteristic (ROC) curve (AUC) or c-statistic. Calibration assesses the agreement between the predicted and observed outcomes, typically presented graphically as observed risks versus predicted risks or in tabular format (Collins et al., 2014). Some studies also report the accuracy of the tool using the ‘*Brier Score’*. The Brier score is a simultaneous measure of calibration and discrimination (Alzahrani et al., 2020).

A preliminary review of studies reporting on the external validation of risk prediction tools for assessing the risk of post-operative morbidity or mortality in any surgical settings identified a large volume of studies and a very complex picture of the evidence base. As such, it was decided that an evidence review would be conducted which included an initial rapid evidence map (REM) to understand this complex evidence landscape, and an in-depth summary of the findings for a selection of the risk prediction tools. The REM provides a description of the intended purpose of each tool, the context within which they have been developed and used, and the amount of external validation studies available. The findings of this initial map were then used to select a manageable sub-set of relevant risk prediction tools for a more in-depth evaluation of their validity.

There is a need to identify which validated risk prediction tools are most suitable for use/application to the Welsh population, and which ones are best for selecting patients that can safely go to surgical hubs. A review of external validation studies of risk prediction tools could help decision-makers when selecting which tool(s) to use for assessing patients’ risk of adverse outcomes, and could help to optimise resource allocation or candidacy for surgery in low-risk settings such as surgical hubs. As surgical hubs are a relatively new concept, and can be limited to specific surgical specialties, little evidence exists around the use of risk prediction tools within these settings. Therefore, the aims of this review were:

- To identify and map the evidence for validated pre-operative surgical risk prediction tools currently used in Wales in any elective, or non-emergency surgical settings (See Section 2).
- To provide a more in-depth summary of the findings for a selection of the tools deemed to be the most applicable on a population level (See Section 3).

This will allow a clearer picture of which tools may be most suitable for use within low-risk surgical settings, such as surgical hubs. As surgical hubs are intended as low-risk settings, this review focuses on external validation studies of risk prediction models for assessing the risk of **preoperative morbidity and complications**.

### 1.3 NICE guidance on preoperative risk stratification tools

A recent review conducted by NICE (2020) sought to explore which of three risk prediction tools (P-POSSUM, SORT, or ACS NSQIP) could best identify the risk of mortality and morbidity in adults undergoing surgery (NICE 2020). While separate findings were reported for morbidity and mortality, some of the included studies reporting morbidity also considered mortality as a complication. As surgical hubs are low-risk settings, our review focusses on complications only and not mortality. However, our review compliments the NICE evidence review by focussing on complications that do not include mortality, thus making it more applicable to low-risk settings, and by updating the evidence base to include more recently published studies. The studies included in the NICE review were screened for inclusion in this review, and any studies that reported complications separately to mortality were included in our review.

Whilst we recognise mortality can occur as a post-operative complication, the focus of our review is on morbidity only given the likelihood of death in low-risk surgical settings should be significantly lower than other settings. However, in order to better reflect the entire evidence base, a summary of studies included in the NICE review that were identified during screening that included mortality and complications in one composite outcome will be provided to ensure the evidence base as a whole is represented (See Appendix 1).

## 2. Summary of the evidence base included in the Rapid Evidence Map

The eligibility criteria used to select relevant validation studies for inclusion in this review and a description of the methods used to conduct the overall review are described in Section 6. This section starts with a description of the risk prediction tools included in the evidence map. Table 1 provides details about the original purpose of the tools, their uses and the outputs they provide. This is followed by a detailed description of the evidence base identified across all risk prediction tools. Table 2 includes how many studies were identified for each tool, the overall sample sizes used across studies, any specific population group being used to validate the tool, the number of studies identified for each surgical specialty, and the number of studies reporting on the different outcomes.

**Table 1.** Summary of risk prediction tools.

| Tool name | Purpose* | Who is it completed by | Surgical disciplines | Output* |
| --- | --- | --- | --- | --- |
| ACS NSQIP | Estimates the chance of an unfavourable outcome (such as a complication or death) after surgery through a series of questions relating to patient severity and surgical information.<br>The ACS NSQIP surgical risk calculator is a decision-support tool based on reliable multi-institutional clinical data, which can be used to estimate the risks of most operations. | Clinician | Any procedure, in most surgical subspecialties, | Individual risk scores for up to 19 different outcomes within 30-days following surgery, including a predicted length of stay. |
| POSSUM | Estimates morbidity and mortality for general surgery patients through a series of questions around patient physiological score which looks at age, cardiac and respiratory function, GCS, blood work, ECG, and operative severity which interrogates the procedure being undertaken. | Clinician | Emergency and elective general surgical procedures | A single overall score for predicted morbidity and a single overall score for predicted mortality. |
| P-POSSUM | POSSUM-P is very similar to the POSSUM tool but uses a slightly different formula to calculate risk. | Clinician | Same as POSSUM | Same as POSSUM |
| RCRI | Estimates risk of cardiac complications after non-cardiac surgery in patients ≥45 years old (or 18-44 years old with significant cardiovascular disease). Six questions focus on surgical risk, history of heart and cerebrovascular disease, preoperative treatment with insulin and preoperative creatinine levels. | Clinician | Elective non-cardiac surgery or urgent/semi-urgent (non-emergent) non-cardiac surgery. | A single overall score for predicted postoperative cardiac complications |
| ARISCAT | Predicts risk of pulmonary complications after surgery, including respiratory failure. Seven questions focussed on oxygen saturation levels, respiratory infections, anaemia, and surgery information. | Clinician | Surgery performed under general, neuraxial, or regional anaesthesia | A single overall score for predicted postoperative pulmonary complications |
| CPET | Provides an objective assessment of exercise capacity preoperatively and identifies the causes of exercise limitation. | Clinician | Abdominal, colorectal, urological, hepatobiliary, liver, bariatric, vascular, thoracic, oesophageal gastric. | Scores for VO<sup>2</sup> peak and anaerobic threshold |
| DASI | Measures functional capacity and assesses aspects of quality of life by asking 12 questions. Noted not to be as accurate as objective measures such as exercise stress testing. | Patients self-administer | Not specified but used during preoperative assessments for surgery and before and during exercise programmes | A single overall score for functional capacity |
| ASA Classification system | Classifies health of patients prior to surgery. Should not be used alone to determine appropriateness of surgery, but as part of a comprehensive preoperative evaluation. Other evaluations to be considered prior to surgery may include functional capacity, classification of surgery type, and other chronic conditions. The tool asks two questions relating to overall health and if surgery is an emergency procedure. | Clinician | Not specified however it should not be used alone to predict risk of morbidity. | Provides an ASA classification (I-VI) where a higher grade suggests higher operative risk |
| PONV | Predicts risk of postoperative nausea and vomiting (PONV) through four questions looking at gender, smoking status, history of motion sickness or PONV, and use of postoperative opioids. | Clinician | Any surgery performed under general anaesthesia | Provides an overall score for the risk of experiencing PONV |
| SORT | Provides an estimate of the risk of death within 30 days of an operation through a series of questions looking at: the procedure; ASA classification; clinical urgency based on the NCEPOD classification of intervention (2004); and patient age. This tool also requires a clinical risk assessment to estimate 30-day mortality (ideally made by a senior clinician in the multi-disciplinary peri-operative care team). | Clinician | Surgery not specified, but for use in adults only | Provides an overall risk prediction score for mortality |
| NRS-2002 | Predicts risk of malnutrition in hospitalised patients through four questions relating to weight, diet and if an ICU patient. | Clinician | N/A not originally designed for surgery | Provides an overall score for malnutritional risk |
| CFS | Launched during the COVID-19 crisis, it measures frailty to predict survival in medium and long term (mortality and need for institutional care) through a 9-point scale. . | Clinician | N/A not originally designed for surgery | Provides an overall frailty score |

\*See Section 2.2 for more details on what the tools took into consideration as part of their risk calculation and how risk is determined. Abbreviations: ACS NSQIP - The American College of Surgeons National Surgical Quality Improvement Program Tool; CPET - Cardiopulmonary exercise testing; RCRI - The Revised Cardiac Risk Index; POSSUM- The Physiological and Operative Severity Score for the enUmeration of Mortality and Morbidity; ASA Classification system - The American Society of Anesthesiologists Physical Status Classification System; PONV - Postoperative Nausea and Vomiting; P-POSSUM - The Portsmouth Physiological and Operative Severity Score for the enUmeration of Mortality and Morbidity; NRS-2002 - The Nutritional Risk Screening 2002; ARISCAT - The Assess Respiratory Risk in Surgical Patients in Catalonia; CFS - The Clinical Frailty Scale; SORT - The Surgical Outcome Risk Tool; DASI - The Duke Activity Status Index; GCS - Glasgow Coma Scale; ECG - Electrocardiogram; ICU – Intensive care unit.

**Table 2:**
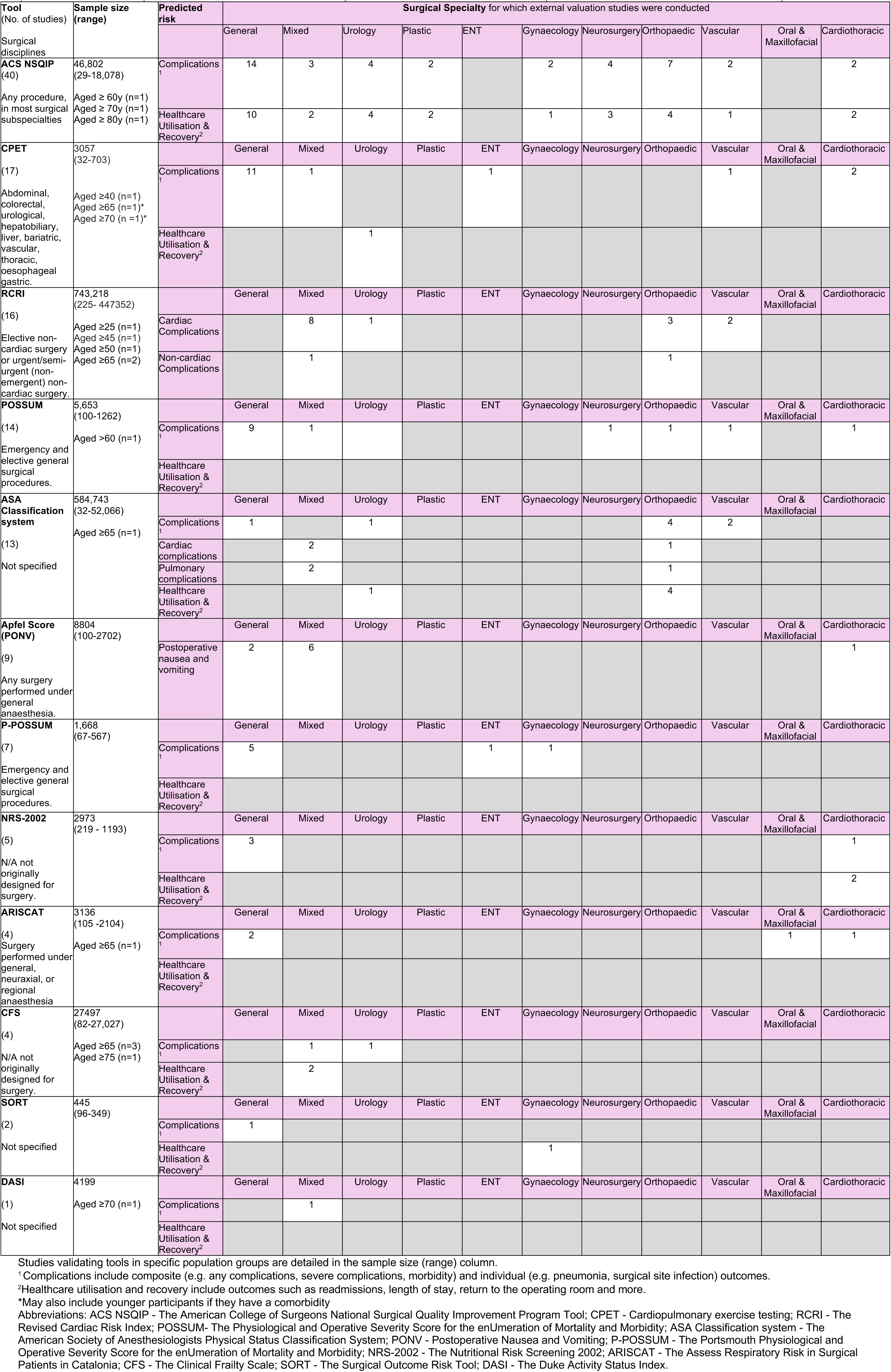
Summary of the available external validation studies for each risk prediction tool. (For a detailed description of the individual outcomes reported across studies see the risk prediction tool summaries in Section 2.2).

### 2.1 Risk prediction tools included in the Rapid Evidence Map

The evidence map focuses on 14 risk prediction tools currently utilised within the surgical setting across Wales. These were identified and provided by the stakeholders and include the following tools:

- The American College of Surgeons National Surgical Quality Improvement Program (ACS NSQIP)
- Cardiopulmonary exercise testing (CPET)
- Revised Cardiac Risk Index for Pre-Operative Risk (RCRI)
- The American Society of Anesthesiologists Physical Status (ASA) Classification System
- Apfel score (PONV)
- Portsmouth Physiological and Operative Severity Score for the enUmeration of Mortality and morbidity (P-POSSUM)
- Physiological and Operative Severity Score for the enUmeration of Mortality and morbidity (POSSUM)
- Nutritional Risk Screening 2002 (NRS-2002)
- Assess Respiratory Risk in Surgical Patients in Catalonia (ARISCAT)
- Clinical Frailty Scale (CFS)
- Surgical Outcome Risk Tool (SORT)
- Duke Activity Status Index (DASI)
- Carlisle Risk Calculator
- National Emergency Laparotomy Audit Parsimonious Risk Score (NELA-PRS)

We initially set out to identify evidence in relation to the external validation of 13 risk prediction tools provided by the stakeholders. However, we identified external validation studies for a further relevant risk prediction tool, POSSUM, which is an earlier version of the P-POSSUM tool (identified by the stakeholders). It is very similar but uses a slightly different formula to calculate risk. The POSSUM was therefore also included, which brought the total number of risk prediction tools included in this evidence map to 14. However, we did not identify evidence relating to the external validation of two of these risk prediction tools: the Carlisle Risk Calculator and the National Emergency Laparotomy Audit Parsimonious Risk Score. **Table 1** provides details about the 12 individual tools included in this evidence map, their purpose, who the tools are completed by, the surgical disciplines they are used in, and how their findings are provided.

### 2.2 Overview of the available evidence on external validation

A summary of the available evidence underpinning each tool is provided in Table 2. A total of 118 validation studies (85 retrospective and 33 prospective) met our inclusion criteria. The summary table (Table 2) shows how many studies were identified for each tool, the overall sample sizes used across studies, any specific population group being used to validate the tool, the number of studies identified for each surgical specialty, and the number of studies reporting on the different outcomes. All included studies conducted external validation evaluation of surgical risk prediction tools in adults undergoing non-emergency surgery, assessing their ability to predict complications.

Our included studies examined risk prediction tools currently used in Wales. In order of the number of included studies, these were:

- The American College of Surgeons National Surgical Quality Improvement Program (ACS NSQIP) (n=40)
- Cardiopulmonary exercise testing (CPET) (n=17)
- Revised Cardiac Risk Index for Pre-Operative Risk (RCRI) (n=16)
- Physiological and Operative Severity Score for the enUmeration of Mortality and morbidity (POSSUM) (n=14)
- Society of Anesthesiologists Physical Status (ASA) Classification System (n=13),
- Apfel score (n=9)
- Portsmouth Physiological and Operative Severity Score for the enUmeration of Mortality and morbidity (P-POSSUM) (n=7)
- Nutritional Risk Screening 2002 (NRS-2002) (n=5)
- Assess Respiratory Risk in Surgical Patients in Catalonia (ARISCAT) (n=4)
- Clinical Frailty Scale (CFS) (n=4)
- Surgical Outcome Risk Tool (SORT) (n=2)
- Duke Activity Status Index (DASI) (n=1).

Thirteen studies assessed multiple surgical risk prediction tools of interest: 12 studies assessed two tools; and one study assessed three tools. With the exception of the POSSUM and P-POSSUM tools, studies that assessed modifications of the surgical risk prediction tools were excluded. Tool modifications can be used to tailor risk prediction tools to specific surgical disciplines or medical conditions and can improve its’ predictive ability and strengthen the validity of the tool (Hageman et al., 2023). However, the inclusion of these studies would have complicated the evidence base further, so for simplicity these were excluded. Despite excluding external validation studies of tool modifications from our review, in order to reflect the totality of the evidence base for non-specific tools used within surgical settings in Wales, those that were identified during our screening process are referenced in Appendix 2.

The included validation studies were from a range of countries including; USA (n=39), UK (n=16), China (n=12), Germany (n=6), Canada (n=5), Turkey (n=5), Italy (n=4), South Korea (n=4), Australia (n=2), India (n=2), Romania (n=2), Taiwan (n=2), and one study each from Africa, Austria, Denmark, France, Ireland, Japan, Latin America, Netherlands, New Zealand, Philippines, Russia, Sri Lanka, Sweden, Tanzania and one study included data from Germany and Finland combined. The country was not reported in four studies. The sample sizes of included studies varied, ranging from 29 to 447,352. Studies assessing the RCRI had the highest number of overall participants (n=742,794), whereas SORT had the lowest number of participants (n=445).

External validity was assessed in patients undergoing various types of surgeries. Where possible, surgery types were categorised using the surgical specialties described by the Royal College of Surgeons of England (2024). However, an additional surgical specialty was also included to cover gynaecological surgeries and a ‘mixed surgeries’ category for studies that included a mixed sample of different surgery specialties. More information on the methods used to data extract, code and chart information, can be found in Section 6.4.

Most of the studies included patients undergoing general surgeries (n=43), followed by mixed surgeries (n=23), orthopaedic (n=16), cardiothoracic (n=9), urology (n=8), vascular (n=6), neurosurgery (n=5), plastic (n=2), gynaecology (n=2), ENT (n=2), urogynaecology (n=1), and oral and maxillofacial (n=1). While the majority of included studies included adults aged 18 years and over, a total of 18 studies validated the risk prediction tools in specific age groups (≥25yrs n=1; ≥40yrs n=1; ≥45yrs n=1; ≥50yrs n=1; ≥60yrs n=2; ≥65yrs n=6; ≥70yrs n=3; ≥75yrs n=1; ≥80yrs n=1).

Of the 16 UK external validation studies, the following risk prediction tools were assessed: DASI, P-POSSUM, POSSUM, CPET, and ACS NSQIP. The most commonly assessed tool was CPET (n=12 studies), followed by POSSUM (n=2). DASI, P-POSSUM, and ACS NSQIP were each assessed in one study.

Predicted outcomes of interest were categorised into complications, and healthcare utilisation and recovery. Where possible, individual categories for cardiac complications, non-cardiac complications, and postoperative nausea and vomiting were used. However, this was not always possible due to differences between tools and reporting in studies. Complications included composite (grouped outcomes e.g. any complications, severe complications) and individual outcomes (e.g. pneumonia, surgical site infection). Following the approach taken by the NICE review, morbidity was included as a composite complication (NICE 2020). Healthcare utilisation and recovery included outcomes such as readmissions, length of stay, return to the operating room and more. Each relevant outcome reported by the studies are detailed in Section 2.3.

### 2.3 Narrative Summary of the evidence base identified for each risk prediction tool

The sub-sections below outline a brief summary of each surgical risk prediction tool. They provide an overview of the tools, their uses and details the number of studies included that evaluated its external validity, the study region, participant information, surgical disciplines, and what outcomes were assessed. The summaries provide additional information to Tables 1 and 2.

#### 2.3.1 ACS NSQIP

The American College of Surgeons (ACS) National Surgical Quality Improvement Program (NSQIP) was developed in 1994 to estimate the chances of unfavourable outcomes for veterans attending surgery. It is suitable for any procedure, in most surgical subspecialties. Risk is determined through a series of questions relating to patient condition, severity and information around the surgery being conducted and individual risk scores for up to 19 different outcomes are reported included a predicted length of stay (American College of Surgeons National Surgical Quality Improvement Program, 2025).

A total of 40 studies externally validated the ACS NSQIP tool. These were published between 2016 and 2024. A wide range of countries were used including USA (n=24), Italy (n=3), Canada (n=2), South Korea (n=2), Australia (n=1), China (n=1), Latin America (n=1), Netherlands (n=1), New Zealand (n=1), Philippines (n=1), Sri Lanka (n=1), Turkey (n=1), and the UK (n=1). The sample sizes of included studies varied, ranging from 29 to 18,078. A total of 46,802 participants were included across all the studies. Most of the included studies included patients aged 18 or over (n=37), however, three studies focussed on specific age groups (≥60 n=1; ≥70 n=1; ≥80 n=1).

The ACS NSQIP was assessed across nine surgical specialties. General surgeries were most commonly studied (n=14), followed by orthopaedic (n=7), urology, (n=4) neurosurgery (n=4), mixed (n=3), plastic (n=2), thoracic (n=2), vascular (n=2), urogynaecology (n=1), and gynaecology (n=1). While all studies assessed the predictive ability of ACS NSQIP, some studies also compared the predictive ability of ACS NSQIP to other risk prediction tools of interest including the P-POSSUM (n=3) and RCRI (n=2).

Included studies assessed the ACS NSQIP’s ability to predict a broad range of outcomes including both composite and individual complications, as well as health care utilisation and recovery outcomes. This included any complication (n=24), serious complications (n=24), major complications (n=1), postoperative complications (n=1), cardiac complications (n=13), pneumonia (n=20), surgical site infection (n=19), urinary tract infections (n=19), venous thromboembolism (n=19), sepsis (n=4), renal complications (n=17), systemic complications (n=1), thyroidectomy complications (n=1). readmission (n=21), discharge to rehabilitation/skilled nursing facility or location other than home (n=18), return to operating room (n=14), length of stay (n=11), and reoperation (n=6) were assessed as health care utilisation and recovery.

#### 2.3.2 CPET

The Cardiopulmonary exercise testing (CPET) is an objective preoperative assessment of exercise capacity and aims to identify the causes of exercise limitation. The test itself consists of four main phases: rest, unloaded cycling, ramp exercise, and recovery. The dynamic metabolic challenge imposed by peri-operative CPET provides an objective means of evaluating exercise capacity. It can be used to evaluate chronic comorbidities and may enable identification of new pathology that requires treatment, optimization, or both preoperatively. It is used for a variety of different surgeries including abdominal, colorectal, urological, hepatobiliary, liver, bariatric, vascular, thoracic and oesophageal-gastric and provides scores for VO2 peak and anaerobic threshold (Levett et al., 2018).

A total of 17 studies evaluated the predictive ability of the CPET tool. Studies were published between 2008 and 2024 and were from the UK (n=12), USA (n=1), Sweden (n=1), China (n=1), Russia (n=1), and Australia (n=1). The sample sizes of included studies varied, ranging from 32 to 703. A total of 3,057 participants were included across all the studies. Most of the included studies included patients aged 18 or older, however three studies focused on specific age groups (≥40 (n=1), ≥65 or younger with a comorbidity (n=1), ≥70 or younger with a comorbidity (n=1)).

Studies assessed CPET across six surgical specialties. General surgeries were the most commonly studied (n=11), followed by cardiothoracic (n=2), ENT (n=1), mixed (n=1), urology (n=1) and vascular (n=1). One study compared the predictive ability of CPET to the ASA classification system.

Studies assessed the predictive ability of CPET predominantly for complications, with one study evaluating its ability to predict critical care unit admissions (n=1). Complication outcomes were all complications (n=3), major complications (n=1), cardiopulmonary complications (n=2), pulmonary complications (n=1), postoperative morbidity (n=5), cardiovascular complications (n=2), cardiopulmonary morbidity (n=1), postoperative complications (n=2).

#### 2.3.3 RCRI

The Revised Cardiac Risk Index for Pre-Operative Risk (RCRI) is used in patients ≥45 years old (or 18 to 44 years old with significant cardiovascular disease) undergoing elective non-cardiac surgery or urgent/semi-urgent (non-emergent) non-cardiac surgery (MDCALC, 2024). It predicts based on six risk factors: high-risk surgery, history of ischemic heart diseases, history of congestive heart failure, history of cerebrovascular disease, pre-operative treatment with insulin and pre-operative creatinine >2 mg/dL / 176.8 µmol/L and provides a single overall score for predicted postoperative cardiac complications (Lee et al.,1999).

A total of 16 studies evaluated the predictive ability of the RCRI tool. Studies were published between 2010 and 2024. Study regions included USA (n=9), Austria (n=1), Canada (n=1), Denmark (n=1), Germany (n=1), Tanzania (n=1), the Philippines (n=1), and a dataset of multiple African countries (n=1). The sample sizes of included studies varied, ranging from 225 to 447,352. A total of 743,218 participants were included across all the studies. Most of the included studies included patients aged 18 or older, however, five studies focused on specific age groups (≥25 n=1; ≥45 n=1; ≥50 n=1; ≥65 n=2).

The RCRI was assessed across four surgical specialties. Mixed surgeries were the most commonly studied (n=9), followed by orthopaedic (n=4), vascular (n=2) and urology (n=1).

While all studies assessed the predictive ability of RCRI, some studies also compared the predictive ability of RCRI to other risk prediction tools of interest including the ASA classification system (n=3) and ACS NSQIP (n=3).

Although the RCRI typically predicts the risk of major cardiac events, identified studies assessed its ability to predict a range of both cardiac and non-cardiac outcomes. Outcomes included major cardiac complications (n=8), adverse cardiac event (n=3), myocardial infarction or cardiac arrest within 30 days (n=2), postoperative cardiac morbidity (n=1), pulmonary complications (n=1), non-cardiac complications (n=1), and a composite morbidity endpoint consisting of cardiac and non-cardiac complications (n=1).

#### 2.3.4 POSSUM

The Physiological and Operative Severity Score for the enUmeration of Mortality and Morbidity (POSSUM) estimates morbidity and mortality for emergency and general surgery patients through a series of questions around patient physiological score which looks at age, cardiac and respiratory function, GCS, blood work, ECG, and operative severity, which interrogates the procedure being undertaken (MDCALC, 2024; Copeland et al.,1991). It provides a single overall score for predicted morbidity and a single overall score for predicted mortality.

A total of 14 studies evaluated the predictive ability of the POSSUM tool. Studies were published between 2003 and 2024. Study regions included China (n=5), UK (n=2), Korea (n=2), Germany (n=1), Romania (n=1). The region of three studies was unclear. The sample sizes of included studies varied, ranging from 100 to 1,262. A total of 5,653 participants were included across all the studies. Most of the included studies included patients aged 18 or older, however, one study focused on patients over 60 years.

POSSUM was assessed across six surgical specialties categories. General surgeries were the most commonly studied (n=9), followed by mixed (n=1), neurosurgery (n=1), orthopaedic (n=1), vascular (n=1) and cardiothoracic (n=1). One study compared the predictive ability of POSSUM to P-POSSUM and SORT.

Outcomes of interest were grouped as complications. These included morbidity or postoperative complications (n=14), pulmonary complications (n=1), cardiovascular complications (n=1), infectious complications (n=1) and nonfatal complications (n=1). No healthcare utilisation and recovery outcomes were assessed.

#### 2.3.5 ASA classification system

The American Society of Anesthesiologists Physical Status (ASA) Classification System assesses a patients’ pre-anaesthesia co-morbidities. While it should not be used alone, when used with other factors the classification system can be used to predict peri-operative risk and provides an ASA classification (I-VI) where a higher grade suggests higher operative risk (Saklad, 1941; American Society of Anesthesiologists, 2020).

A total of 13 studies assessed the ability of ASA classification system to accurately predict complications after surgery. Studies were published between 2006 and 2024. Study regions included the USA (n=9), Canada (n=1), Germany (n=1), Italy (n=1), and Tanzania (n=1). A total of 584,743 participants were included across all the studies, with sample sizes ranging from 32 to 52,066. The majority of studies included patients aged 18 or over but one study focused on patients aged 65 or older. While all studies assessed the predictive ability of the ASA classification system, some studies compared this to other risk prediction tools of interest including the RCRI (n=3), ARISCAT (n=1), CPET (n=1), and POSSUM (n=1) (See comparison of tools section, Section 3.1).

Orthopaedic surgeries were most commonly studied (n=6), followed by studies that included a mix of surgery types (n=3), vascular surgeries (n=2), urology surgeries (n=1), and general surgeries (n=1). Outcomes of interest included a range of composite complications (grouped complications) such as, complications, cardiac complications, and pulmonary complications. A number of healthcare utilisation and recovery outcomes including, readmission, length of stay, and discharge to higher level of care or discharge not to home, were also reported.

#### 2.3.6 Apfel Score (PONV)

The Apfel score predicts and provides an overall score for the risk of experiencing postoperative nausea and vomiting (PONV) through four questions looking at gender, smoking status, history of motion sickness or PONV, and use of postoperative opioids among patient’s undergoing general anaesthesia (Apfel et al.,1999).

A total of nine studies evaluated the ability of the Apfel score to predict postoperative nausea and vomiting. Studies were published between 1999 and 2024. Study regions included Germany (n=2), China (n=2), Germany & Finland (n=1), Turkey (n=1), France (n=1), Japan (n=1), and Taiwan (n=1). The sample sizes of included studies varied, ranging from 100 to 2,702. A total of 8,804 participants were included across all the studies and all studies included adults aged 18 or over.

Studies were included across three of the eleven surgical specialties with mixed surgeries being the most commonly studied (n=6), followed by general (n=2), and cardiothoracic (n=1). As the Apfel score only predicts the risk of postoperative nausea and vomiting no other outcomes were reported.

#### 2.3.7 P-POSSUM

The Portsmouth Physiological and Operative Severity Score for the enUmeration of Mortality and Morbidity (P-POSSUM) is a modification of the POSSUM system, that uses 12 physiological score parameters and 6 operation severity parameters. It was developed to adjust the logistic regression analysis used in POSSUM scoring to better predict mortality (Prytherch et al.,1998). However, P-POSSUM still provides a single overall score for predicted morbidity and a single overall score for predicted mortality.

A total of seven studies assessed the ability of the P-POSSUM to accurately predict complications after surgery. Studies were published between 2018 and 2024. Study regions included Turkey (n=2), China (n=1), Ireland (n=1), Italy (n=1), Romania (n=1), and the UK (n=1). A total of 1,668 participants were included across studies, with sample sizes ranging from 67 to 567. All studies included adults aged 18 or over. Four studies also compared the predictive ability of P-POSSUM to other risk prediction tools of interest (POSSUM and SORT n=1 and ACS NSQIP n=3).

The seven included studies included three different surgery types, these were general surgeries (n=5), ENT (n=1) and gynaecology surgeries (n=1). Outcomes included mostly composite complications. This included, any complications (n=2), severe complications (n=1), morbidity (n=4) and pancreaticoduodenectomy morbidity (n=1).

#### 2.3.8 NRS-2002

The Nutritional Risk Screening 2002 (NRS-2002) predicts risk of malnutrition in hospitalised patients through four screening questions relating to weight, diet and patient ICU status and provides an overall score for malnutritional risk (Kondrup et al., 2003). This risk prediction tool does not appear to be specific to surgery.

A total of five studies evaluated the predictive ability of the NRS-2002. Studies were published between 2012 and 2022. Study regions included Germany (n=2), China (n=1), Turkey (n=1), and India (n=1). The sample sizes of included studies varied, ranging from 219 to 1,193. A total of 2,973 participants were included across all the studies and all included adults aged 18 or over.

Samples included in studies underwent mixed surgeries (n=3), and cardiothoracic surgery (n=2). Included studies assessed the NRS-2002’s ability to predict a range of outcomes. These included complications (n=1), postoperative complications (n=2), postoperative pulmonary complications (n=1), anastomotic leakage (n=1), infectious complications (n=1), overall complications (n=1) and major complications (n=1). Healthcare utilisation and recovery outcomes included ICU Stay >4 Days (n=1), ICU Stay >5 Days (n=1), readmission to the ICU (n=1), and delayed hospital discharge (n=1).

#### 2.3.9 ARISCAT

The Assess Respiratory Risk in Surgical Patients in Catalonia (ARISCAT) predicts risk of pulmonary complications after surgery, including respiratory failure. This tool evaluates based on seven risk factors: oxygen saturation levels, respiratory infections, age, anaemia and surgery information such as surgical incision, duration of surgery and emergency procedure. It was developed for patients undergoing general, neuraxial or regional anaesthesia and provides a single overall score for predicted postoperative pulmonary complications (Canet et al., 2010).

A total of four studies assessed the ability of the ARISCAT tool to accurately predict complications after surgery. Studies were published between 2020 and 2024. Study regions included India (n=1), Italy (n=1), USA (n=1) and it was unclear in one study. A total of 3,136 participants were included across studies, with sample sizes ranging from 105 to 2,104. Three studies included patients aged 18 or older while one study focused on patients aged 65 or older. While all studies assessed the predictive ability of the ARISCAT tool, one study also compared the predictive ability of ARISCAT to the ASA classification system.

Studies included general surgeries (n=2), oral and maxillofacial surgeries (n=1) and cardiothoracic (n=1). The outcome of interest in all studies was postoperative pulmonary complications (n=3).

#### 2.3.10 CFS

The Clinical Frailty Scale (CFS) was originally developed to summarise the overall level of fitness or frailty of an older adult. It measures frailty to predict survival, medium- and long-term outcomes (mortality and need for institutional care) through an inclusive 9-point scale and provides an overall frailty score. Higher scores indicate greater risk. It focuses on items including mobility, balance, use of walking aids, and the ability to eat, dress, shop, cook, and bank (Rockwood et al., 2020). This risk prediction tool does not appear to be specific to surgery.

A total of four studies evaluated the predictive ability of the CFS. Studies were published between 2020 and 2023. Study regions included China (n=2), Canada (n=1), and Taiwan (n=1). The sample sizes of included studies varied, ranging from 82 to 27,027. A total of 27,497 participants were included across all the studies. All four studies focused on specific age groups (≥65 n=3; ≥75 n=1).

Studies mainly included mixed surgeries (n=3) but also urology (n=1). Outcomes of interest included complications: major complication (n=1) and postoperative complications (n=1); and healthcare utilisation and recovery: prolonged hospital stays (10+ days) (n=1), unplanned hospital readmission (30 days) (n=1), long-term hospitalisation (90 days) (n=1), and long-term care admission (1 year) (n=1).

#### 2.3.11 SORT

The Surgical Outcome Risk Tool (SORT) comprises of six variables (ASA classification system grade, urgency of surgery, high-risk surgical specialty, surgical severity, cancer and age 65 years or over) and provides an overall risk prediction score of death within 30 days of surgery. It was developed for adults undergoing surgery (Protopapa et al., 2014).

A total of two studies assessed the ability of the SORT tool to accurately predict complications after surgery. Studies were published between 2023 and 2024. Study regions included China (n=1) and Turkey (n=1). A total of 445 participants were included across studies, with sample sizes ranging from 96 to 349. All studies included adults aged 18 or over. While both studies assessed the predictive ability of the SORT tool, one study compared the predictive ability of SORT to other risk prediction tools of interest including POSSUM and P-POSSUM.

Studies included general (n=1) and gynaecology surgeries (n=1). Outcomes of interest included complications; early complication (n=1) and healthcare utilisation & recovery; postoperative admission to ICU (n=1).

#### 2.3.12 DASI

The Duke Activity Status Index (DASI) is a 12-item, self-administered questionnaire that measures functional capacity and assesses quality of life developed for use during preoperative assessments (Hlatky et al., 1989). It provides a single overall score for functional capacity.

One study was identified that assessed the ability of DASI to accurately predict complications after surgery. The study was published in 2024 and was conducted in the UK assessing 4,199 participants aged 70 years or above. Surgeries included a mix of non-cardiac surgeries. Outcomes of interest included complications, more specifically the need for a blood transfusion after surgery.

## 3. Summary of the findings for ACS NSQIP, P-POSSUM, RCRI and the ASA classification system

It became clear that not all of the risk prediction tools were specifically developed for surgery and therefore may not be appropriate for the identification of patients suitable for treatment in low-risk surgical settings such as surgical hubs. Given the large evidence base identified, this summary of findings focusses on four of the risk prediction tools (ACS NSQIP, RCRI, P-POSSUM and the ASA classification system). These tools were selected as being potentially the most impactful at a population level and P-POSSUM was selected over POSSUM for inclusion in the summary as it is the most recent version of the tool. Following the approach utilised by the NICE review (NICE 2020), composite outcomes were used to compare the predictive ability of each tool. Studies that report mortality as part of their composite complications were not included to allow for a more direct comparison. Some studies reported multiple composite outcomes, in this case the largest composite (including the most complications but not including mortality) was taken for the results. A full breakdown of the different composite complications and individual complications reported, along with the findings have been tabulated for each tool in Appendix 3. In addition, where more than one study was reported for a surgical specialty, the findings have also been tabulated in Appendix 3 for each tool.

This section begins with a summary of the findings from the studies that directly compared multiple risk prediction tools (Section 3.1). This is followed by a breakdown of the overall findings for each of the tools of interest (Section 3.2 – 3.5). This includes a look at any findings related to the discrimination, calibration and accuracy of the risk prediction tools, along with findings in relation to the different surgical specialties and a summary of any healthcare utilisation and recovery outcomes that were reported.

Discrimination measures how well a tool differentiates between patients who do and do not experience an event, quantified using the area under the receiver operating characteristic (ROC) curve (AUC) or c-statistic. Within the literature, a c-statistic of 90% or greater is typically considered an excellent level of discriminative ability, a c-statistic of 80% or greater is considered a good level of discriminative ability, a c-statistic of 70% or greater is considered a fair level of discriminative ability, a c-statistic of 60% or greater is considered a poor level of discriminative ability and a c-statistic of 50% or greater is considered a very poor level of discriminative ability (or no better than chance) (NICE 2020; Çorbacıoğlu and Aksel, 2023).

Calibration assesses the agreement between the predicted and observed outcomes, typically presented graphically as observed risks versus predicted risks or in tabular format (Collins et al., 2014). Following the approach utilised by the NICE review, an observed/expected (O/E) ratio of 0.9-1.1 would be considered a fair level of calibration.

In the literature, the accuracy of the risk prediction tools was assessed using the ‘Brier Score’. The Brier score is a simultaneous measure of calibration and discrimination. It is reported as a score between 0 and 1. A score of 0 indicates no difference between the predicted and actual outcome, thus indicating the best possible test result. A score of 1 indicates that the test did not predict the outcome. The Brier score is compared with a Brier score cut-off, which is partially based on the incidence in the sample, and a score above the cut-off is considered not useful (Alzahrani et al., 2020). Brier score cut-offs are dependent on the datasets of individual studies, as the value of the Brier score depends on both the prevalence of the event in the data and the performance of the model (Huang et al., 2021). As such, direct comparisons across studies may not be appropriate, for the purposes of this in-depth summary, the range of the Brier scores reported across studies will be highlighted.

### 3.1 Studies comparing the predictive ability of multiple risk prediction tools

A total of 9 studies compared three of the four risk prediction tools of interest (See Table 3 below). Three studies compared the predictive ability of the ASA classification system to the RCRI, three studies compared ACS NSQIP with P-POSSUM, and three studies compared ACS NSQIP with the RCRI tool. These assessed each tools’ predictive ability using a variety of outcomes in several different surgical disciplines.

**Table 3.** Results of studies comparing multiple tools.

| Outcome | Reference | Surgery | Risk Prediction Tool |  |  |  | P value |
| --- | --- | --- | --- | --- | --- | --- | --- |
|  |  |  | ASA | P-POSSUM | ACS NSQIP | RCRI |  |
| **Morbidity** | Karabulut et al., (2024) | General |  | AUC ▼<br>Brier Score 0.2567<br>O/E event ratio 1.48 | AUC ◀▶<br>Brier Score 0.2156<br>O/E event ratio 1.36. |  | ACS NSQIP:<br>0.186<br>P-POSSUM:<br>0.352 |
| **Morbidity** | Sevinyan et al., (2024) | Gynaecology |  | AUC ▼<br>Brier score 0.1359<br>O/E event ratio 0.183 | AUC ▼<br>Brier score 0.1827<br>O/E event ratio 0.136 |  | P-POSSUM's overestimation of morbidity was statistically significant (0.018) |
| **Any complication** | Angelucci et al., (2024) | General |  | AUC ▼<br>Brier Score 0.229 | AUC ▼<br>Brier Score 0.231 |  | - |
| **Severe complication** | Angelucci et al., (2024) | General |  | AUC ▼<br>Brier Score 0.205 | AUC ▼<br>Brier Score 0.206 |  | - |
| **Any complication** | Bronheim et al., (2018) | Spinal | AUC ◀▶ |  |  | AUC ▼ | <0.001 |
| **Cardiac complications\*** | Bronheim et al., (2019) | Spinal | AUC ◀▶ |  |  | AUC ▲ | 0.0004 |
| **Cardiac complications** | Christant et al., (2024) | Mixed | AUC ◀▶ |  |  | AUC ◀▶ | 0.817 |
| **Pulmonary complications** | Christant et al., (2024) | Mixed | AUC ◀▶ |  |  | AUC ◀▶ | 0.469 |
| **Major cardiac complications 30 day** | Cohn and Ros (2018) | Mixed |  |  | AUC ◀▶ | AUC ▼ | - |
| **All cardiac complications in-hospital** | Cohn and Ros (2018) | Mixed |  |  | AUC ▲ | AUC ▲ | - |
| **All cardiac complications 30 day** | Cohn and Ros (2018) | Mixed |  |  | AUC ▲ | AUC ◀▶ | - |
| **Adverse cardiac events** | Moses (2019) | Vascular |  |  | O/E event ratio 1.60 | O/E event ratio of 2.23 | - |
| **Cardiac complication/ MACE (major adverse cardiac event)** | Yap et al., (2018) | Mixed |  |  | AUC ▲<br>Brier Score 0.08 | AUC ▲ | - |
| Effect direction: ▲=Good to excellent (80%+) ◀▶= Fair (70-79%) ▼=Poor (50-60%) |  |  |  |  |  |  |  |
| * Denotes the mean of two individual outcomes reported in a study. These were cardiac arrest requiring CPR and myocardial infarction<br>O/E: Observed/Expected event ratio<br>-: information not provided by study |  |  |  |  |  |  |  |

#### ASA and RCRI

When comparing the predictive ability of the ASA classification system and the RCRI the findings were mixed. Of the three studies comparing the ASA classification system and the RCRI, none reported any calibration or accuracy findings, only reporting the discriminative ability of the tools (c-statistics). One study (Chrisant et al., 2024) reported both tools to have a fair discriminative ability in predicting cardiac and pulmonary complications in elective non-cardiothoracic surgery. One study (Bronheim et al., 2018) reported the ASA classification system to have a fair discriminative ability whereas the RCRI was found to have a poor discriminative ability to predict a composite of non-cardiac complications after posterior lumbar decompression. However, a second study from the same author (Bronheim et al.,2019), found the RCRI to have a good discriminative ability for predicting myocardial infarction and cardiac arrest requiring cardiopulmonary resuscitation (CPR), whereas the ASA classification system was found to have a fair and poor discriminative ability, respectively.

#### ACS NSQIP and P-POSSUM

When comparing the predictive ability of ACS NSQIP and P-POSSUM the findings were mixed. One study reported a poor discriminative ability and inaccuracy for both the P-POSSUM tool and ACS NSQIP after retroperitoneal sarcoma surgery (Angelucci et al., 2024). However, ACS NSQIP was found to have a fair discriminative ability compared to P-POSSUM’s poor discriminative ability in another study after gynaecological surgery (Karabulut et al., 2024). Calibration was not reported in either study. Lastly, one study found ACS NSQIP to have a poor discriminative ability compared to P-POSSUM’s very poor discriminative ability for predicting complications after hepatobiliary surgery (Sevinyan et al., 2024), however, the calibration was slightly better in the P-POSSUM tool compared with ACS NSQIP for morbidity.

#### ACS NSQIP and RCRI

Finally, three studies compared the predictive ability of ACS NSQIP and the RCRI and the findings were mixed. One study in four cardiac surgeries (Cohn and Ros, 2018) found good discriminative ability for ‘All cardiac complications in-hospital’ for both tools. In the same study, the ACS NSQIP tool had a fair discriminative ability in ‘Major cardiac complications 30 day’ compared to poor discriminative ability for RCRI for the same outcome. For the outcome of ‘All cardiac complications 30 day’ the ACS NSQIP tool had good discriminative ability compared to a fair discriminative ability in the RCRI tool. This study did not report calibration or accuracy. In the second study comparing ACS NSQIP with the RCRI in vascular surgery (Moses et al., 2019), only calibration was reported. For the outcome of ‘Adverse Cardiac Events’ the ACS NSQIP tool reported a ratio of 1.60 of observed to predicted adverse events; the RCRI tool reported a ratio of 2.23 for the same outcome suggesting both tools underpredicted adverse events. In a third study in mixed surgeries (Yap et al., 2018) both tools had excellent discriminative ability for the outcome of ‘Cardiac complication/ MACE (major adverse cardiac event)’. This study also reported excellent calibration in the ACS NSQIP tool for the same outcome, while calibration scores were not reported for the RCRI tool.

### 3.2 ACS NSQIP findings

#### 3.2.1 ACS NSQIP Discrimination findings

A total of 33 studies reported c-statistics on composite complications for ACS NSQIP. Two studies (Chudgar et al., 2021; Chudgar et al., 2022) appear to use the same data so have only been reported once. Six studies reported multiple c-statistics for different surgeries (Campagnaro et al., 2023; Fruscione et al., 2018; Houdek et al., 2020; Labbot et al., 2021a; Labbot et al., 2021b; Rivard et al., 2016), details of the different surgeries reported can be seen in Figure 1 which plots the c-statistics taken from each paper. **Overall, c-statistics ranged from 45% to 93% with a median c-statistic of 60.35% suggesting ACS NSQIP had a poor predictive ability for composite complications across all 33 studies.**

**Figure 1.**
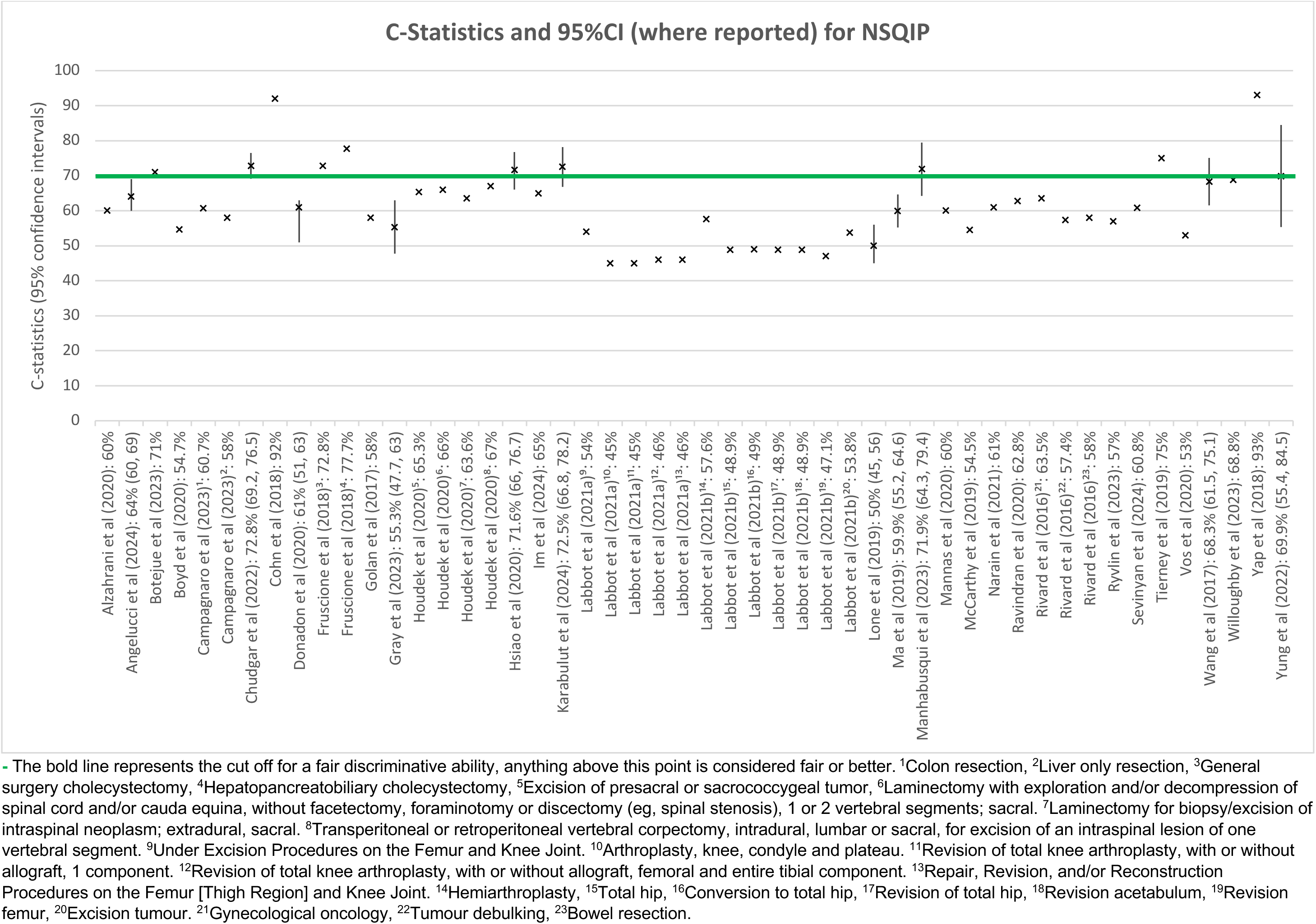
ACS NSQIP C-statistic Plot.

The majority of studies were conducted in populations receiving general surgeries (n=12), followed by orthopaedic surgeries (n=6), neurosurgery (n=4), urology (n=3), mixed surgeries (n=3) and plastic surgeries (n=2). One study included gynaecology surgeries, one study included vascular surgeries, and one study included thoracic surgeries. The findings by surgery type can be seen in Table 4.

**Table 4.** C-statistic results for ACS NSQIP by surgical specialty.

| Surgical specialty | No of studies | c-statistic range | Median c-statistic | Discriminative ability |
| --- | --- | --- | --- | --- |
| General surgery | 12 | 53% - 77.7% | 63.15% | Poor |
| Orthopaedic surgery | 6 | 45% - 71.9% | 49.5% | Very poor |
| Neurosurgery | 4 | 57% - 68.3% | 65.3% | Poor |
| Urology surgery | 3 | 50% - 60% | 58%% | Very poor |
| Mixed surgery | 3 | 54.7% - 93% | 92% | Excellent |
| Plastic surgery | 2 | 69.9% - 75% | 72.45% | Fair |
| Thoracic surgery | 1 | N/A | 72.8% | Fair |
| Vascular surgery | 1 | N/A | 59.9% | Very poor |
| Gynaecology surgery | 1 | N/A | 60.8% | Poor |

#### 3.2.2 ACS NSQIP Calibration findings

A total of 21 studies reported the number of observed complications (O) and the number of complications predicted (E) by ACS NSQIP (O/E ratio). Two studies (Chudgar et al., 2021; Chudgar et al., 2022) appear to have used the same data, so are only been reported once below. One study reported multiple O/E ratios for different surgeries (Fruscione et al., 2018) details of the different surgeries reported can be seen in Figure 2 which plots the O/E ratios taken from each study The O/E ratio ranged from 0.737 to 11.833. **Overall, the median O/E ratio for ACS NSQIP was 1.552.**

**Figure 2.**
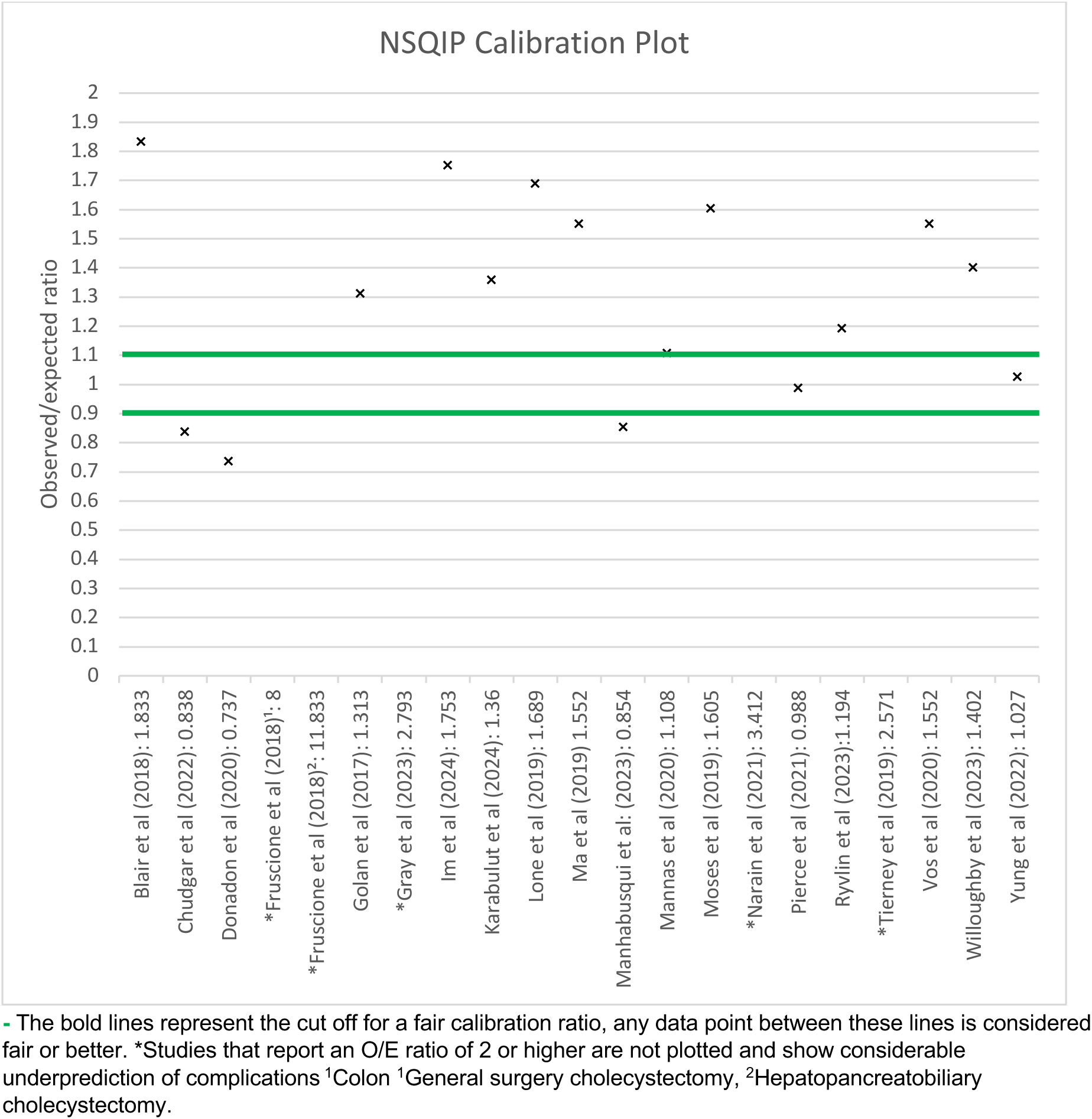
ACS NSQIP calibration plot.

The majority of studies were conducted in populations receiving general surgery (n=5), followed by orthopaedic surgery (n-4), urology surgery (n=4), neurosurgery (n=2), vascular surgery (n=2), plastic surgery (n=2), and one study included thoracic surgery. The calibration findings for ACS NSQIP by surgical specialty are displayed in Table 5.

**Table 5.**
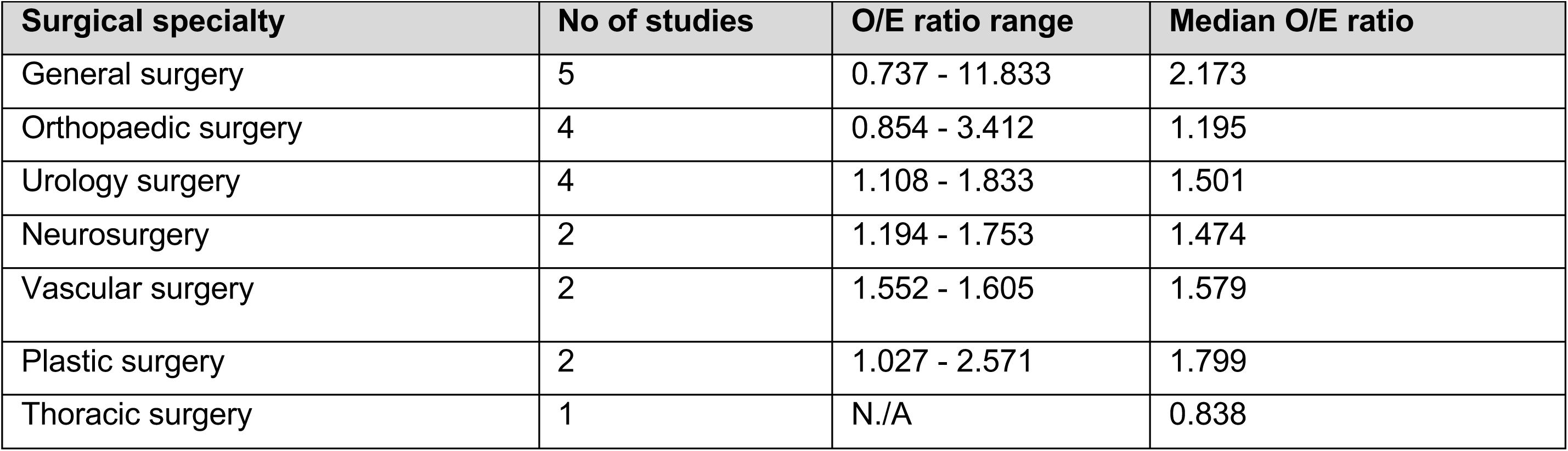
ACS NSQIP calibration findings by surgical specialty.

| Surgical specialty | No of studies | O/E ratio range | Median O/E ratio |
| --- | --- | --- | --- |
| General surgery | 5 | 0.737 - 11.833 | 2.173 |
| Orthopaedic surgery | 4 | 0.854 - 3.412 | 1.195 |
| Urology surgery | 4 | 1.108 - 1.833 | 1.501 |
| Neurosurgery | 2 | 1.194 - 1.753 | 1.474 |
| Vascular surgery | 2 | 1.552 - 1.605 | 1.579 |
| Plastic surgery | 2 | 1.027 - 2.571 | 1.799 |
| Thoracic surgery | 1 | N/A | 0.838 |

#### 3.2.3 ACS NSQIP Accuracy findings

A total of 22 studies assessing ACS NSQIP reported a Brier score for composite complications (See Figure 3). **Overall, the Brier scores for ACS NSQIP ranged from 0.00000196 to 0.722.**

**Figure 3.**
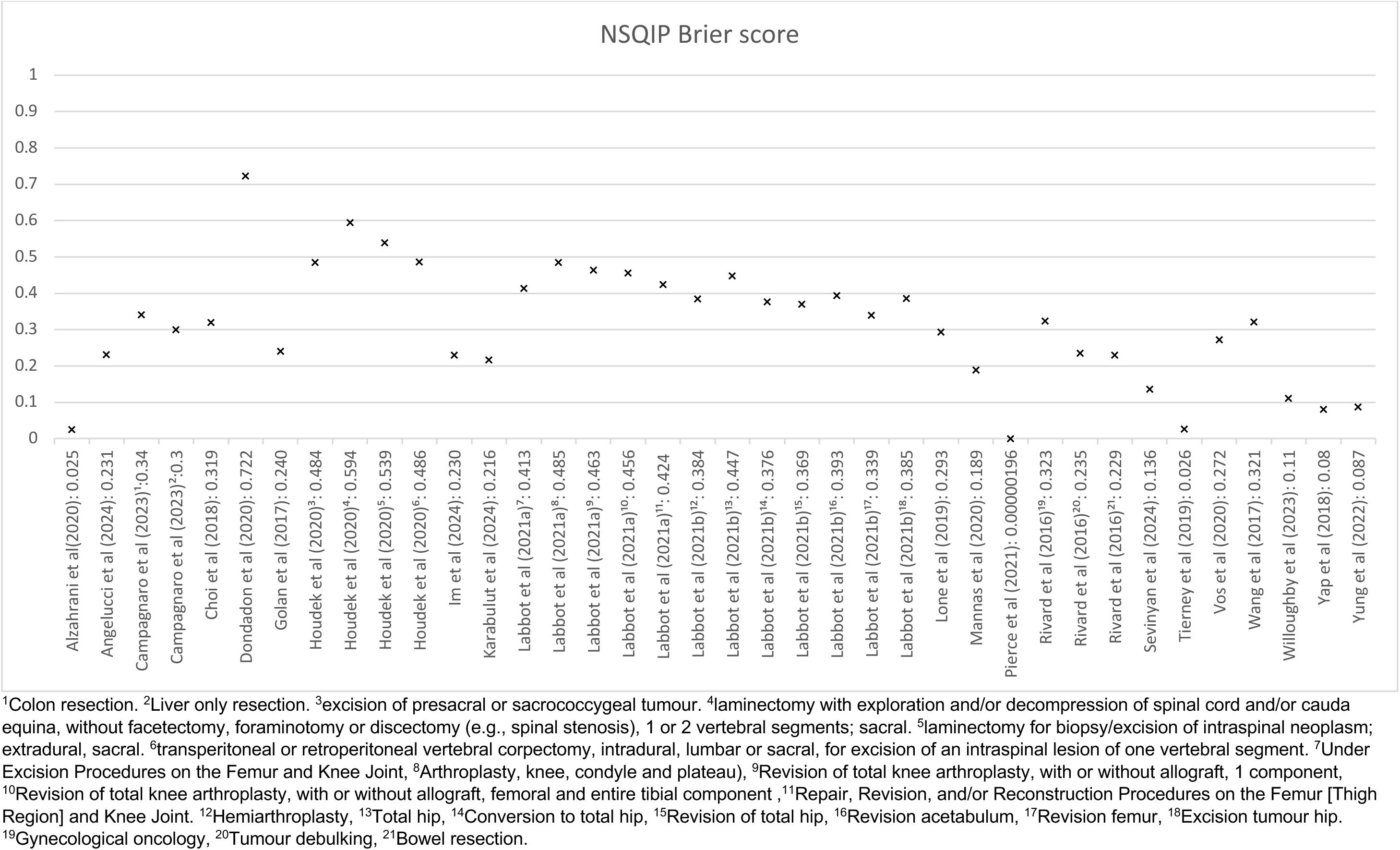
ACS NSQIP Brier score plot.

The majority of studies were conducted in populations receiving general surgery (n=8), followed by orthopaedic surgery (n=4), neurosurgery (n=3), urology surgery (n=3), plastic surgery (n=2), one study included mixed surgery, and one study included gynaecology surgery. Accuracy findings by surgery type can be seen in Table 6.

**Table 6.** ACS NSQIP Brier results by surgical specialty.

| Surgical specialty | No of studies | Brier score range |
| --- | --- | --- |
| General surgery | 8 | 0.025 - 0.722 |
| Orthopaedic surgery | 4 | 0.00000196 - 0.485 |
| Neurosurgery | 3 | 0.230 - 0.594 |
| Urology surgery | 3 | 0.189 - 0.293 |
| Plastic surgery | 2 | 0.026 - 0.087 |
| Mixed surgery | 1 | 0.08 |
| Gynaecology surgery | 1 | 0.136 |

#### 3.2.4 ACS NSQIP Healthcare utilisation and recovery outcomes

A total of 29 of the 40 studies assessing the predictive ability of ACS NSQIP reported five healthcare utilisation and recovery outcomes. The findings for all healthcare utilisation and recovery outcomes can be seen in Appendix 3. This included readmission, return to operating room, length of stay, discharge to a facility other than home and adverse discharge. The studies included all surgical specialities, apart from ENT and oral & Maxillofacial surgery. While discrimination, calibration and accuracy scores were reported, they were not consistently reported by each study, with the c-statistic being the most commonly reported measure. The findings varied considerably across studies with wide ranges being reported however, overall ACS NSQIP was found to be very poor at predicting readmission (59% across 24 studies); return to operating room (57.8% across 25 studies) and length of stay (56% across 7 studies). ACS NSQIP was found to have a fair discriminative ability for discharge to a facility other than home across 17 studies (ranging from 58.5% to 90% with a median c-statistic of 70%) and a fair calibration ratio for discharge to a facility other than home (ranging from 0.129 to 3.630, with a median O/E ratio of 0.938) however, the large ranges highlight these findings were not consistently reported across studies and further evidence would be needed.

#### 3.2.5 Bottom line summary for ACS NSQIP

There is evidence to suggest that when looking at composite complications ACS NSQIP had a poor discriminative ability across 33 studies overall (median c-statistic of 60.35%). When looking at the discriminative ability of ACS NSQIP by surgical specialty the c-stats varied, ranging from very poor (orthopaedic, urology and vascular surgery) to excellent (mixed surgery). However, the findings for the individual surgical specialties are limited and showed wide ranges across studies, reducing the confidence in the findings. As such, the findings should be interpreted with caution.

Calibration findings suggest ACS NSQIP under predicted complications overall across 21 studies (median O/E ratio of 1.552). ACS NSQIP was found to under predict complications across all surgical specialties assessed apart from thoracic surgery where it was found to over predict complications (O/E ratio of 0.838), however this finding was only reported by one study and so further evidence would be needed to confirm this. When looking at accuracy the Brier scores for ACS NSQIP ranged from 0.00000196 to 0.722 across 22 studies.

The findings for ACS NSQIP’s ability to predict healthcare utilisation and recovery outcomes also varied considerably across studies with wide ranges being reported, suggesting further evidence is needed. However overall, there is evidence to suggest ACS NSQIP was very poor at predicting readmission; return to operating room and length of stay and fair at predicting discharge not to home.

The evidence directly comparing the predictive ability of ACS NSQIP to P-POSSUM or the RCRI was limited, and the findings appear to be mixed, suggesting further evidence is needed.

### 3.3 P-POSSUM findings

#### 3.3.1 P-POSSUM Discrimination findings

A total of 7 studies reported c-statistics for the P-POSSUM on composite complications (See Figure 4). **Overall c-statistics ranged from 55.1%-78.1% with a median c-statistic of 67.2% suggesting P-POSSUM had a poor predictive ability for composite complications across all seven studies.**

**Figure 4.**
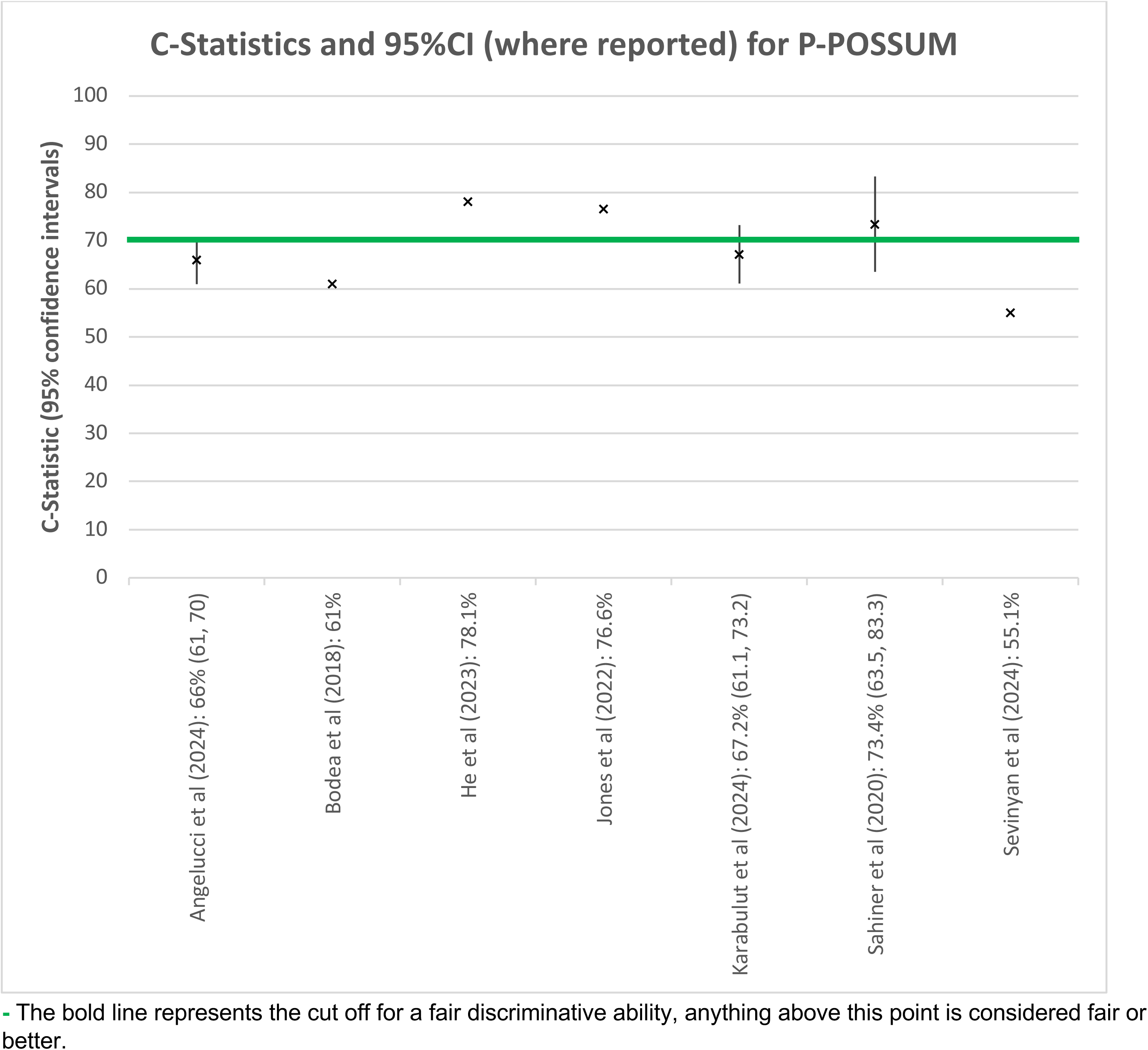
P-POSSUM C-Statistics plot.

The majority of studies were conducted in populations receiving general surgeries (n=5), one study included ENT surgeries, and one study included gynaecology surgeries. The findings by surgery type can be seen in Table 7.

**Table 7.** P-POSSUM c-statistics by surgical specialty.

| Surgical specialty | No of studies | c-statistic range | Median c-statistic | Discriminative ability |
| --- | --- | --- | --- | --- |
| General surgery | 5 | 61% - 78.1% | 67.2% | Poor |
| Gynaecology surgery | 1 | N/A | 55.1 | Very poor |
| ENT surgery | 1 | N/A | 76.6% | Fair |

#### 3.3.2 P-POSSUM Calibration findings

A total of three studies reported the number of observed complications and the number of complications predicted by P-POSSUM (O/E ratio) (See Figure 5). **Overall, the median O/E ratio of P-POSSUM for composite complications was 1.480 across all three studies.**

**Figure 5.**
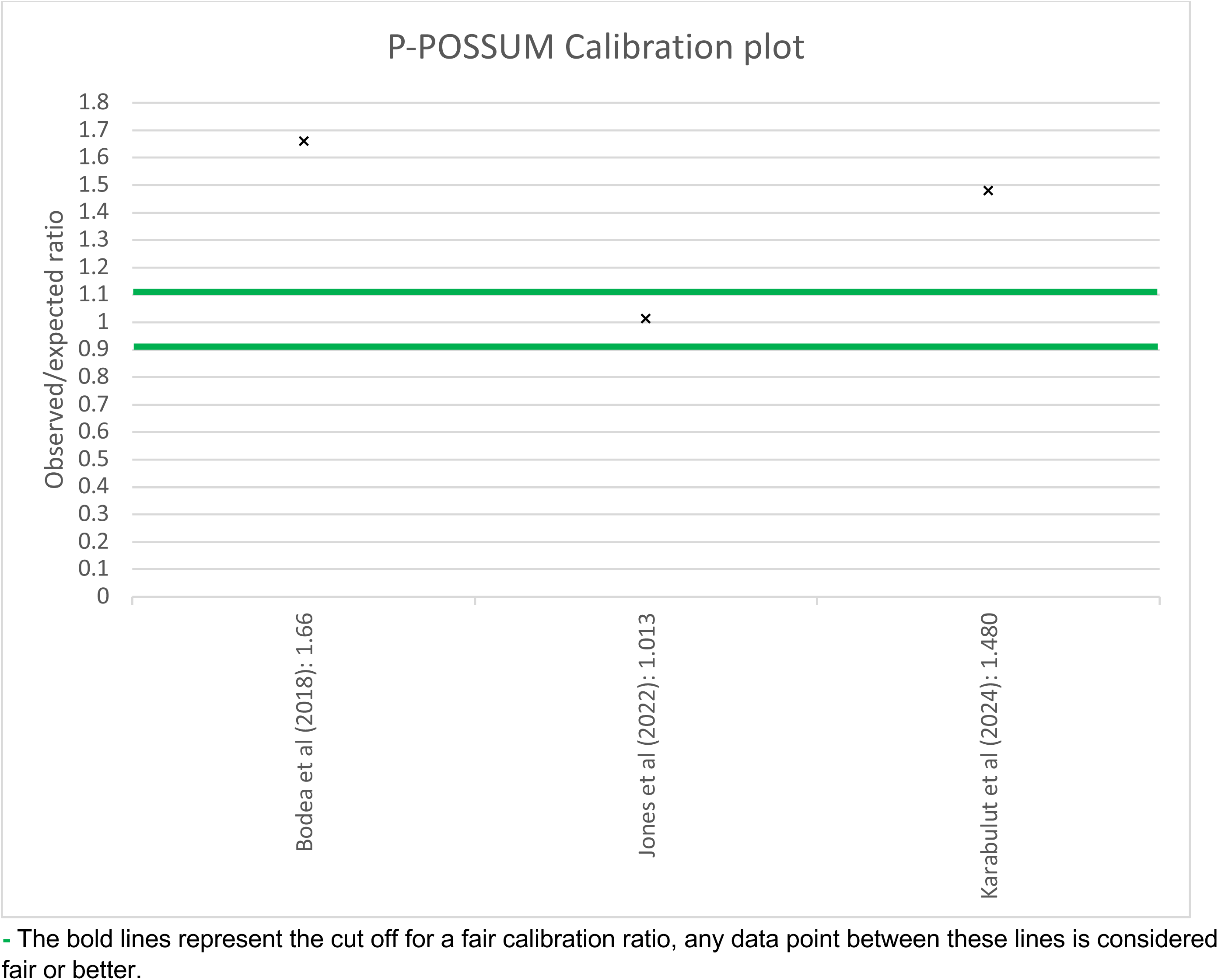
P-POSSUM calibration plot.

Two of the studies included populations receiving general surgery and one study included ENT surgery. The P-POSSUM calibration findings by surgical specialties can be seen in Table 8.

**Table 8.** P-POSSUM calibration findings by surgical specialty.

| Surgical specialty | No of studies | O/E ratio range | Median O/E ratio |
| --- | --- | --- | --- |
| General surgery | 2 | 1.48 - 1.66 | 1.57 |
| ENT surgery | 1 | N/A | 1.013* |

\*Fair level of calibration.

#### 3.3.3 P-POSSUM Accuracy findings

A total of three studies assessing P-POSSUM reported a Brier score for composite complications (See Figure 6). **Overall, the Brier scores for P-POSSUM ranged from 0.183 – 0.257 across all three studies.**

**Figure 6.**
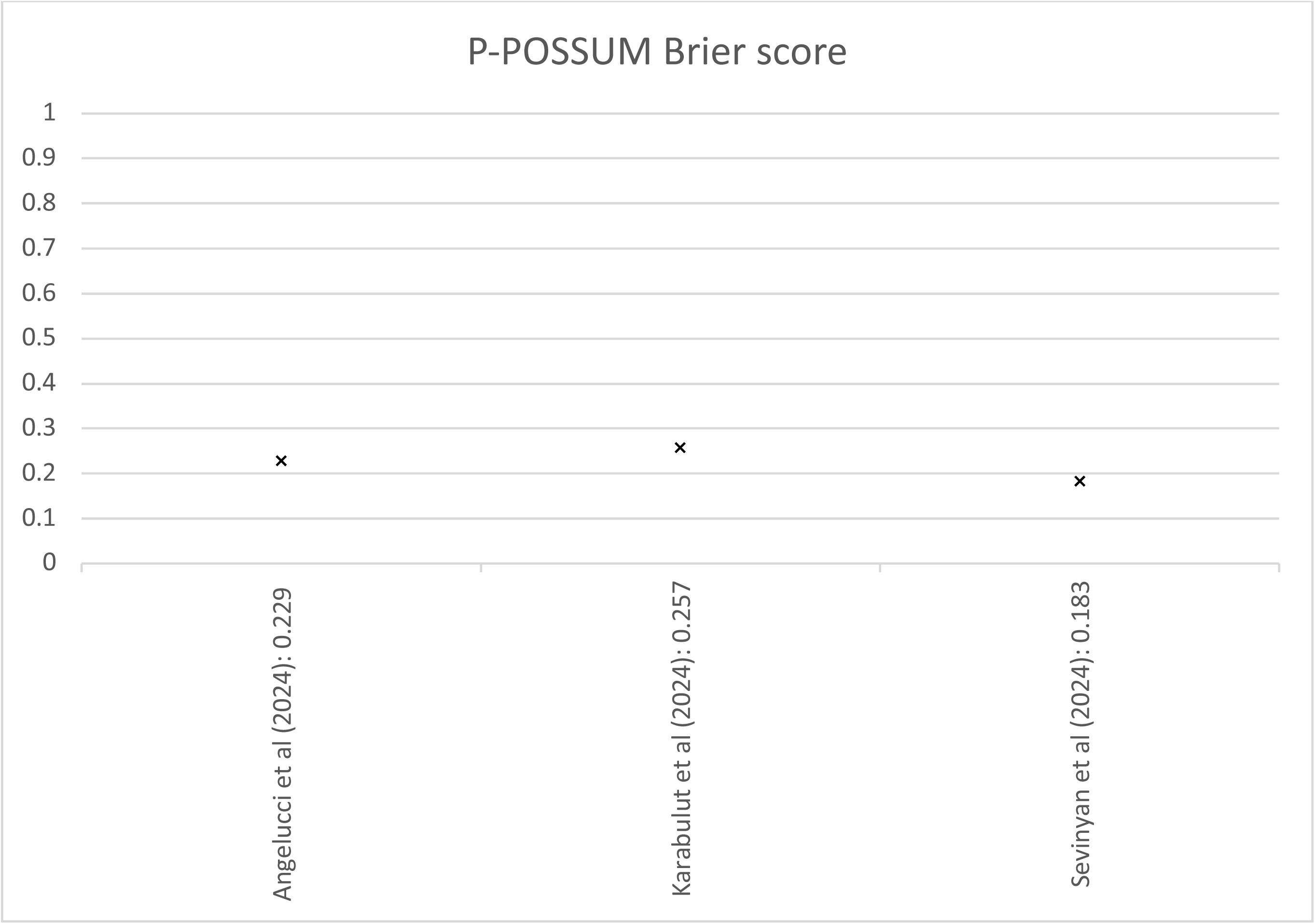
P-POSSUM Brier score plot.

Two of the studies included populations receiving general surgery and one study included gynaecology surgery. Accuracy findings by surgery type can be seen in Table 9:

**Table 9.** P-Possum Brier score by surgical specialty.

| Surgical specialty | No of studies | Brier score range |
| --- | --- | --- |
| General surgery | 2 | 0.229 - 0.257 |
| Gynaecology surgery | 1 | 0.183 |

#### 3.3.4 P-POSSUM healthcare utilisation and recovery outcomes

No studies assessing the predictive ability of P-POSSUM reported any healthcare utilisation and recovery outcomes.

#### 3.3.5 Bottom line summary for P-POSSUM

There is evidence to suggest that when looking at composite complications P-POSSUM had a poor discriminative ability across seven studies (median c-statistic 67.2%). When looking at the discriminative ability of P-POSSUM by surgical specialty the c-stats varied, ranging from very poor in one study after gynaecology surgery to fair in one study after ENT surgery. However, the findings for the individual surgical specialties are very limited, reducing the confidence in the findings, and further evidence is needed. Calibration findings suggest P-POSSUM under predicted complications across three studies overall (with a median O/E ratio of 1.480). P-POSSUM was found to under predict complications in general surgery across two studies but was found to have a fair calibration score in one study after ENT surgery (1.013). In terms of accuracy the Brier scores for P-POSSUM ranged from 0.183 – 0.257 across three studies.

No studies assessing the predictive ability of P-POSSUM reported any healthcare utilisation and recovery outcomes. The evidence directly comparing the predictive ability of P-POSSUM compared to ACS NSQIP was limited and the findings appear to be mixed, suggesting further evidence is needed.

### 3.4 RCRI findings

#### 3.4.1 RCRI Discrimination findings

A total of 13 studies reported c-statistics on composite complications for the RCRI tool (See Figure 7). One study reported c-statistics for the predictive ability of RCRI for cardiac complications and pulmonary complications (Chrisant et al, 2024). **C-statistics ranged from 54%-93% with a median c-statistic of 72% suggesting the RCRI had a fair predictive ability for composite complications across all 13 studies.**

**Figure 7.**
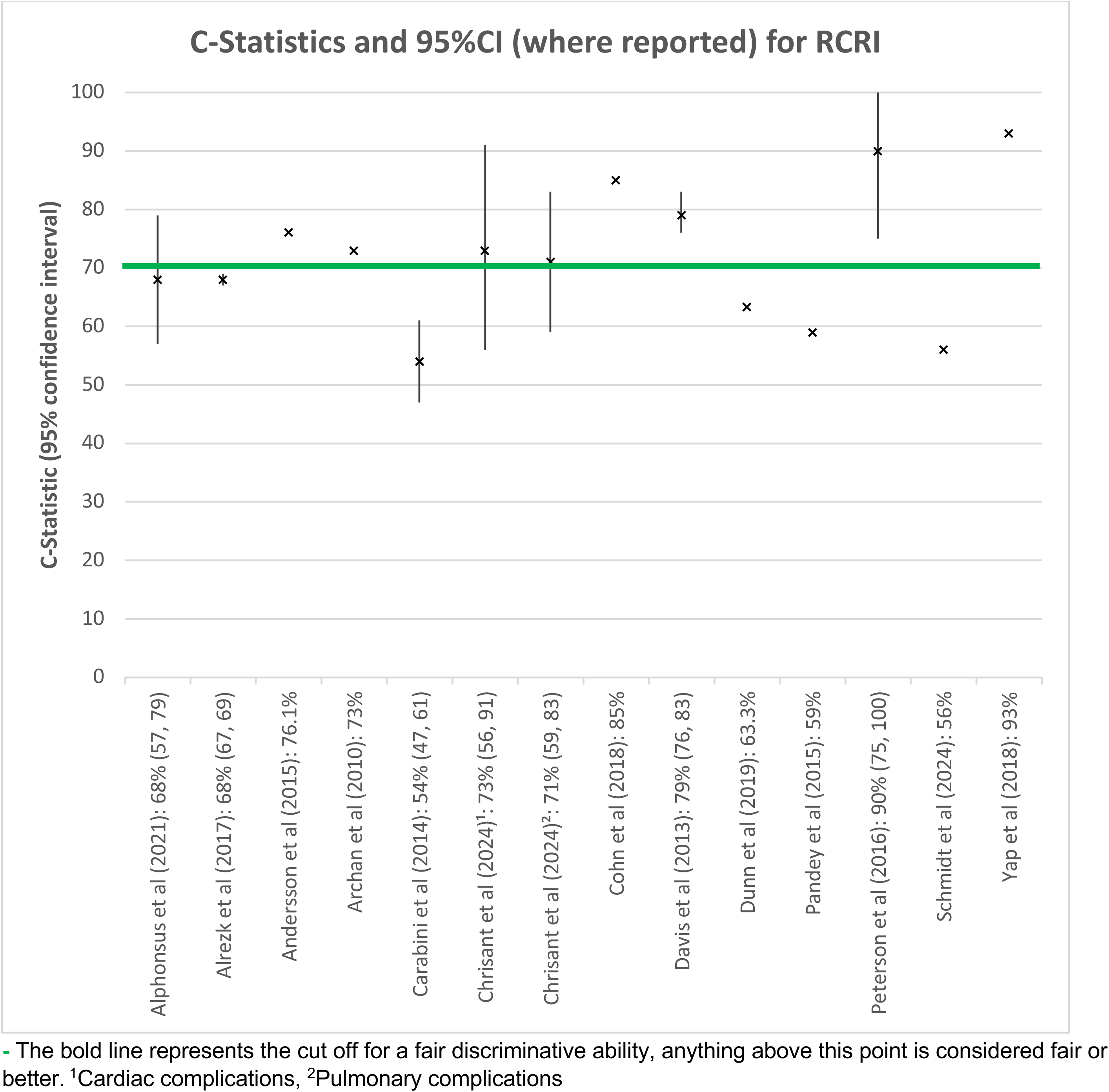
RCRI C-Statistics plot.

The majority of studies were conducted in populations receiving mixed surgeries (n=9), two studies included orthopaedic surgeries, one study included vascular surgeries, and one study included urology surgeries. Findings by surgery type can be seen in Table 10.

**Table 10.**
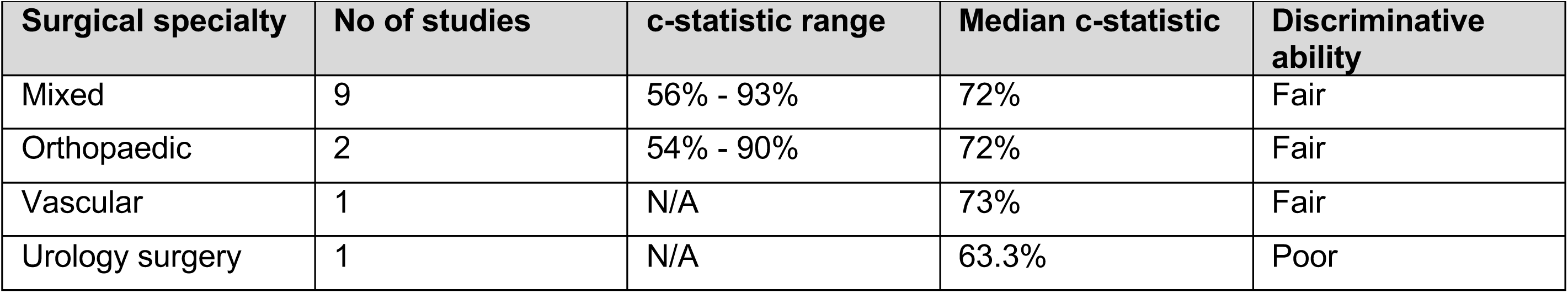
RCRI c-statistics by surgical specialty.

| Surgical specialty | No of studies | c-statistic range | Median c-statistic | Discriminative ability |
| --- | --- | --- | --- | --- |
| Mixed | 9 | 56% - 93% | 72% | Fair |
| Orthopaedic | 2 | 54% - 90% | 72% | Fair |
| Vascular | 1 | N/A | 73% | Fair |
| Urology surgery | 1 | N/A | 63.3% | Poor |

**Table 11.** RCRI calibration findings by surgical specialty.

| Surgical specialty | No of studies | O/E ratio range | Median O/E ratio |
| --- | --- | --- | --- |
| Vascular surgery | 1 | N/A | 2.23 |

#### 3.4.2 RCRI Calibration findings

Only one study assessing the RCRI reported O/E ratios (Moses et al., 2019). When looking at the RCRI’S ability to predict adverse cardiac events in vascular surgeries, **the O/E ratio was 2.23.**

#### 3.4.3 RCRI Accuracy findings

None of the 15 studies assessing the RCRI reported a Brier score.

#### 3.4.4 RCRI Healthcare utilisation and recovery outcomes

A total of two of the 16 studies assessing the predictive ability of RCRI reported on two healthcare utilisation and recovery outcome (readmission and reoperation) after orthopaedic surgery (Bronheim et al., 2018) or mixed surgeries (Schmidt et al., 2024). Only c-statistics were reported. The RCRI was found to have a poor discriminative ability for readmission (45.7% to 83.5%, with a median c-statistic of 64.6%) in two studies and good discriminative ability for reoperation in one study (c-statistic 85%). However as only one or two studies reported these outcomes, further evidence would be needed to draw firm conclusions.

#### 3.4.5 Bottom line summary for RCRI

When looking at composite complications, there is evidence to suggest the RCRI had a fair discriminative ability across 13 studies (median c-statistic of 72%). When looking at the discriminative ability of the RCRI by surgical specialty the c-stats varied, ranging from poor in one study after urology surgery to fair after mixed (n=9), orthopaedic (n=2) and vascular surgery (n=1). However, the findings for the individual surgical specialties are limited and showed wide ranges across studies, reducing the confidence in the findings. Calibration findings suggest the RCRI under predicted complications in the one study that reported this outcome (O/E ratio of 2.23). None of the studies assessing RCRI reported accuracy (Brier) scores.

The findings for the RCRI’s ability to predict healthcare utilisation and recovery outcomes was also very limited, suggesting further evidence is needed. However, there is evidence to suggest the RCRI had a poor discriminative ability for predicting readmission and a good discriminative ability for predicting reoperation.

The evidence directly comparing the predictive ability of the RCRI to the ASA classification system or to ACS NSQIP was limited and the findings appear to be mixed, suggesting further evidence is needed.

### 3.5 ASA classification system findings

#### 3.5.1 ASA classification system Discrimination findings

A total of nine studies reported c-statistics on composite complications for the ASA classification system (See Figure 8). One study reported c-statistics for the predictive ability of the ASA classification system for cardiac complications and pulmonary complications (Chrisant et al., 2024). One study reported c-statistics for the predictive ability of the ASA classification system for a surgical cohort and an endovascular cohort (Feghali et al., 2022), and one study reported c-statistics for patients receiving total hip arthroplasty, and total knee arthroplasty (McConaghy et al., 2021). **C-statistics ranged from 51.4%-77% with a median c-statistic of 60.8% suggesting the ASA had a poor predictive ability for composite complications across all nine studies.**

**Figure 8.**
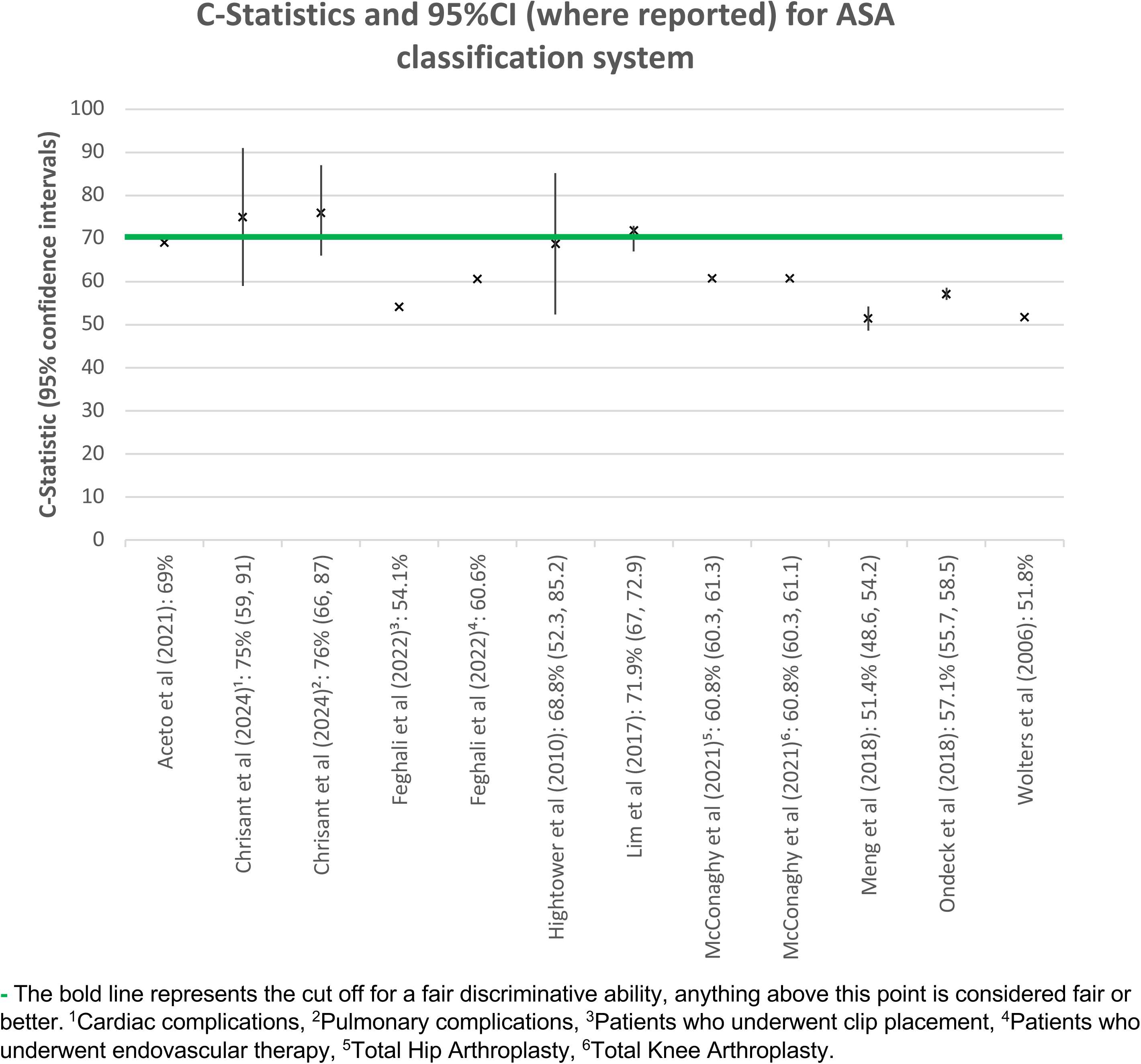
ASA classification system C-Statistics plot.

**Figure 9.**
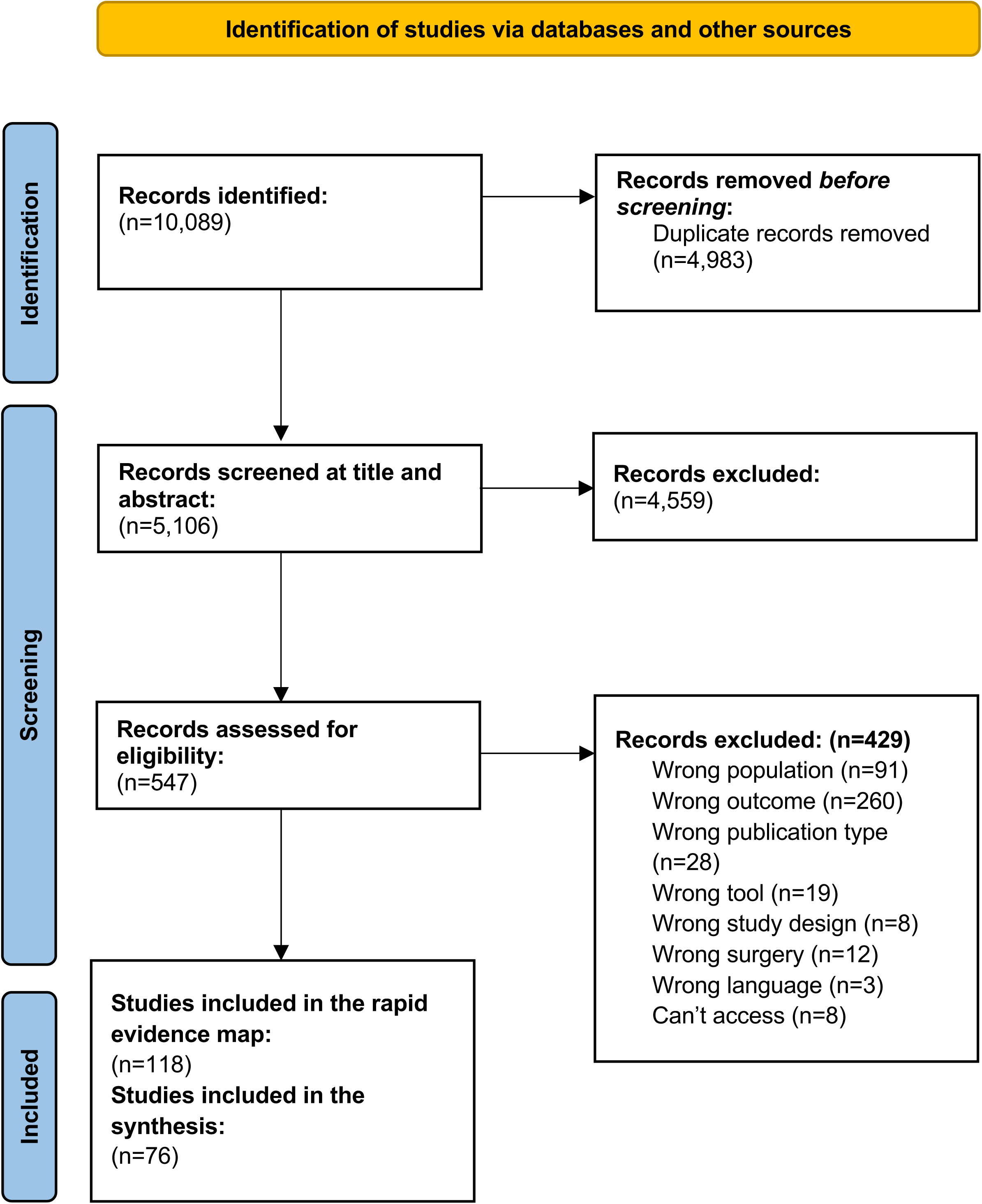
PRISMA Flow Diagram.

The majority of studies were conducted in populations receiving orthopaedic surgeries (n=3), two studies included mixed surgeries, two studies included vascular surgeries, one study included general surgeries, and one study included urology surgeries. The findings by surgery type can be seen in Table 12.

**Table 12.** ASA classification system c-statistics by surgical specialty.

| Surgical specialty | No of studies | c-statistic range | Median c-statistic | Discriminative ability |
| --- | --- | --- | --- | --- |
| Orthopaedic surgery | 3 | 57.1% - 71.9% | 60.8% | Poor |
| Mixed | 2 | 69% - 76% | 75% | Fair |
| Vascular surgery | 2 | 51.8% - 60.6% | 54.1% | Very poor |
| Urology surgery | 1 | N/A | 51.4% | Very poor |
| General | 1 | N/A | 68.8% | Poor |

#### 3.5.2 ASA classification system Calibration findings

None of the 13 studies assessing the ASA classification system reported O/E ratios.

#### 3.5.3 ASA classification system Accuracy findings

None of the 13 studies assessing the ASA classification system reported a Brier score.

#### 3.5.4. ASA classification system Healthcare utilisation and recovery outcomes

A total of five of the 13 studies assessing the predictive ability of the ASA classification system reported on four healthcare utilisation and recovery outcomes. This included readmission, reoperation, length of stay, extended length of stay, and discharge not to home. The studies reporting these outcomes only included orthopaedic surgeries (n=4) or urology surgeries (n=1). Only c-statistics were reported and can be seen in Appendix 3. It is important to note the definition of extended length of stay varied slightly across the studies. Extended length of stay was defined as a hospital stay greater than the 75th percentile, or greater than 3 days (Fu et al., 2018), a hospital stay greater than one day (McConaghy et al., 2021), a hospital stay greater than the 75th percentile (Meng et al., 2018) or a hospital stay greater than or equal to the 75th percentile (Ondeck et al., 2018).

While only a limited number of studies reported each outcome, the evidence suggests that overall, the ASA classification system has a very poor discriminative ability for length of stay (c-statistics of 53.6% from 1 study), and extended length of stay (ranging from 56.1%-63%, with a median c-statistic of 57.2% across three studies). A poor discriminative ability for predicting discharge not to home (ranging from 59%-64%, with a median c-statistic of 63.1% across 4 studies), and an excellent discriminative ability for predicting readmission (90.6% from 1 study) and reoperation (86.6% from one study). However, as each outcome was only reported by a limited number of studies further evidence is needed. Overall, the evidence for the predictive ability of the ASA classification system to assess healthcare utilisation and recovery outcomes was very mixed.

#### 3.5.5. Bottom line summary for ASA classification system

When looking at composite complications, there is evidence to suggest the ASA classification system had a poor discriminative ability across nine studies (median c-statistic of 60.8%). When looking at the discriminative ability of the ASA classification system by surgical specialty the c-stats varied, ranging from very poor after vascular (n=2) or urology surgery (n=1) to fair after mixed surgeries (n=2). However, the findings for the individual surgical specialties are limited and showed wide ranges across studies, reducing the confidence in the findings. None of the studies assessing the ASA classification system reported calibration (O/E ratio) or accuracy (Brier) scores.

The findings for the ASA classification system’s ability to predict healthcare utilisation and recovery outcomes was also very limited, suggesting further evidence is needed. However, overall, there is evidence to suggest the ASA classification system had a very poor discriminative ability for length of stay, or extended length of stay, a poor discriminative ability for predicting discharge not to home, and an excellent discriminative ability to predict readmission or reoperation.

The evidence directly comparing the predictive ability of the ASA classification system compared to the RCRI was limited and the findings appear to be mixed suggesting further evidence is needed.

## 4. DISCUSSION

### 4.1 The evidence base

This review set out to identify and map the evidence for externally validated pre-operative surgical risk prediction tools currently used in Wales for identifying patients suitable for surgery in low-risk regional surgical settings, such as surgical hubs. Accurate risk prediction is particularly valuable in these settings as they help clinicians to identify which patients can safely benefit from treatment while maintaining the efficiency and safety standards required for such facilities. This work builds on the NICE (2020) evidence review exploring which of the P-POSSUM, SORT, or ACS NSQIP risk stratification tools could best identify the risk of mortality and morbidity of adults undergoing surgery by examining more risk stratification tools. However this review focused on post-surgical complications and healthcare utilisation and recovery outcomes, which are likely to be priority considerations in low-risk regional surgical settings, such as surgical hubs.

A total of 118 studies were included in the rapid evidence map across 12 risk prediction tools. The most commonly studied risk prediction tool was ACS NSQIP (n=40), with the least common being the Carlisle Risk Calculator and the National Emergency Laparotomy Audit Parsimonious Risk Score (NELA PRS) where no relevant evidence was identified. Of the three risk prediction tools that were also included in the NICE evidence review (NICE, 2020), our evidence map identified a total of 51 studies meeting our eligibility criteria. Although not examined, it is likely some of these studies are included in both reviews.

The tools have been developed and used for a wide variety of surgical types, with some not specifically developed for surgery (NRS-2002 and CFS). All but one tool was designed to be carried out by clinicians. The exception to this is the DASI tool, which is a self-administered questionnaire, completed by patients. The SORT risk prediction tool and the ACS NSQIP tool is designed to be used in conjunction with the ASA (or a modified version) classification system, as it is included as part of the risk calculation. The ASA classification risk prediction tool should be considered alongside evaluations to determine appropriateness of surgery and should not be used alone. The ARISCAT score, CPET, POSSUM, P-POSSUM and RCRI tools utilise clinical findings in their assessment of pre-surgical risk while the others assess risk based on lifestyle questions or exercise tests. A further challenge relates to differences in assessed outcomes. Some tools predict specific complications (e.g., postoperative nausea and vomiting), while others estimate overall morbidity or surgical fitness (e.g., ASA classification system, CPET). Varying risk definitions further complicate direct comparisons. While a large evidence base exists for these surgical risk prediction tools, given the variability in terms of surgery and outcomes assessed, it is unlikely that any one tool would be suitable for use across all populations and surgery types within a surgical hub setting.

### 4.2 Findings for ACS NSQIP, P-POSSUM, RCRI and the ASA classification system

This review also set out to provide a more in-depth summary of the findings for a selection of the tools deemed to be the most applicable on a population level. When looking at the findings for ACS NSQIP, P-POSSUM, the RCRI and the ASA classification system in more detail, the performance results appeared to be mixed. There was considerable heterogeneity amongst the included studies which limits direct comparisons and reduces the confidence in the overall findings. The evidence available for each tool shows a variation in how well the tool performed, while ACS NSQIP, P-POSSUM and the ASA classification system were all found to have an overall poor predictive ability across all studies, the RCRI was found to have a fair predictive ability across studies. This could suggest that the RCRI was the most effective at predicting complications across a range of surgical disciplines, however, the tools had been assessed in different surgical specialties, with ACS NSQIP being assessed across the largest number of surgical specialities (n=9), followed by the ASA classification system (n=5), the RCRI (n=4) and P-POSSUM (n=3) which could skew the findings if the tools are found to be more effective for predicting morbidity in certain surgical specialties than others. The evidence supports this as the findings for each tool show a considerable range in predictive ability depending on surgical specialty, which suggests the overall scores may not adequately reflect the predictive ability of each tool. P-POSSUM and the ASA classification system were found to range from very poor to fair when the findings were split by surgical specialty, the RCRI was found to range from poor to fair depending on the surgical specialty and ACS NSQIP was the only risk prediction tool that was found to range from very poor (orthopaedic, urology and vascular surgery) to excellent (mixed surgery) depending on the surgical specialty. While the evidence suggests that certain tools may be more effective for specific surgical specialties or even specific outcomes, the evidence base available for each surgical specialty varied between tools and was often very limited. As such, further evidence would be needed to draw firm conclusions, and the results should be interpreted with caution.

### 4.3 Limitations of the available evidence

The evidence highlights a large variation in what the risk prediction tools are predicting within the literature which makes synthesis of included studies very challenging. Some studies provided composite outcomes, incorporating multiple single outcomes, and some studies reported findings for individual outcomes. The studies reporting composite outcomes provide a larger range of outcomes which may make the findings more generalisable. However, the findings may be skewed if a risk prediction tool is found to be extremely good or extremely bad at predicting one of the outcomes included in the composite. For example, ‘cardiac complications may include both ‘cardiac arrest requiring CPR’ and ‘myocardial infarction’; the risk prediction tool’s performance may vary between the two individual outcomes which make up the composite. Some composite complications were also less frequently reported in the literature, such as ‘pulmonary complications’. The studies that reported individual outcomes highlight which specific outcomes the risk prediction tools may be better suited at identifying. Thus, the limitation is that the evidence base may be limited by a smaller number of studies reporting findings for each outcome, which will reduce the overall certainty of the findings.

A limitation of the evidence is the variance in the statistical reporting of outcome results. Most studies reported the discriminative ability of the risk prediction tool using the ‘area under the curve’ (AUC) and a combination of discrimination and calibration using the ‘Brier score’. However, some studies only reported calibration findings, and a smaller number of studies reported only the Brier score. For this reason, comparison of predictive ability across tools was challenging. In addition, although the Brier score is considered the most comprehensive score of accuracy, Brier scores should not be directly compared across studies using different patient data, as the Brier score depends on both the prevalence of the event in the data and the performance of the tool. Thus, overall comparison of a tool’s Brier score was not possible, although Brier scores for individual studies have been reported.

Some tools were well represented in the literature by a larger number of studies, such as ACS NSQIP (n=40), whereas other tools such as P-POSSUM had a smaller number of studies (n=7). Broader surgery types, such as ‘general’ (n=43) and ‘mixed surgeries’ (n=23) were more frequently represented in the literature than other surgery types, again reducing the overall certainty of the findings for the tools with fewer studies and in a less common surgical category. Most studies were conducted in the USA which may limit the generalisability of the findings to the Welsh context.

### 4.4 Summary of the Evidence gaps

While overall a large evidence base was identified, a number of risk prediction tools had very minimal evidence available with the NRS-2002, ARISCAT, CFS, SORT and DASI all being assessed in five studies or less. No evidence was identified assessing the predictive ability of the Carlisle Risk Calculator or the NELA PRS. In addition, no evidence was identified looking at the use of risk prediction tools when identifying patients suitable for treatment in surgical hubs.

### 4.5 Strengths and limitations of this Evidence Review

The studies included in this review were identified through an extensive search of electronic databases, trial registries, grey literature sites and through citation tracking. As the search identified a very large number of studies, the review can be considered a good reflection of the overall evidence base for this topic. However, there is a possibility that additional publications may have been missed or we may have introduced some biases to this review. As no date or country limits were set, it is possible that some of the evidence gained could be outdated (for example if the tool had been updated overtime or if surgical practices have changed, making them safer), and as a wide range of countries were included it is unclear how generalisable the findings would be to the UK context. No quality appraisal of included studies was conducted and therefore we cannot report the quality of the included studies.

Although it has been impossible to directly compare some accuracy measures (such as the Brier score) across the same risk prediction tool, the in-depth summary does compare the discriminative ability and the calibration of the four tools of interest using the AUC and the O/E ratios (where reported), producing median values which give an indication of the overall strength of the tool’s performance. Given the complexity of the evidence base, the map is organised into aspects which would be most useful to clinicians looking to choose a tool for use within a low-risk surgical setting, looking at outcomes of composite complications and overall morbidity.

While the review is focussed on evidence relating solely to morbidity outcomes, we have also included separately evidence in which mortality is included within a composite morbidity outcome, to recognise the frequency in which mortality is included in these composite outcomes (Appendix 1).

In addition, the review excluded studies assessing the external validation of modification of the 13 risk predictions tools of interest. It is common for risk prediction tools to undergo modification in order to make them better at predicting risk in specific situations or for specific groups. However, those identified during the screening process have been collated and referenced in Appendix 2 in order to better reflect the totality of the evidence base.

### 4.6 Implications and next steps

This review provides a summary of 12 risk prediction tools currently used in surgical disciplines across Wales. It can be used to inform practice in low-risk surgical settings in Wales to help clinicians, practitioners and other stakeholders in deciding which tools are most appropriate to use for different surgery types by giving an indication of the predictive ability of four discrete tools: ACS NSQIP, ASA, RCRI and P-POSSUM for composite and morbidity outcomes. However, several limitations were identified, such as inconsistent reporting methods and heterogeneity across the studies, and the variation in the amount of evidence available for each tool. In addition, no quality appraisal of the included studies was conducted and as such the findings should be interpreted with caution. While it is clear that no risk prediction tool adequately predicted complications across all surgical specialties, it may be likely that some tools are better suited for specific surgery types or that a combination of risk prediction tools may be needed to adequately assess an individual’s level of risk.

This review has identified evidence gaps as no external validation studies were retrieved for the Carlisle Risk Calculator and the National Emergency Laparotomy Audit Parsimonious Risk Score (NELA PRS), suggesting further research is needed for these tools. As considerable limitations were identified limiting the comparability between studies, further research should ensure risk prediction tools are assessed in a consistent way to allow for direct comparisons.

### 4.7 Economic considerations*

- Future research into risk prediction tools should incorporate health economic evaluations, to provide consideration of individual risk as well as associated health and social care resource use costs. NICE state that risk prediction tools are freely available, and therefore do not have an associated cost to use. However, they do require some time to complete, but this is typically less than 5 minutes during a preoperative assessment NICE (2020).
- Economic evaluation/impact evaluations of risk prediction tools are a known evidence gap, as described by a systematic review of Health Economic Impact Evaluations of Risk Prediction Models (van Giessen et al., 2017). This review considered any study where a clinical risk prediction model was evaluated by health economic evaluation. Further, an evidence review of preoperative risk stratification tools conducted by NICE also identified no relevant economic evaluations NICE (2020).
- Model-based health economic evaluations may be an appropriate method of conducting health economic analysis of risk prediction tools as it can account for long-term health and cost outcomes. Further, it can be applied across cohorts of different surgical specialties (van Giessen et al., 2017).

\**This section has been completed by the Centre for Health Economics & Medicines Evaluation (CHEME), Bangor University*

## 5. EVIDENCE REVIEW METHODS

### 5.1 Eligibility criteria

**Table 13.**
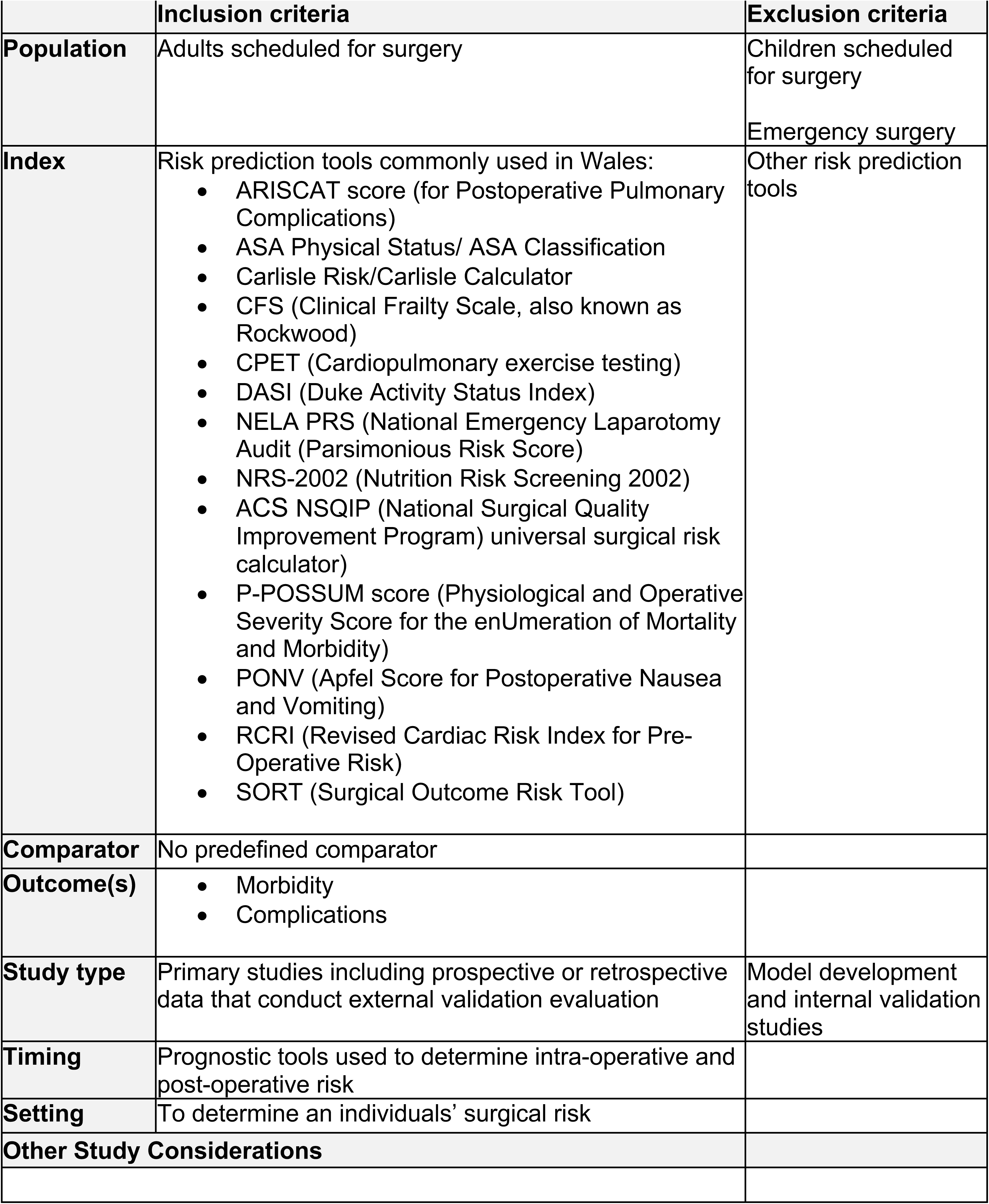
Eligibility criteria.

### 5.2 Literature search

All searches were conducted between 9^th^ Dec −17^th^ Dec 2024. The search strategies used for MEDLINE can be seen in Appendix 4. A search of the following databases and resources was conducted to identify published and ongoing primary studies:

- Medline
- Embase
- CINAHL
- Cochrane Central Register of Controlled Trials (CENTRAL)
- Clinicaltrials.gov
- WHO International Trials Registry Platform
- Scopus
- Google Scholar

Several websites and specialist sources were also searched, including:

- Royal College of Anaesthetists
- Royal College of Surgeons
- The Royal College of Surgeons of England
- NHS Scotland
- NHS England
- NIHR Public Health Research
- NICE
- The Health Foundation
- Centre for Peri-operative Care
- Association of Anaesthetists
- American Society of Anesthesiologists (ASA)
- GIRFT-Getting It Right First Time Home - Getting It Right First Time - GIRFT
- EPPI centre

### 5.3 Study selection process

The searches yielded a total of 10,089 records. Records were imported into an Endnote database library and duplicates were removed. After deduplication, a total of 5,106 records were screened at title and abstract. Title and abstracts were screened by one reviewer in Rayyan, with around 10% being assessed by another reviewer to ensure consistency and minimise bias. Any uncertainty was discussed within the review team. The full texts were screened by two reviewers in duplicate, if disagreements arose, these were discussed, and a third reviewer was consulted to make a final inclusion decision. After full text screening a total of 118 records met the inclusion criteria for the REM.

### 5.4 Data extraction and coding/charting

Data extraction was conducted by one reviewer and consistency checked by another reviewer while creating the evidence map and summaries for each risk prediction tool. Extracted information included the study’s country, study aim, design, population, surgery type, surgery category, sample size, outcomes of interest, and reported outcomes.

The evidence map employed a structured coding approach. Studies were first grouped by the surgical risk prediction tool assessed and then by surgical specialty, following Royal College of Surgeons categorisations. Studies using data from a combination of different surgical specialties were categorised as ‘Mixed’ and where categorisation was not possible or unclear, reviewers used information from the study to code the surgical specialty. Outcomes were coded into broad categories, including complications and healthcare utilisation and recovery. Complications encompassed any reported complications, while healthcare utilisation included measures such as length of stay, readmission, and discharge to higher-level care. Where specified, complications were further classified into subcategories such as cardiac, non-cardiac, and pulmonary complications to accurately represent a tools focus. Once all studies had been grouped and assigned into categories, they were charted onto the evidence map accordingly.

### 5.5 Assessment of methodological quality

None of the studies included in this review were assessed for methodological quality.

## 6. EVIDENCE

### 6.1 Search results and study selection

### 6.2 Data extraction Tables

**Table 14.**
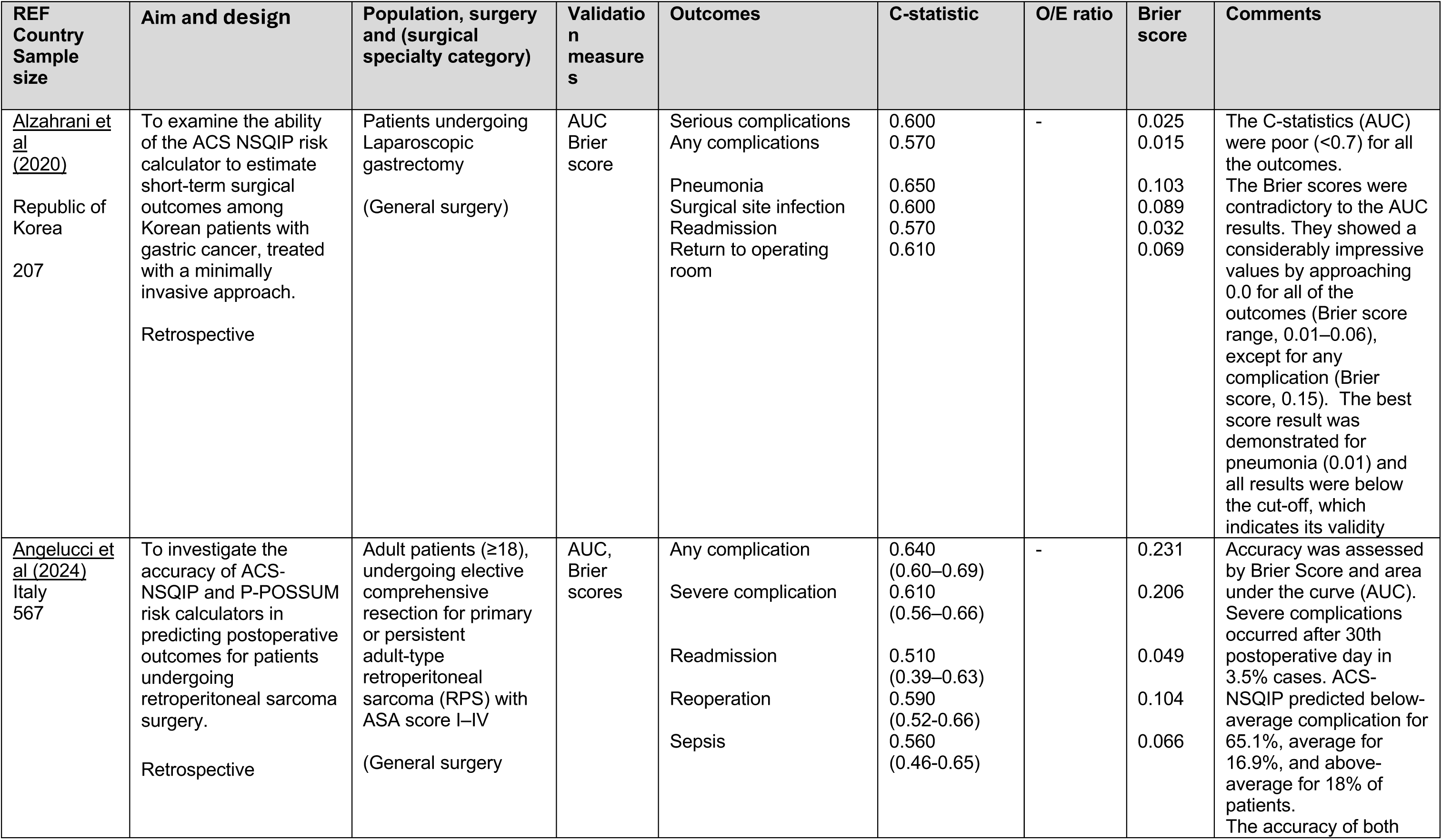
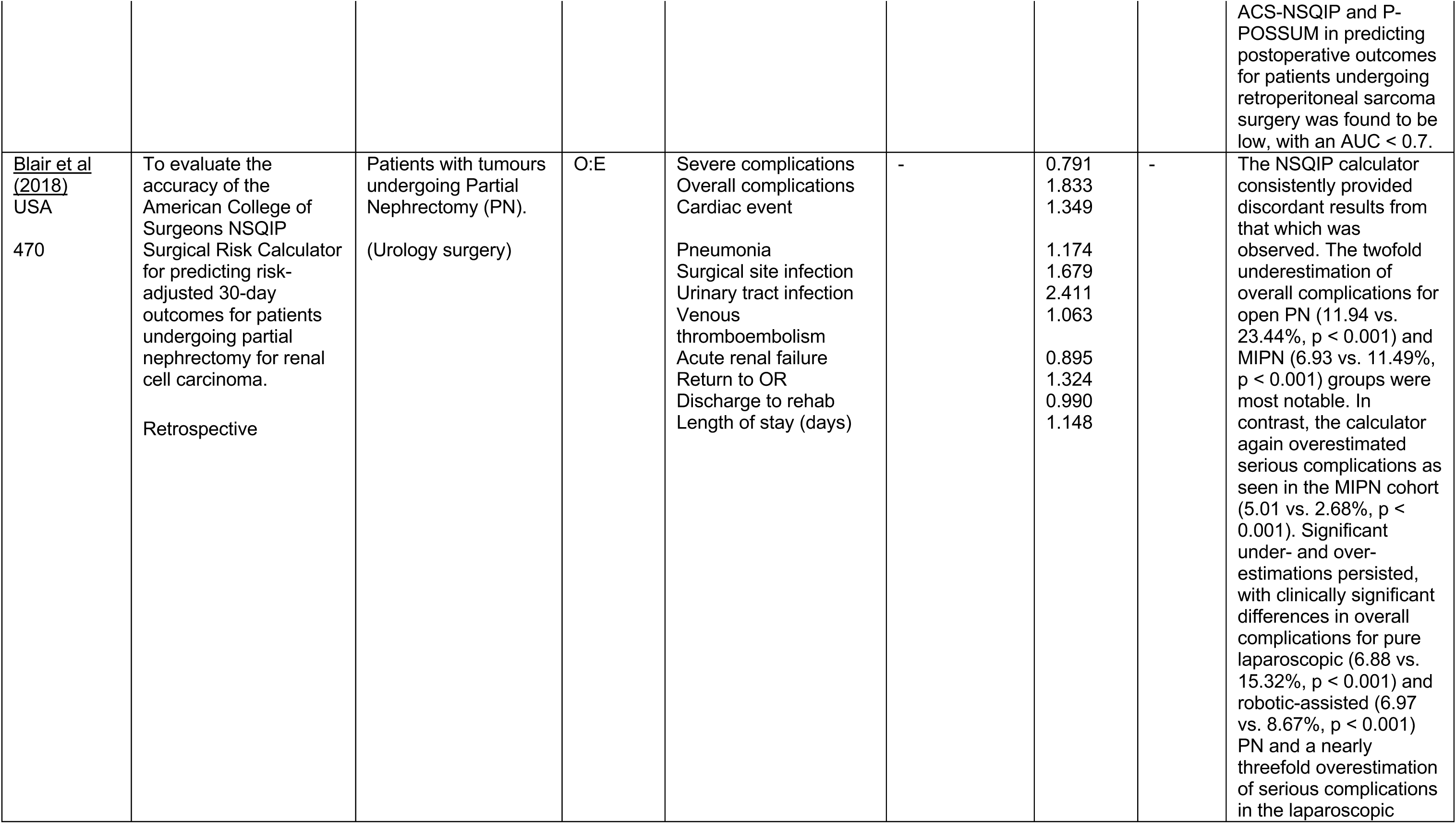
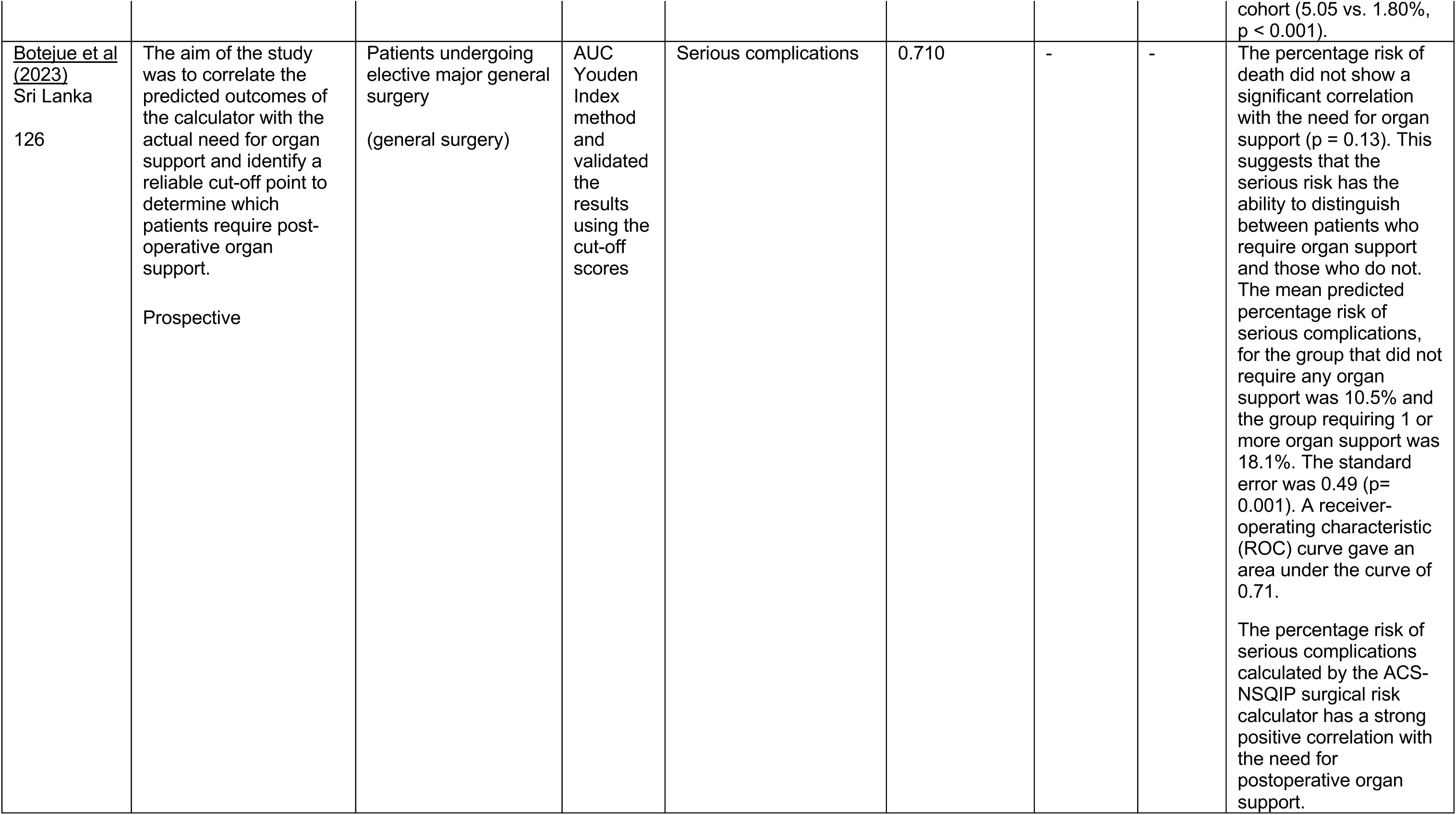
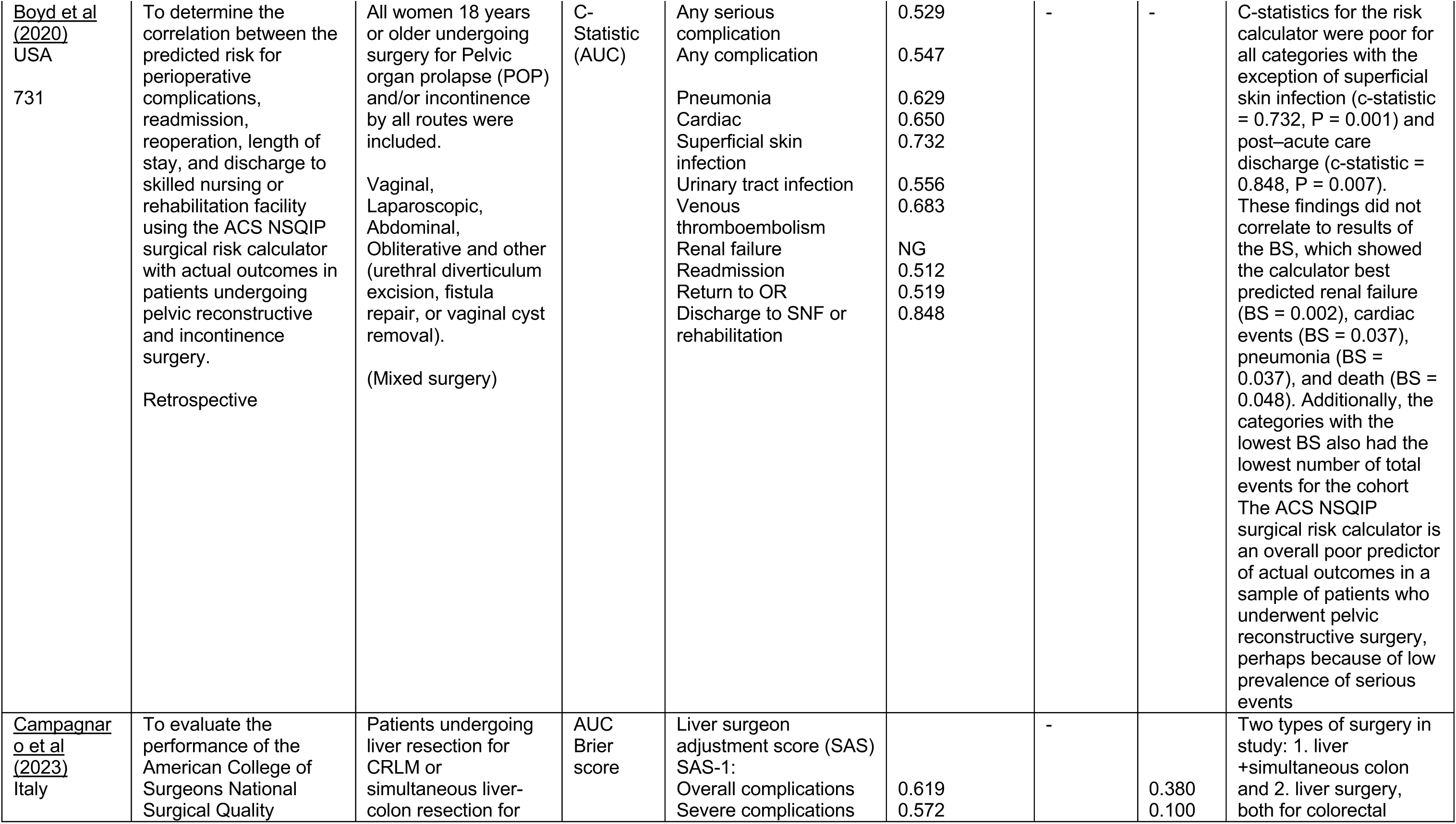
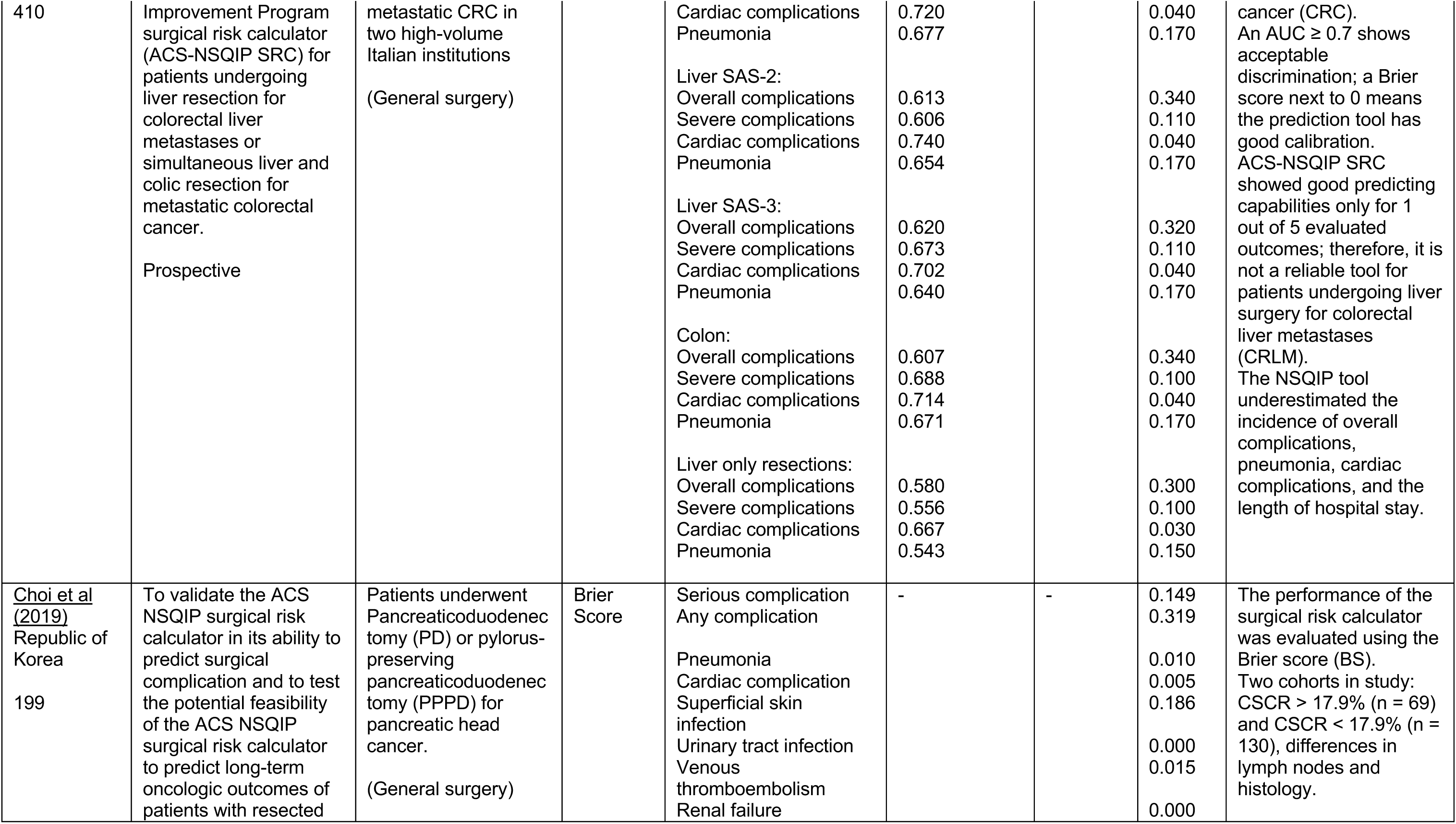
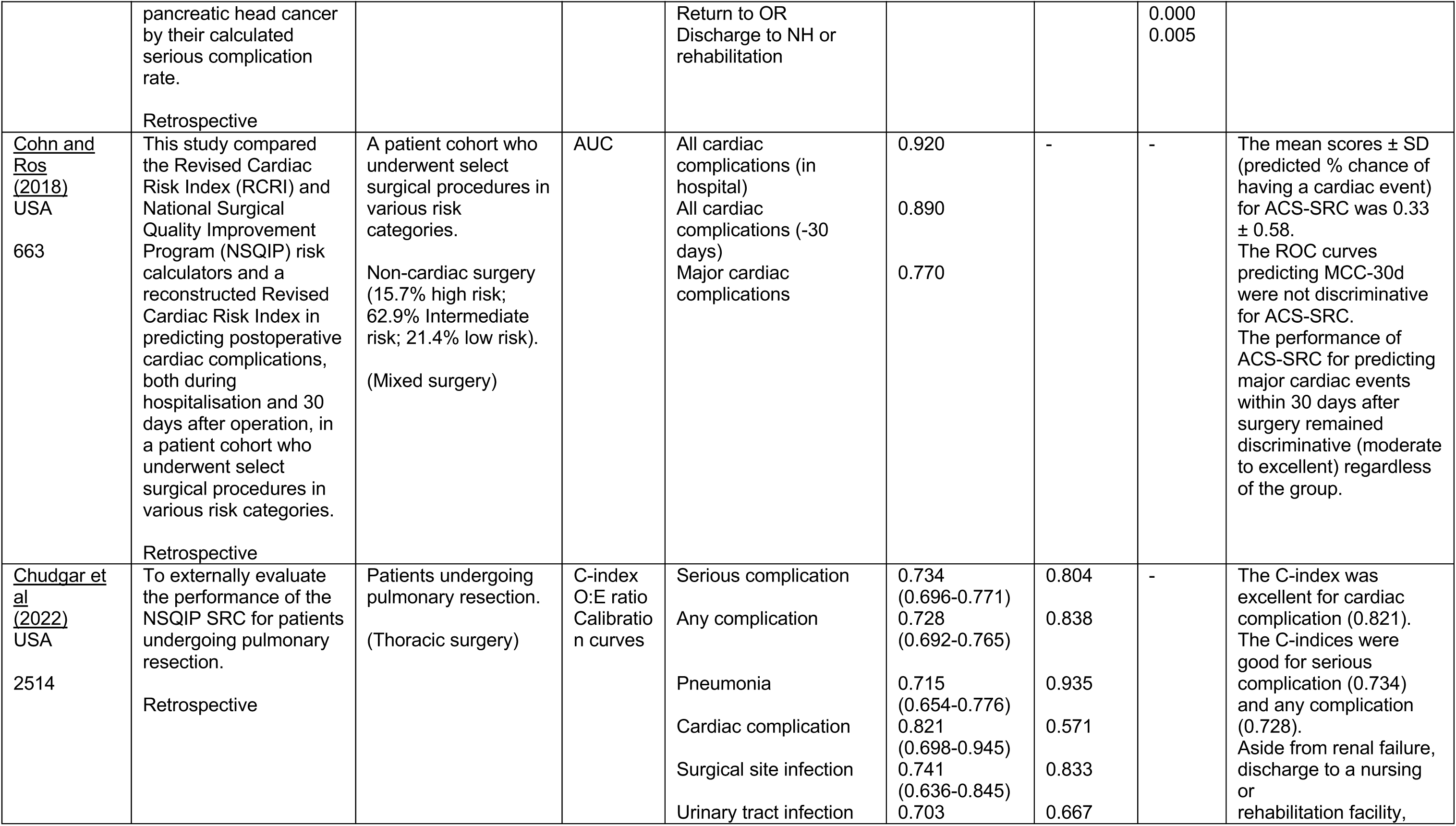
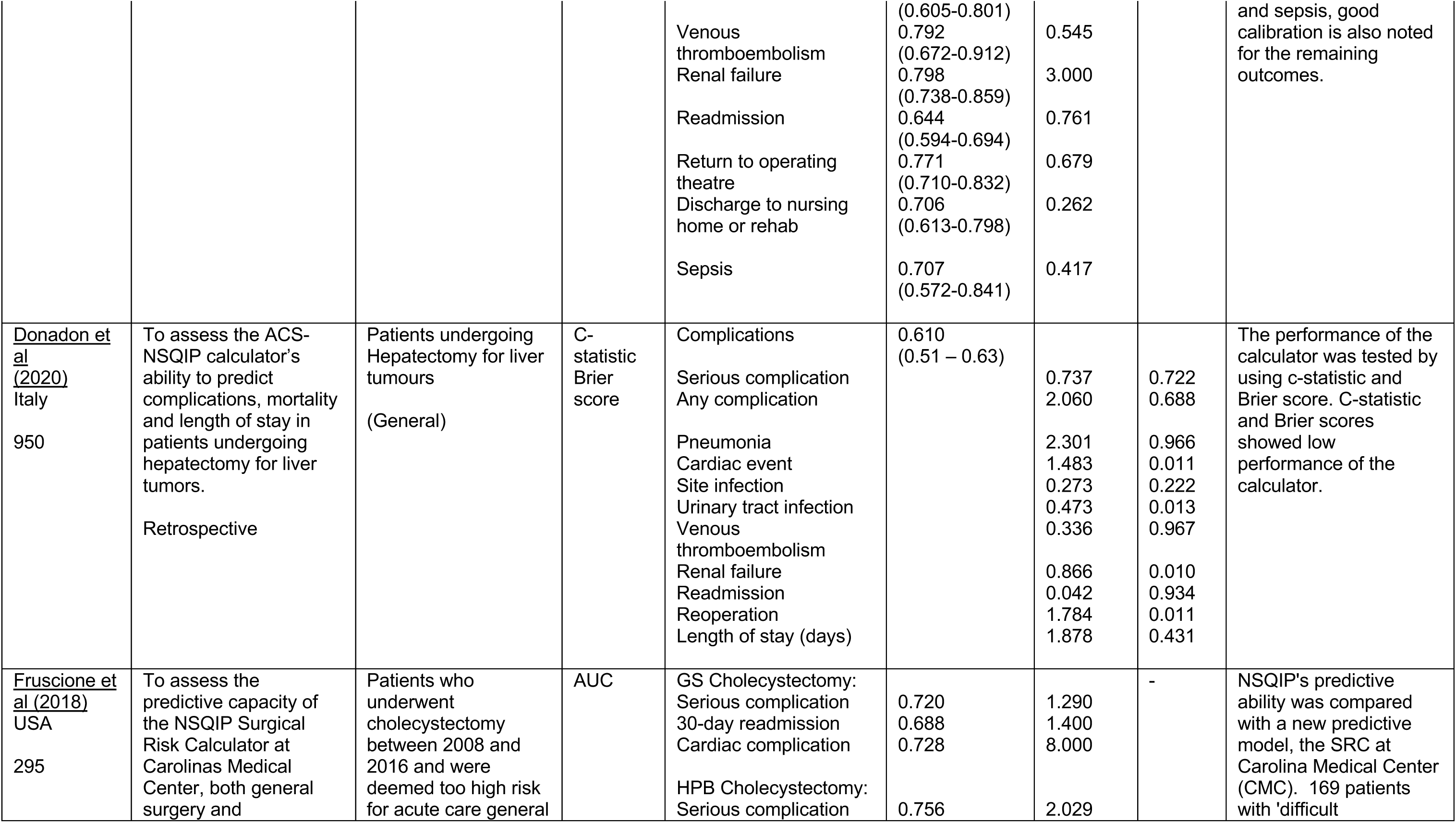
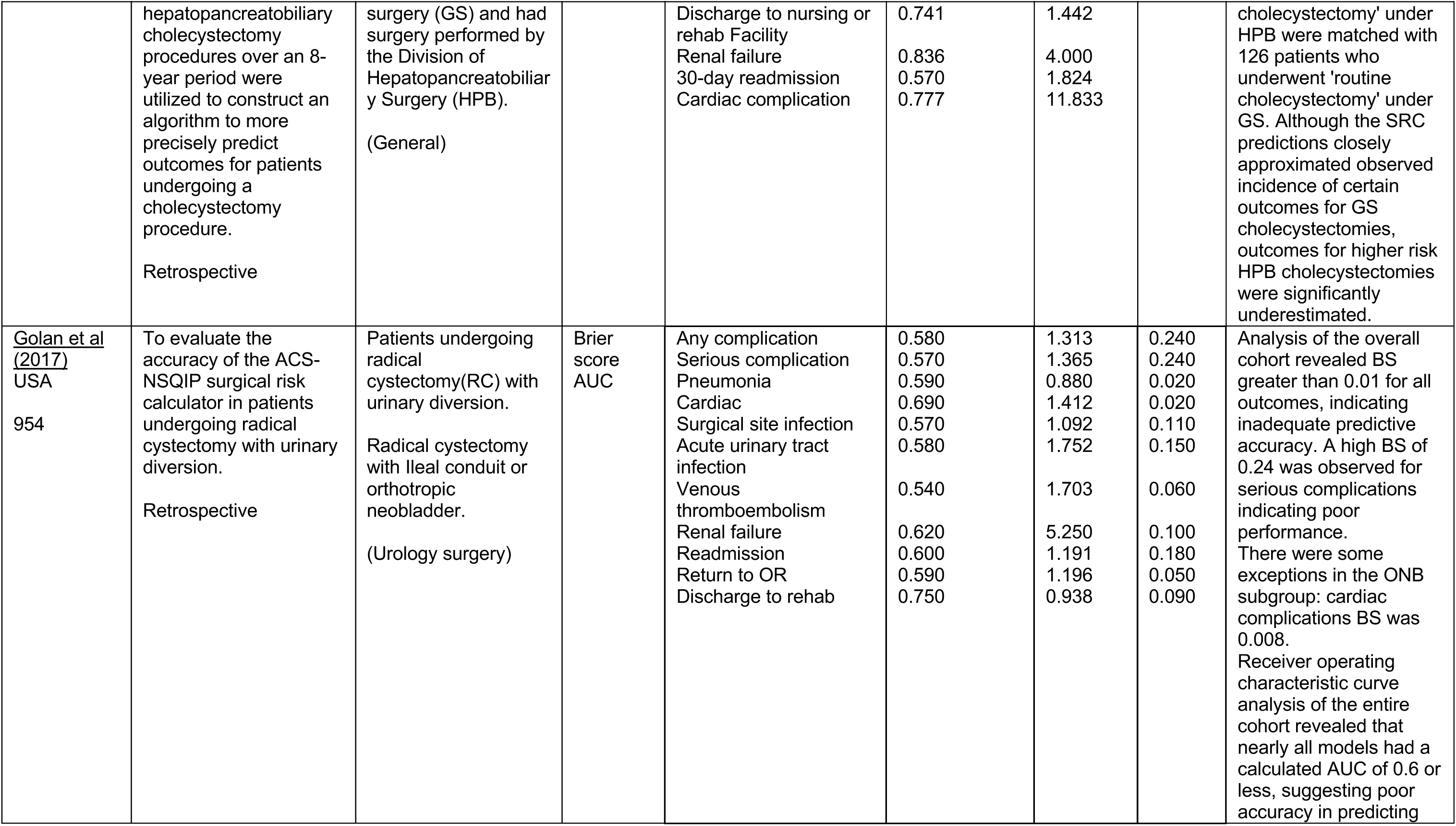
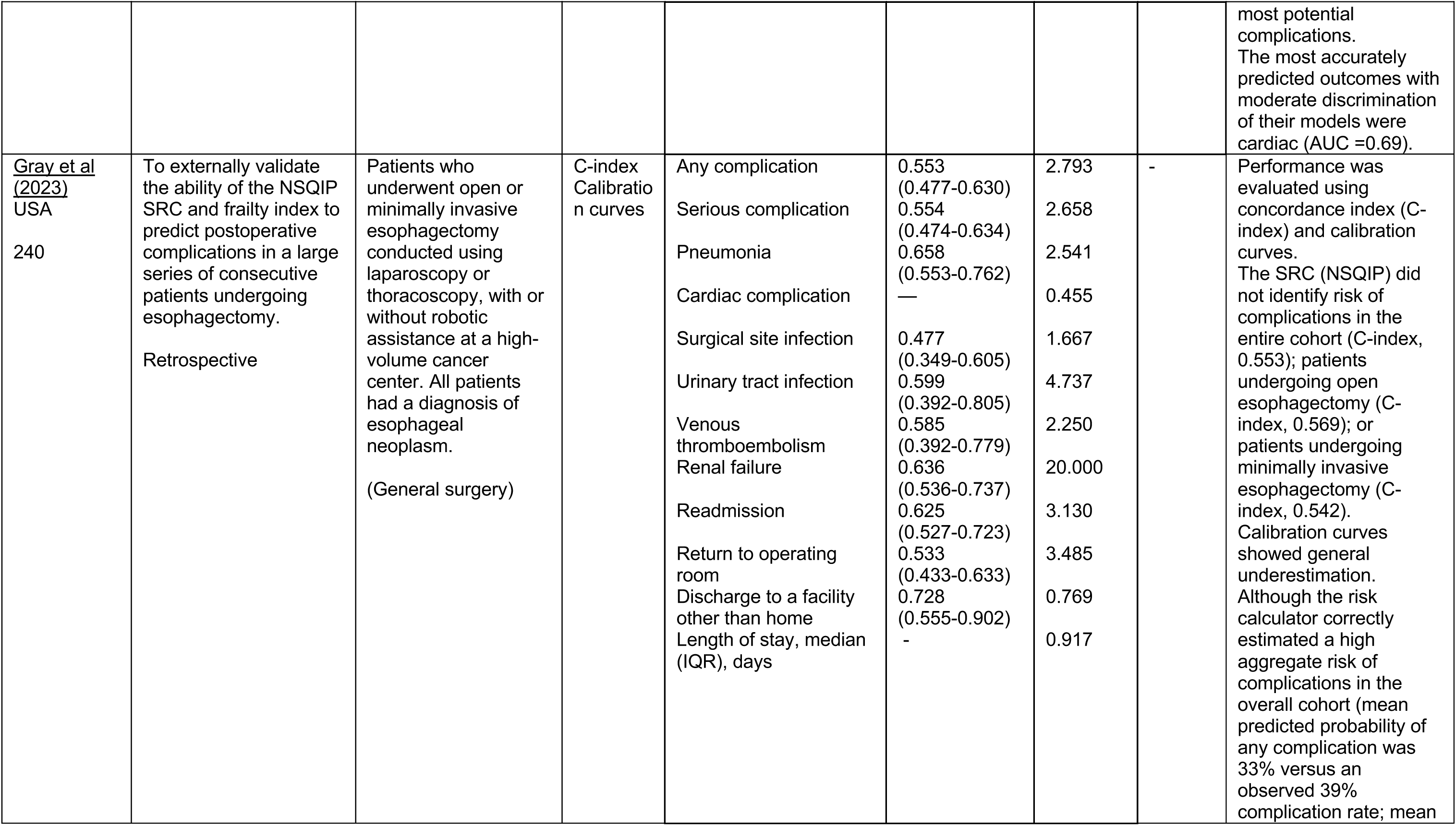
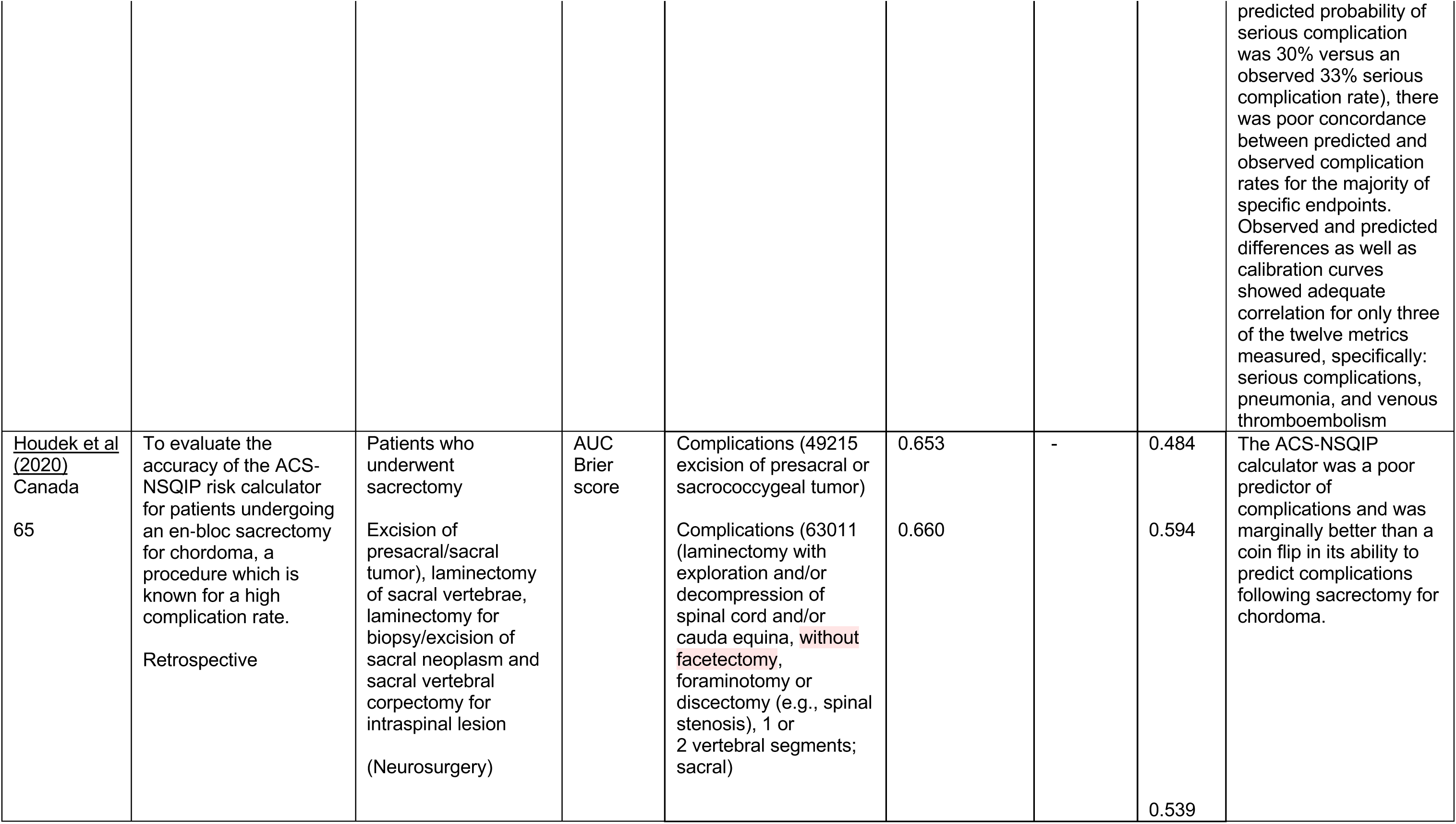
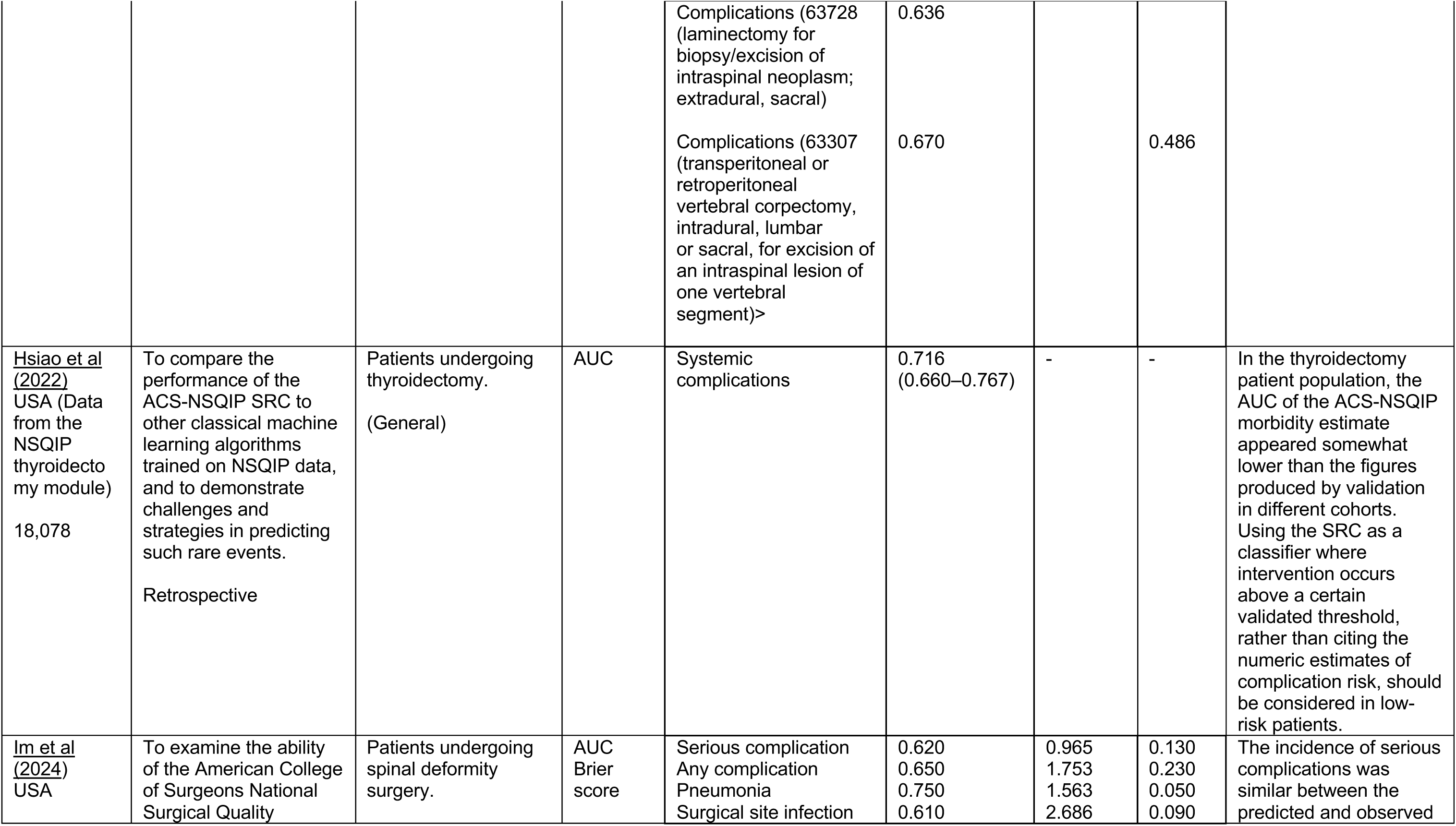
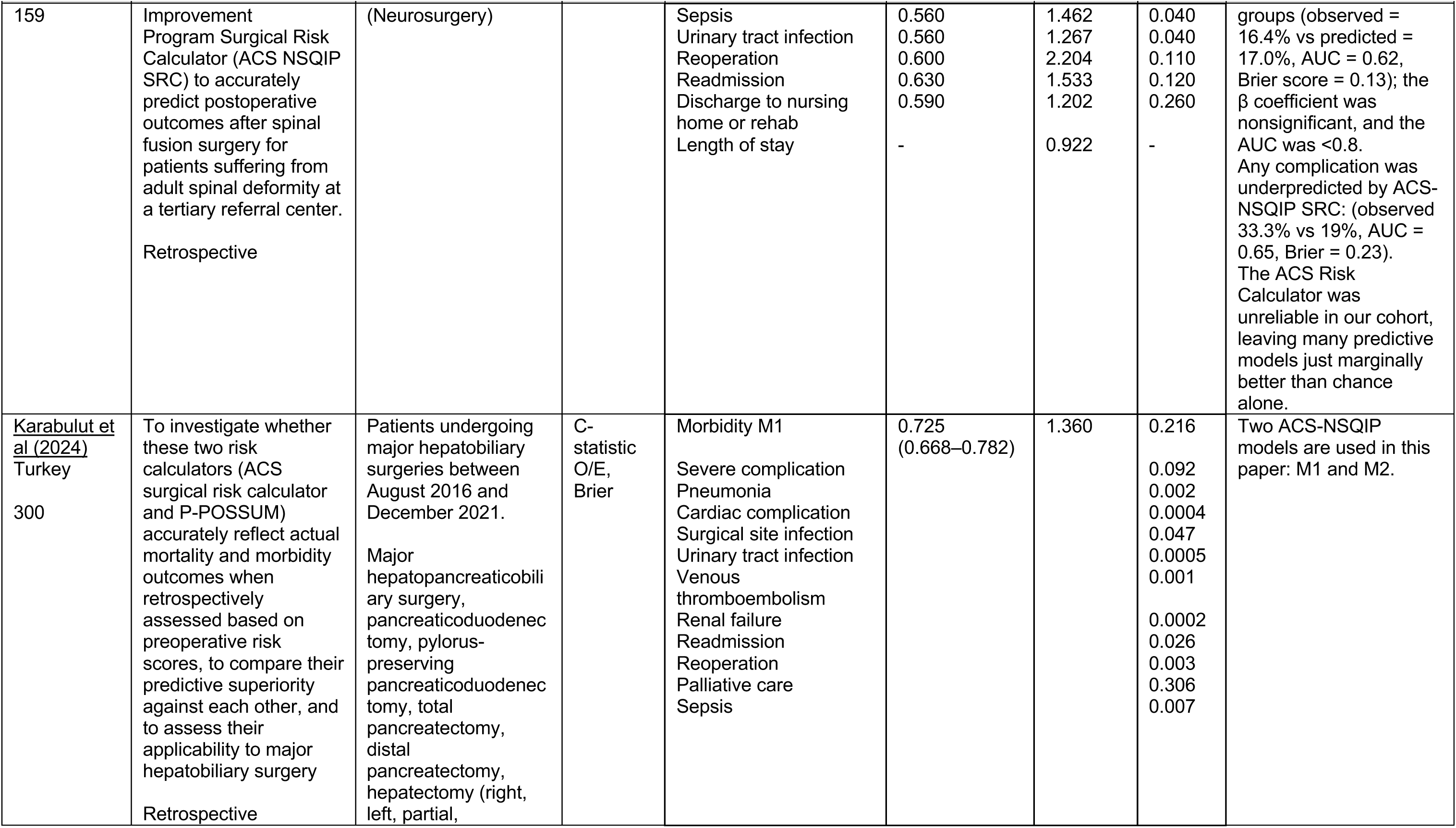
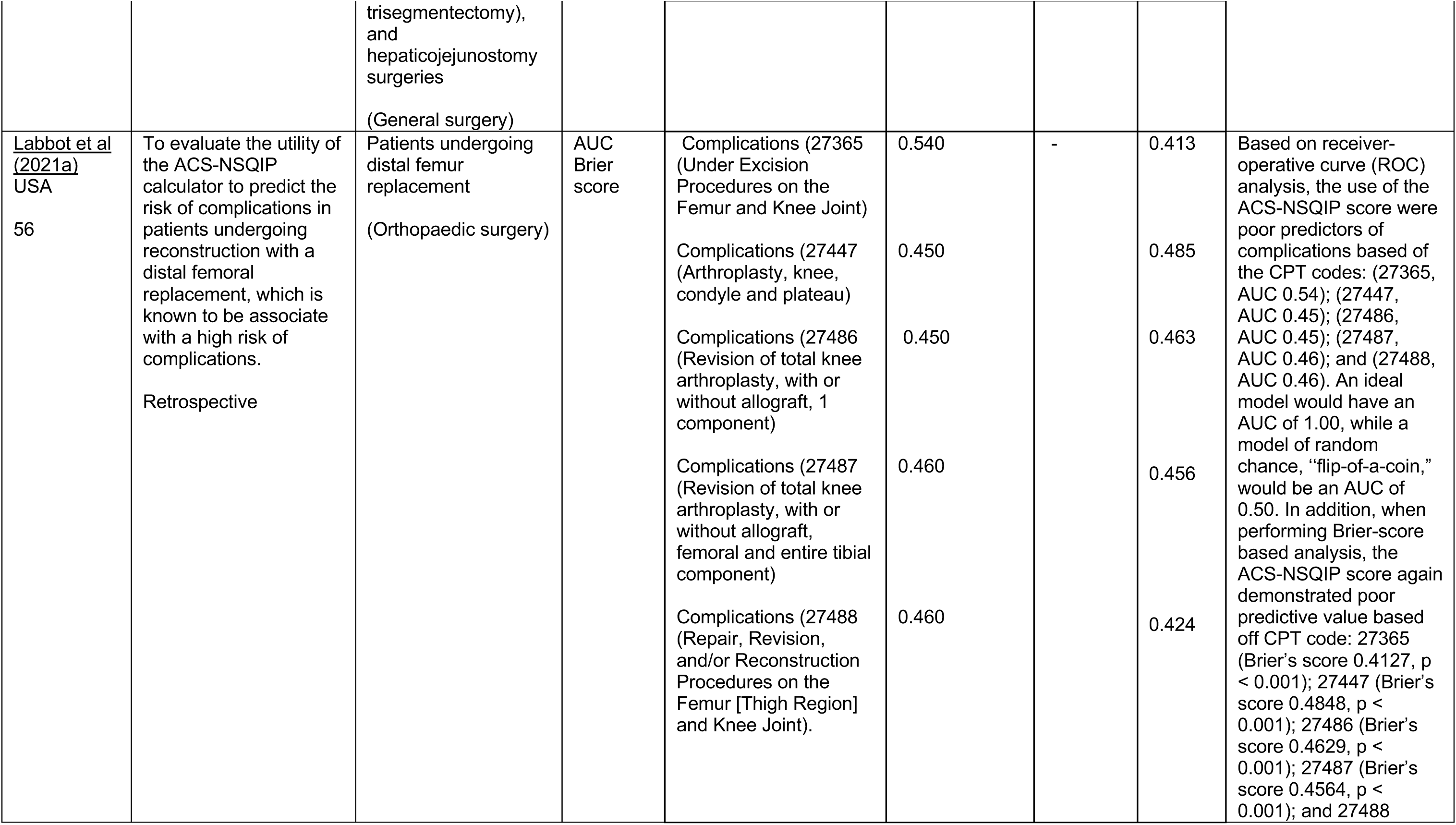
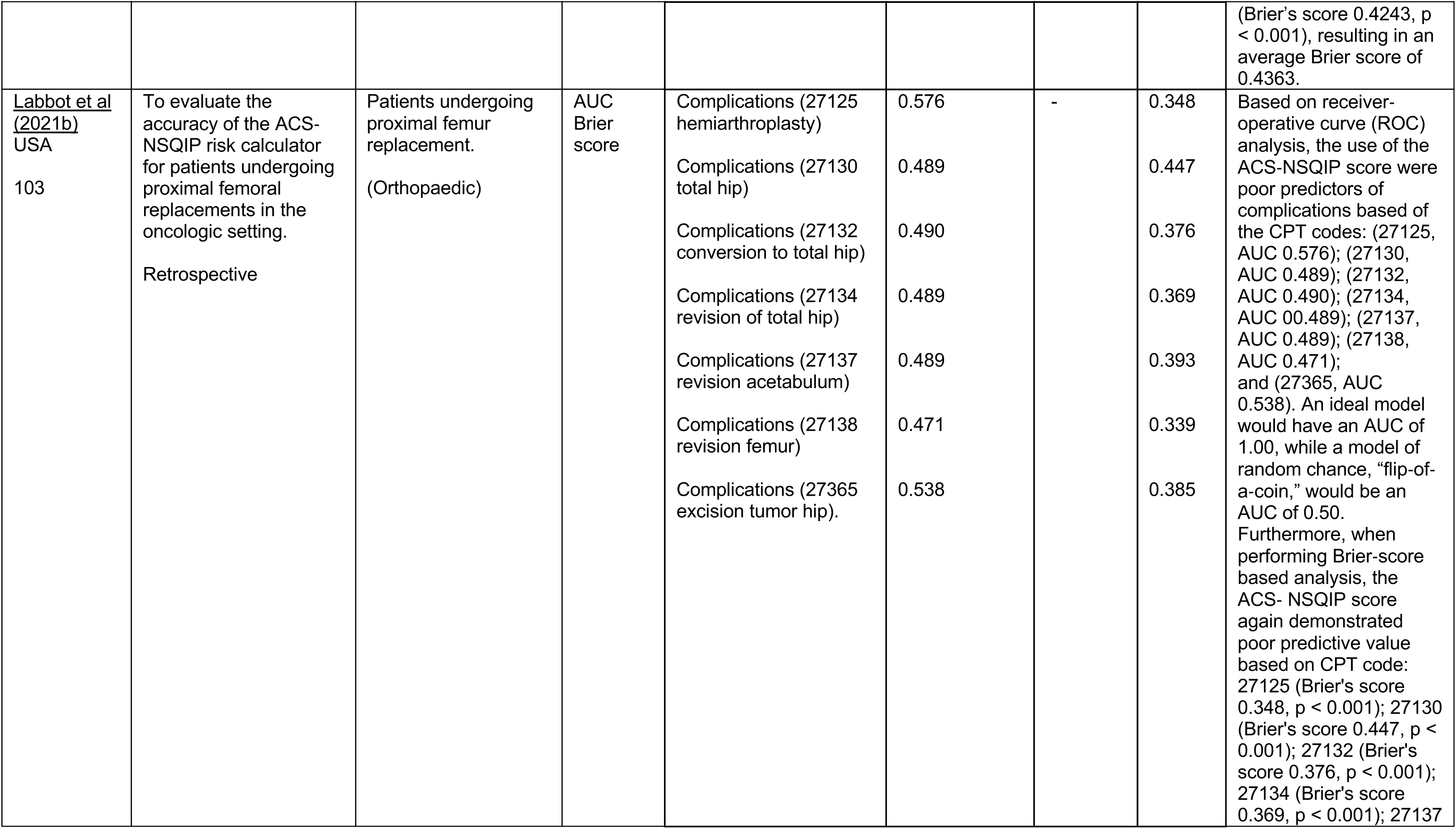
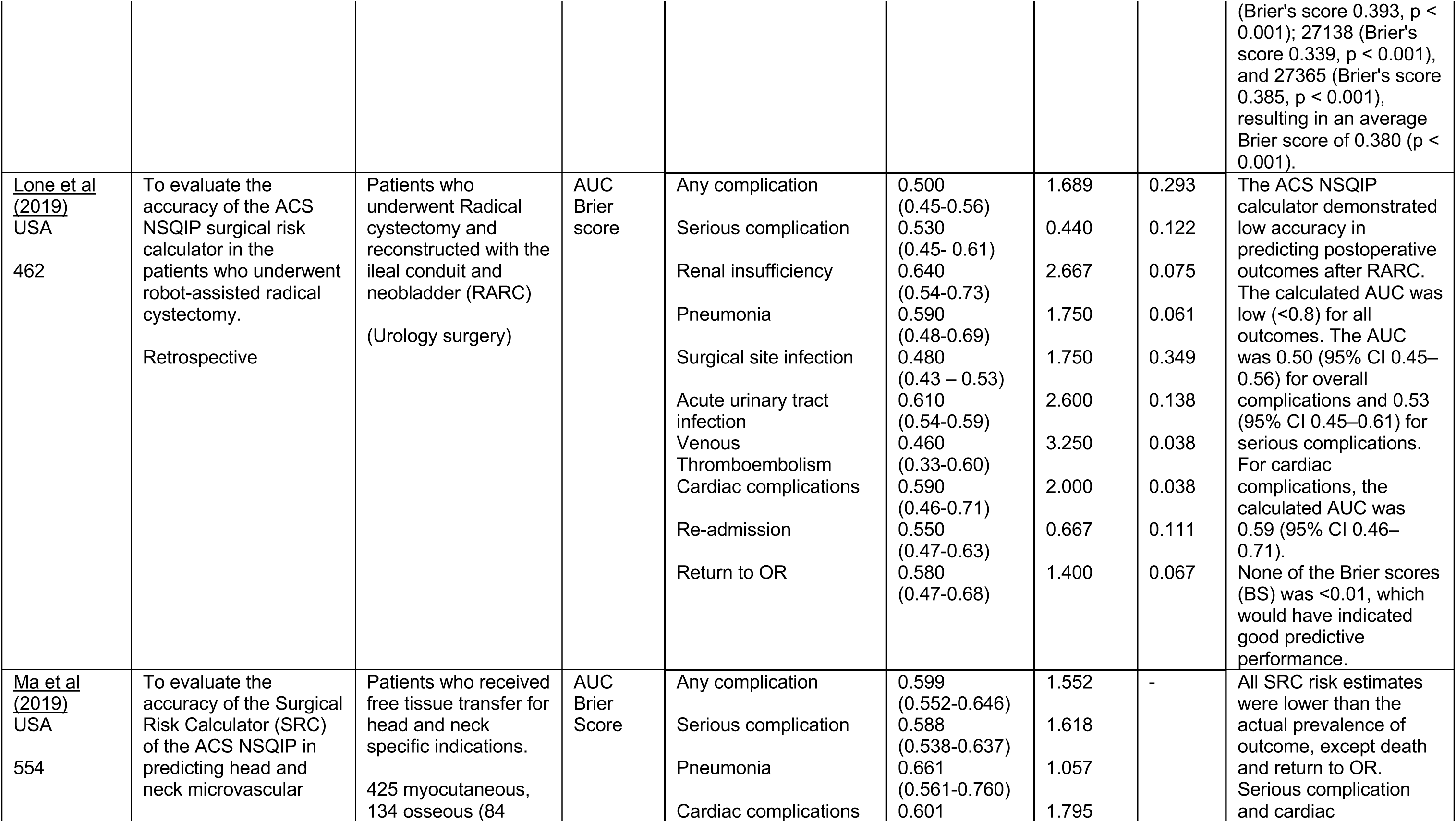
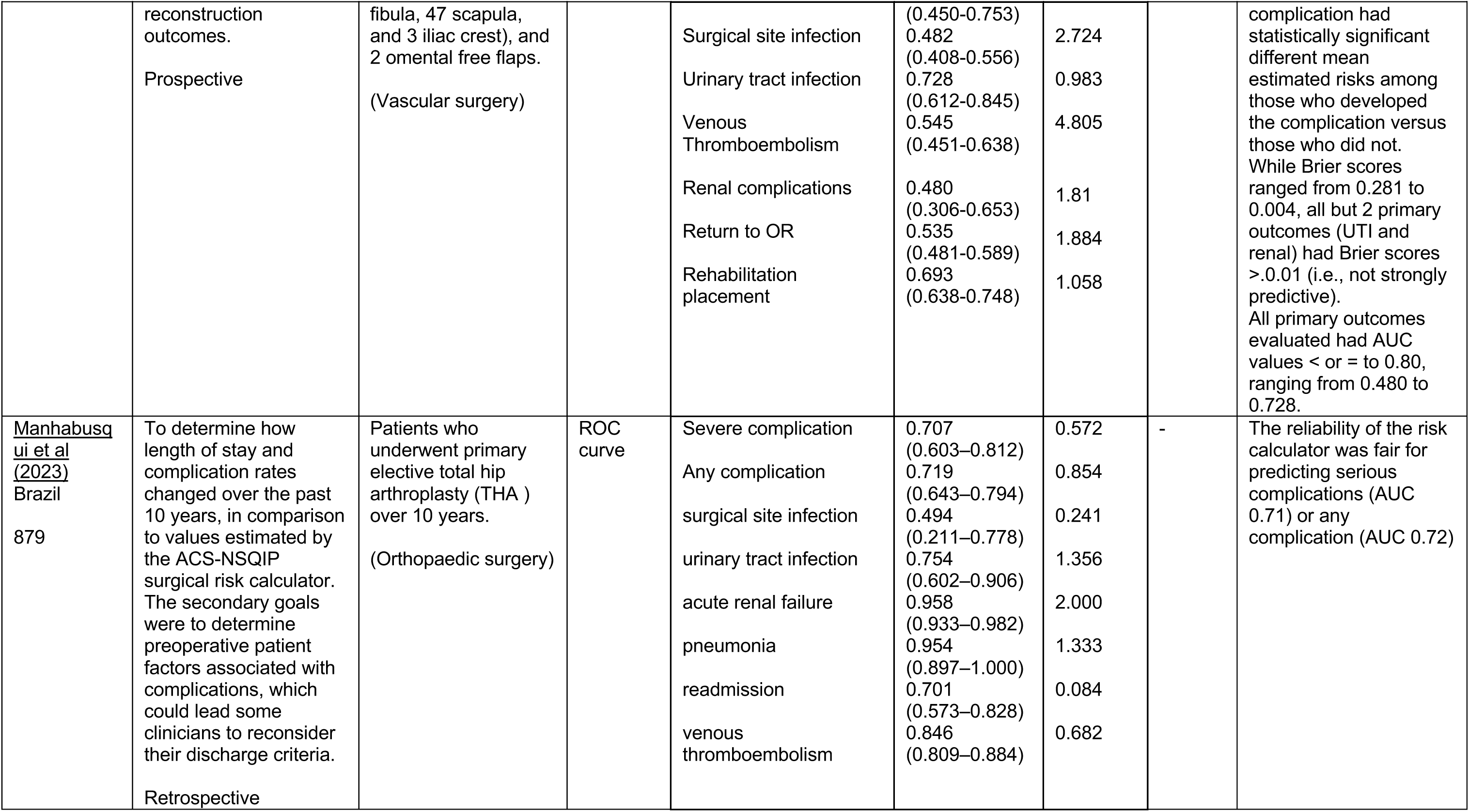
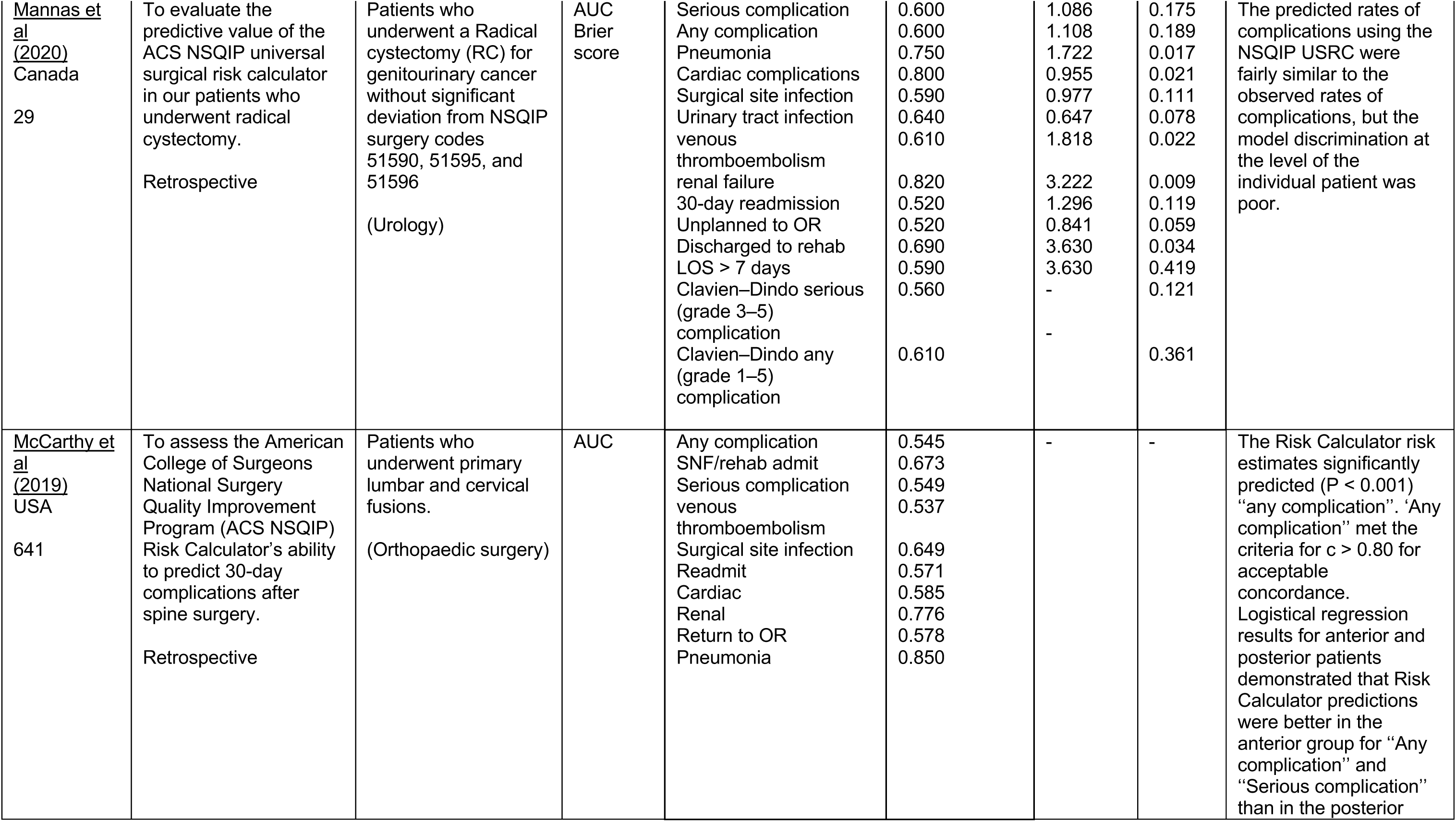
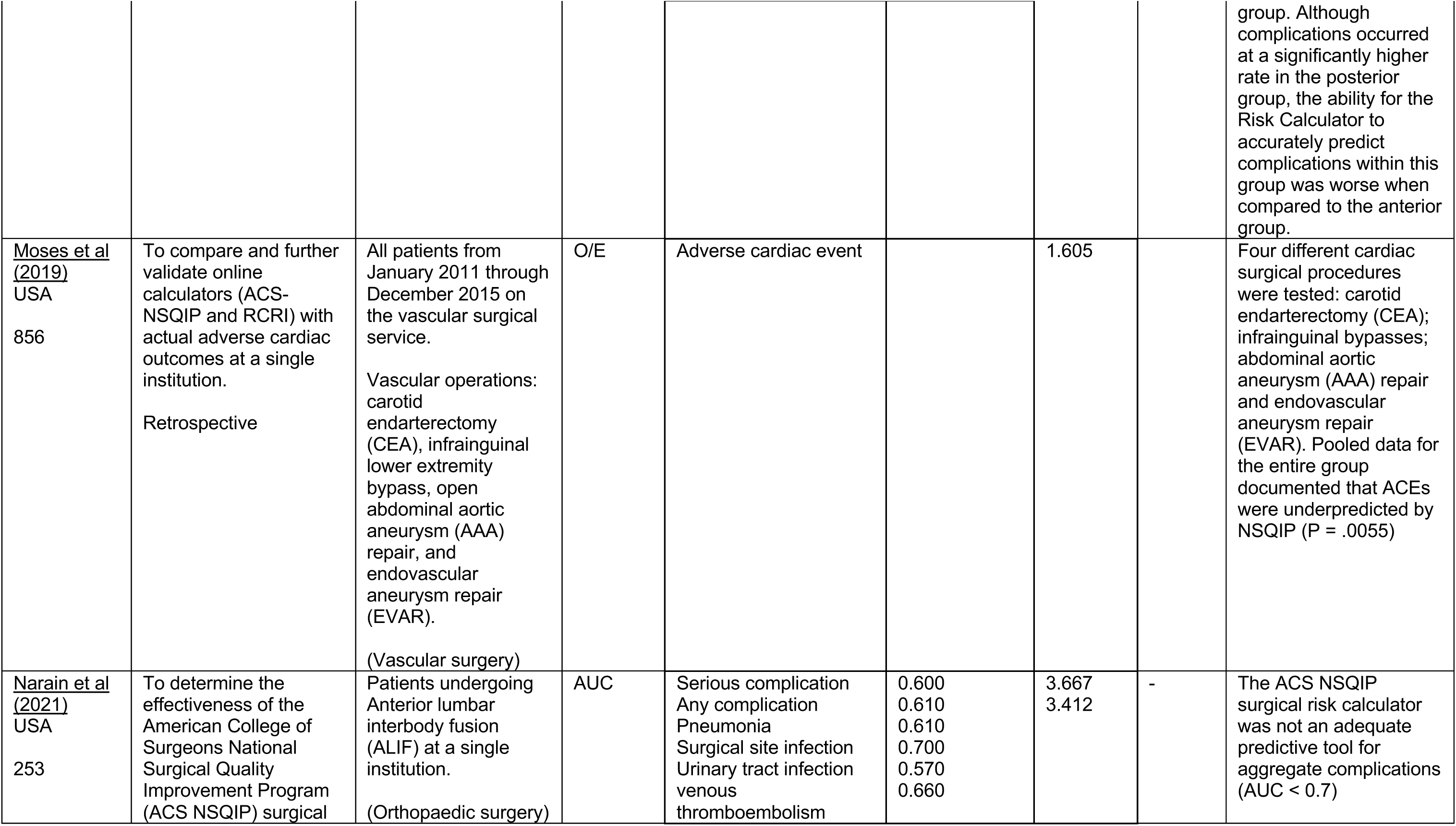
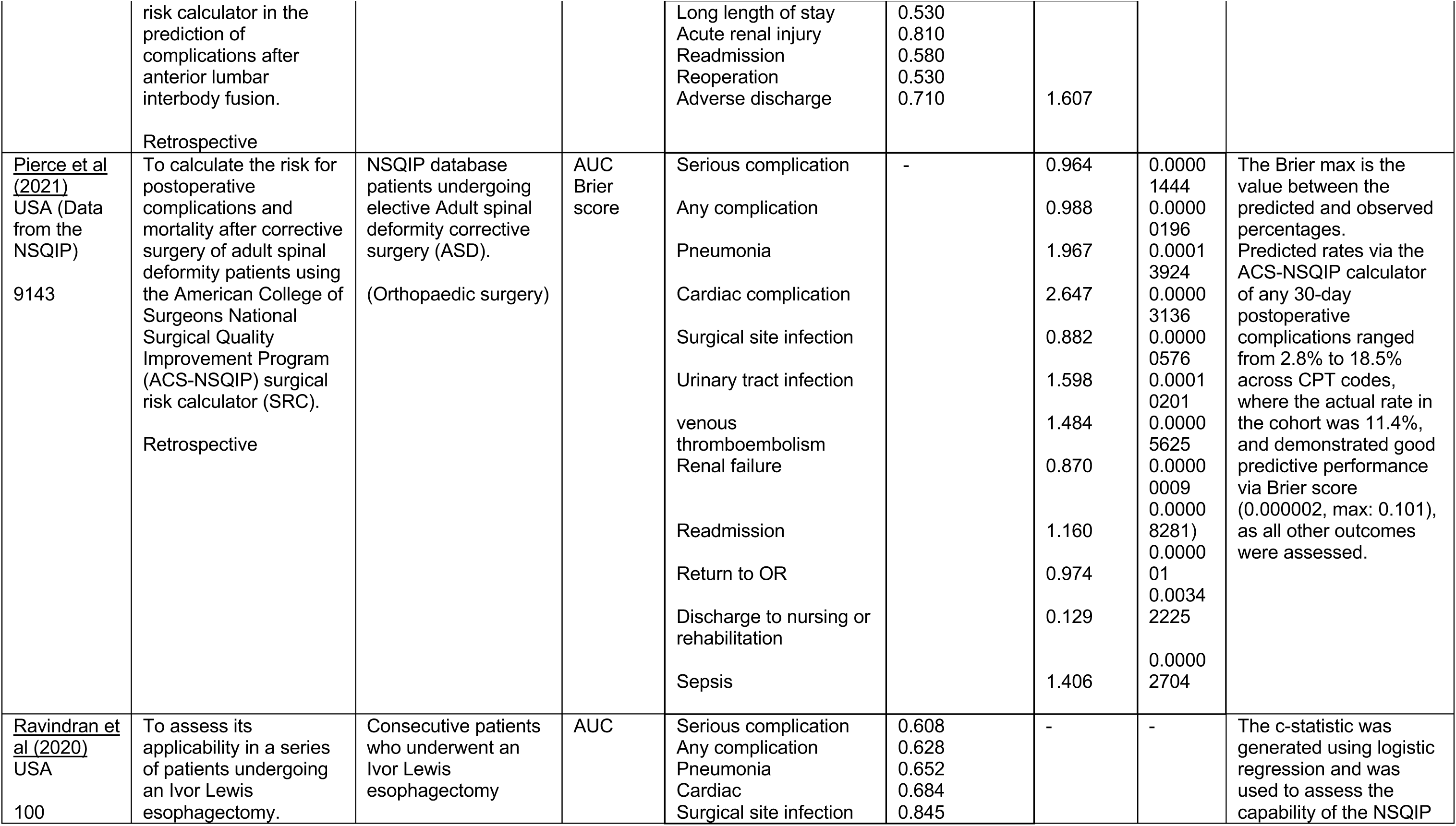
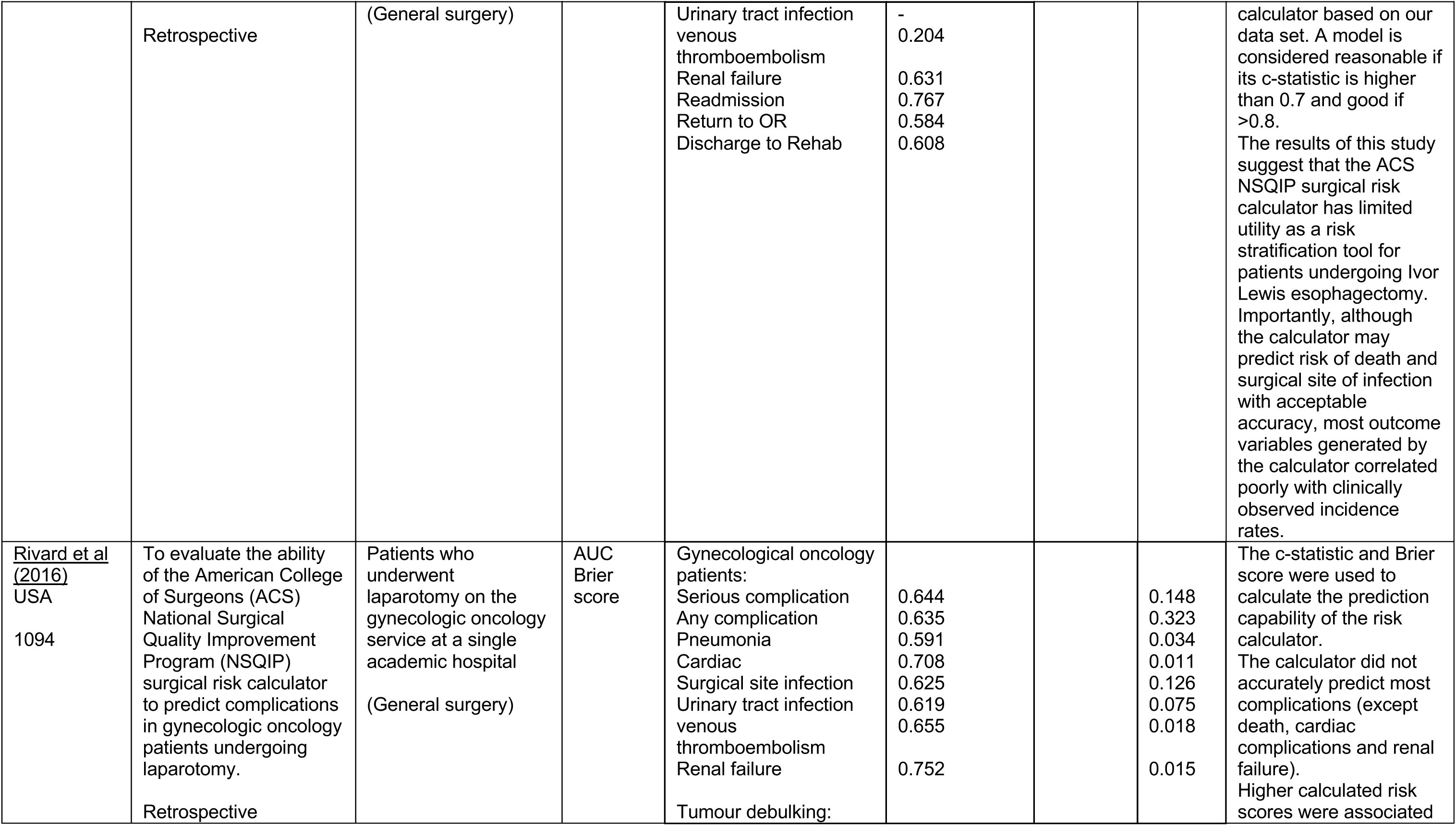
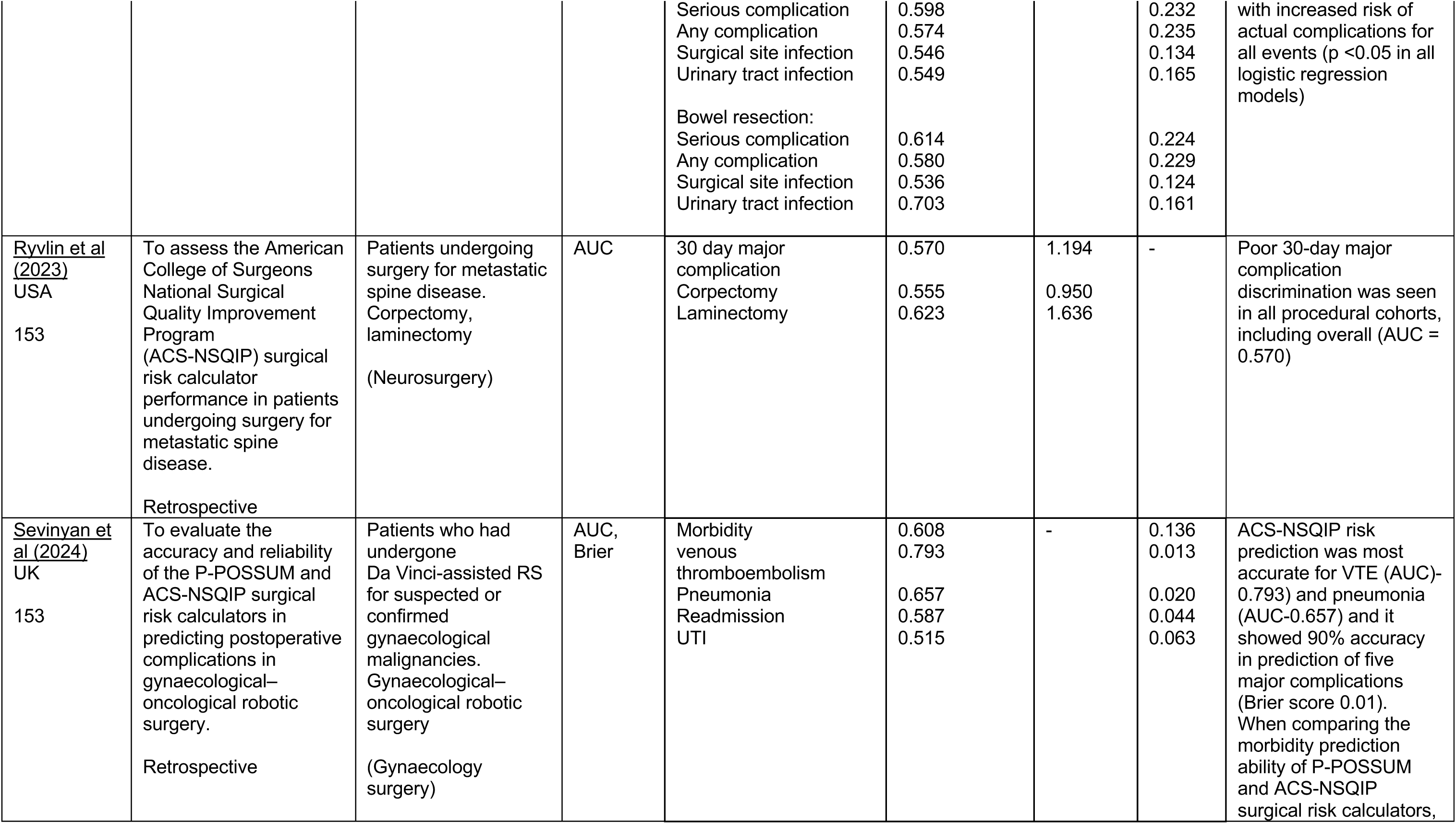
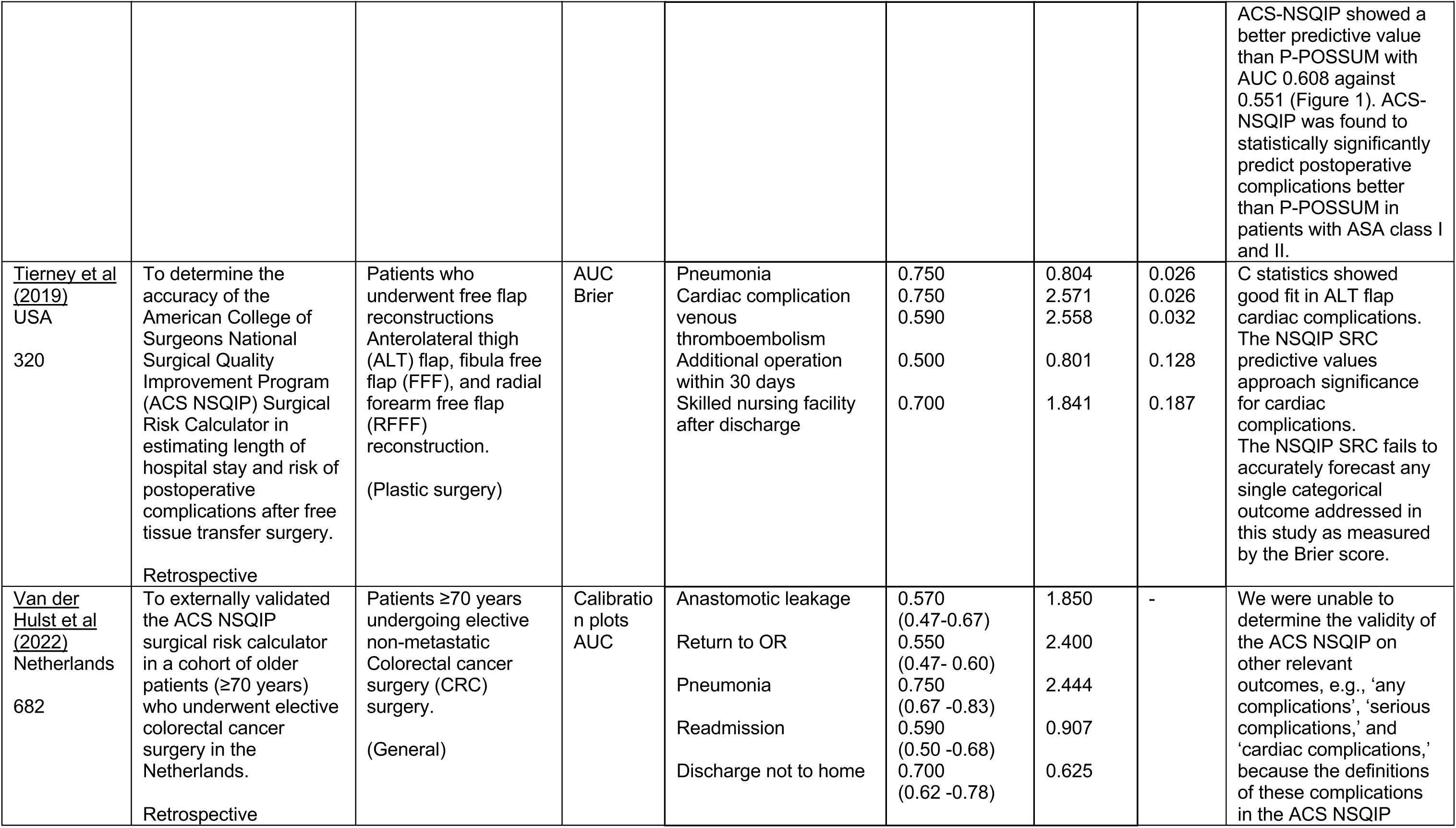
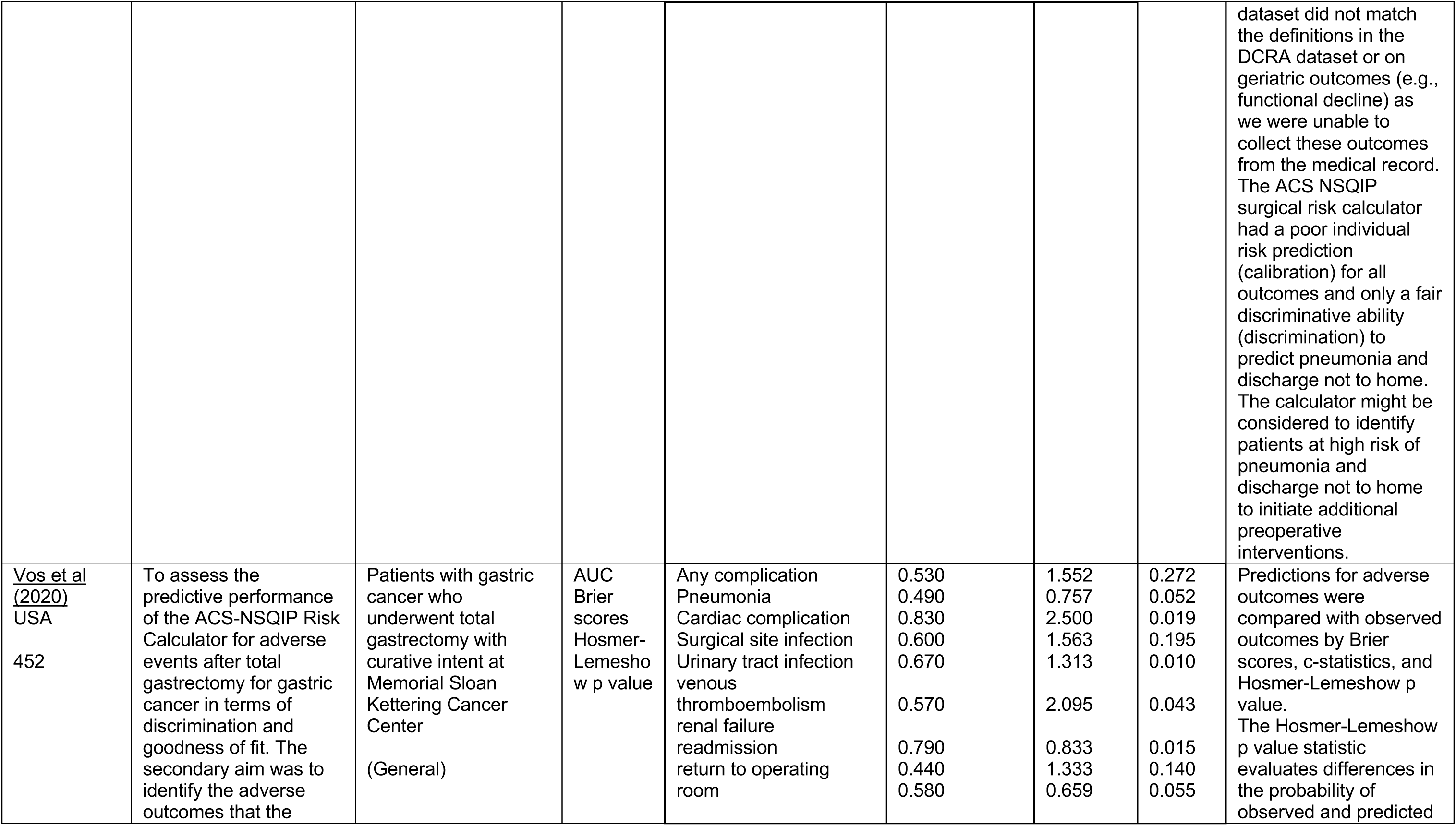
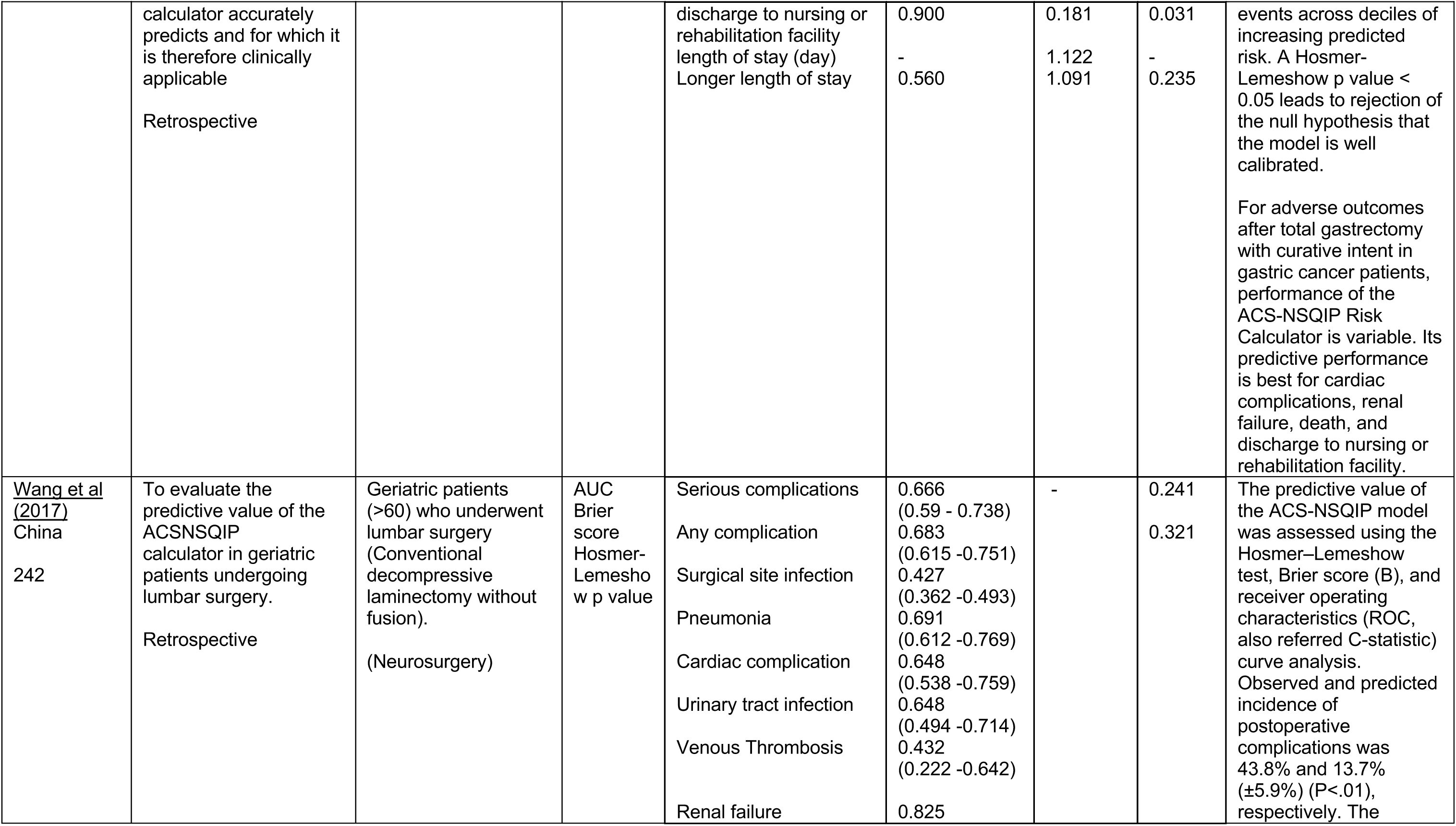
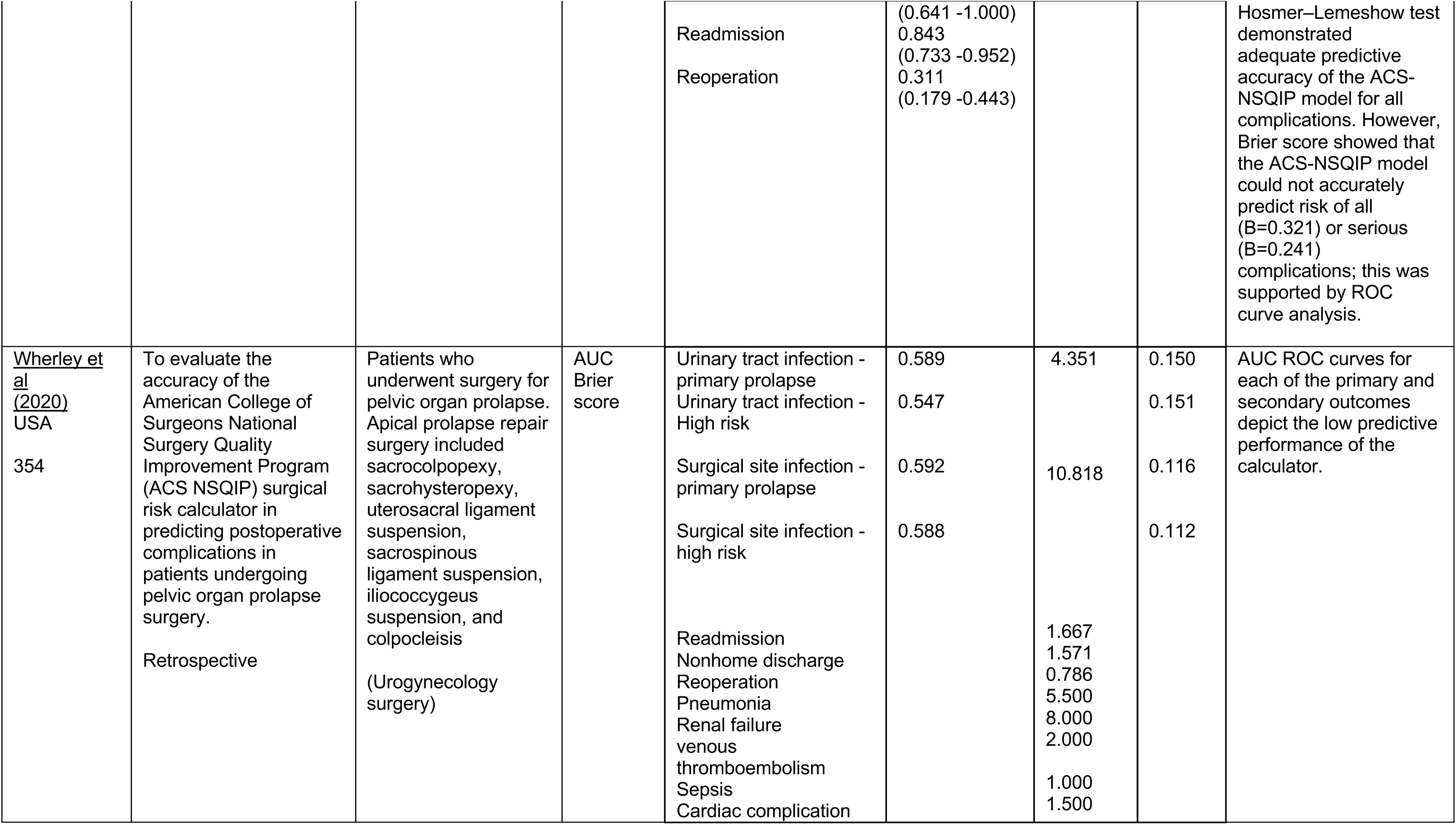
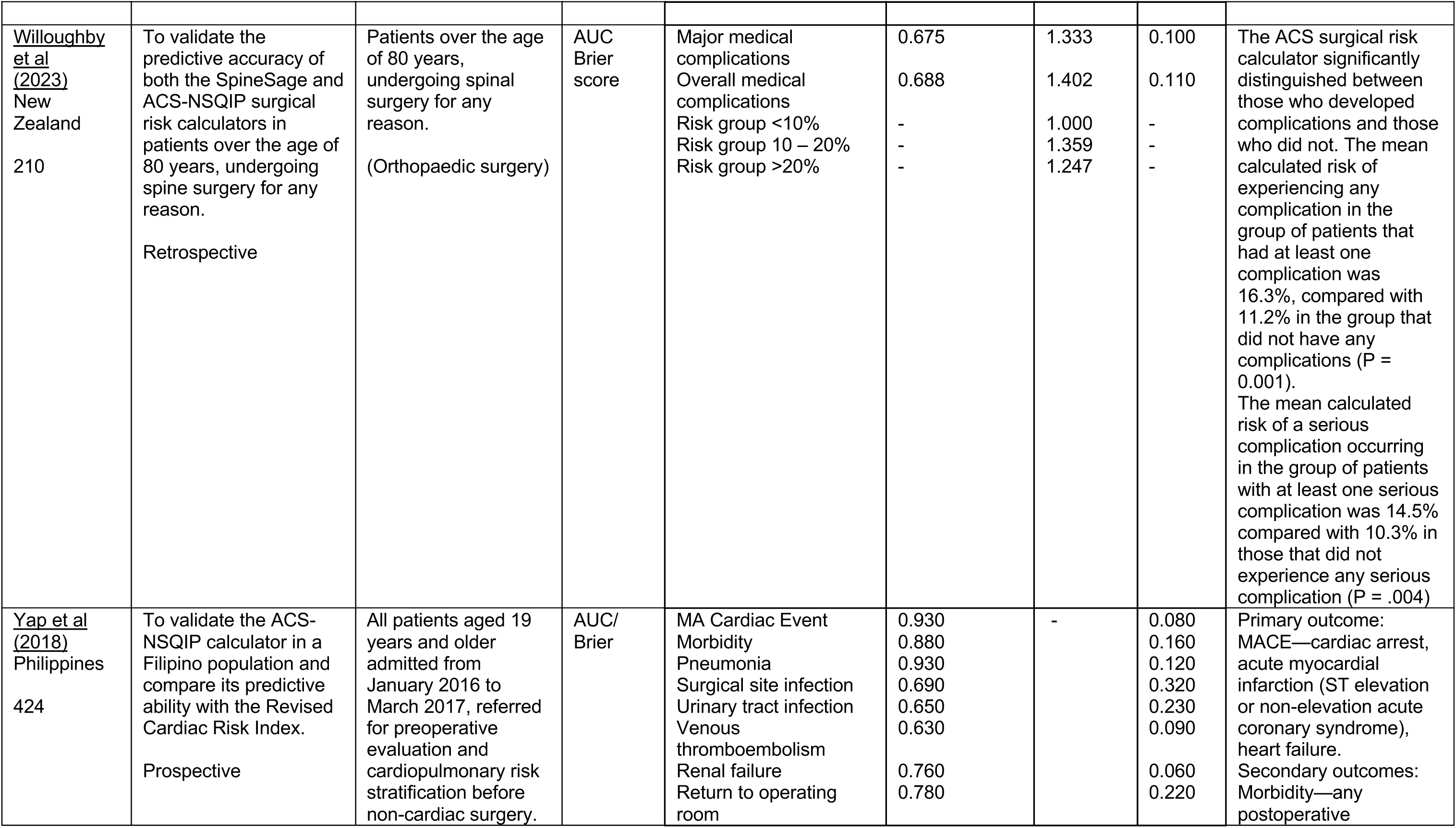
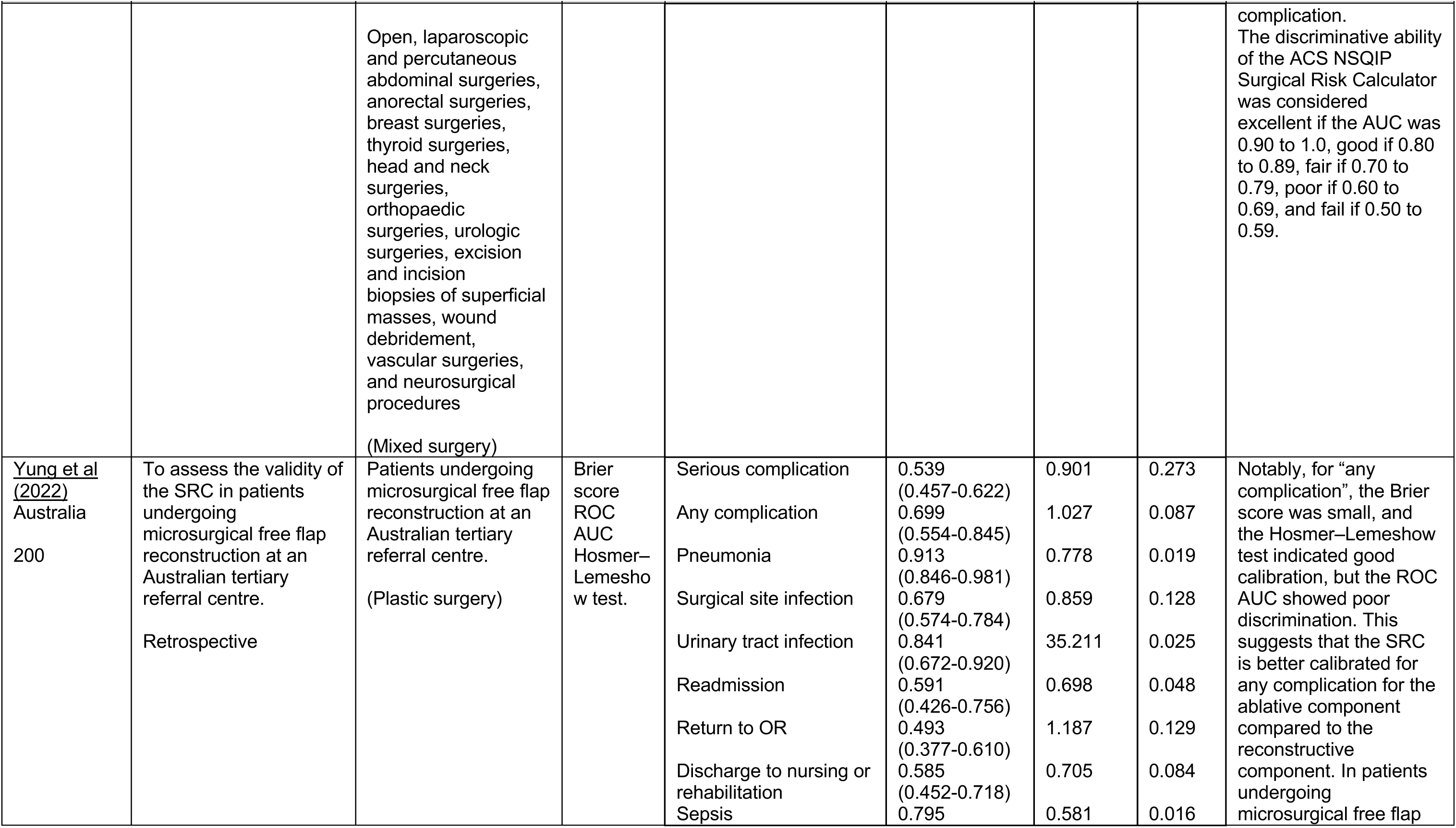
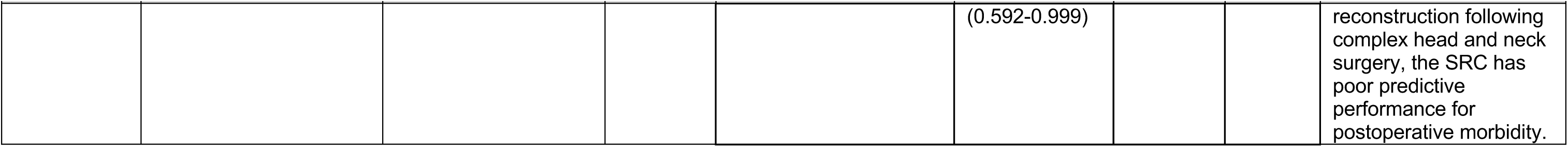
Data Extraction Tables: ACS NSQIP.

**Table 15.** Data Extraction Tables: ASA classification system.

| REF<br>Country<br>Sample<br>size | Aim and design | Population,<br>surgery and<br>(surgical specialty<br>category) | Validation<br>measures | Outcomes | C-statistic | O/E<br>ratio | Brier<br>score | Comments |
| --- | --- | --- | --- | --- | --- | --- | --- | --- |
| <u>Aceto et al</u><br><u>2021</u><br>Italy<br>105 | The primary aim of this prospective cohort study was to evaluate the usefulness of the modified Frailty Index (mFI) score to predict postoperative pulmonary complications (PPCs) in elderly patients undergoing major open abdominal surgery. The secondary purpose was to compare the prediction power of mFI, Ariscat (Assess Respiratory Risk in Surgical Patients in Catalonia), and American Society physical status classification (ASA) scores.<br>Prospective | Patients aged ≥65 years undergoing Open major upper elective (partial/total colectomy; Hartmann's procedure; total/partial gastrectomy; liver resection; and pancreaticoduodenectomy) and lower (nephrectomy, prostatectomy or hysterectomy) abdominal surgery.<br>(Mixed surgery) | ROC/AUC | Postoperative pulmonary complications | 0.690 | - | - |  |
| Bronheim et al 2018<br>USA<br>52066 | To determine the ability of RCRI to predict non-cardiac adverse events after Posterior Lumbar Decompression (PLD)<br>Retrospective | Adults (≥18 years), undergoing Posterior Lumbar Decompression (PLD) between 2006 and 2014<br>(Orthopaedic) | AUC | Unplanned intubation<br>Pulmonary embolism<br>Ventilated >48 hours<br>Acute renal failure<br>Cerebrovascular accident/stroke with neurologic deficit<br>Coma >24 hours<br>Sepsis<br>Septic shock<br>Reoperation<br>Superficial Surgical site infection (SSI)<br>Deep incisional SSI<br>Organ space SSI<br>Wound dehiscence<br>Pneumonia<br>Progressive renal insufficiency<br>Urinary tract infection<br>Peripheral nerve injury<br>Bleeding transfusions<br>Deep vein thrombosis/thrombophlebitis<br>Readmission- | 0.741 (0.736 – 0.745)<br>0.812(0.808 – 0.816)<br>0.742 (0.73.8 – 0.747)<br>0.789 (0.784 – 0.793)<br>0.838 (0.835 – 0.842)<br>0.653 (0.648 – 0.658)<br>0.905 (0.902 – 0.908)<br>0.759 (0.755 – 0.764)<br>0.866 (0.862 – 0.870)<br>0.842 (0.839 – 0.845)<br>0.950 (0.948 – 0.953)<br>0.776 (0.772 – 0.780)<br>0.792 (0.788 – 0.796)<br>0.824 (0.820 – 0.828)<br>0.814 (0.810 – 0.819)<br>0.825 (0.821 – 0.828)<br>0.513 (0.507 – 0.518)<br>0.800 (0.796 – 0.804)<br>0.782 (0.778 – 0.787)<br>0.906 (0.903 – 0.909) | - | - | ASA status had a fair discriminative ability (AUC = 0.770) for a composite of any non-cardiac complication. |
| Bronheim et al 2019<br>USA<br>52066 | To determine the ability of Revised Cardiac Risk Index (RCRI) to predict adverse cardiac events following posterior lumbar decompression<br>Retrospective | Adult (≥18 years), undergoing Posterior Lumbar Decompression PLD between 2006 and 2015<br>(Orthopaedic surgery) | AUC | Cardiac arrest requiring CPR<br>Myocardial Infarction | 0.674 (0.669 – 0.678)<br>0.799 (0.795 – 0.803) | - | - | ASA had discriminative abilities of “fair” for MI (AUC = 0.799) and “poor” for cardiac arrest requiring CPR (AUC = 0.674) |
| Chrisant et al 2024 | To assess the accuracy of RCRI compared to the ASA-PS classification | Patients ≥18 admitted for elective | AUC | Pulmonary complications | 0.760 (0.66 – 0.87)<br>0.750 (0.59 – 0.91) |  |  | There was no significant difference in the predictive ability |
| Tanzania<br>184 | system in preoperative risk assessment for pulmonary and cardiac problems among non-cardiothoracic surgery patients<br>Prospective | non-cardiothoracic surgery<br>(Mixed) |  | Cardiac complications) |  |  |  | of the two tools. Both RCRI and ASA-PS classification systems were equally effective in predicting these complications. |
| <u>Feghali et al (2022)</u><br>USA<br>The institutional database included 275 patients (88 clip placement, 187 endovascular treatment). AND 1047 patients who underwent clip placement were included in the NSQIP database. | To validate the utility of mFI-5 for predicting endovascular and microsurgical treatment outcomes in patients with unruptured aneurysms.<br>Retrospective | Patients with unruptured aneurysm who were treated with clip placement or endovascular therapy between January 2017 and December 2018 from the institutional database and NSQIP database<br>(Vascular surgery) | AUC<br>C-statistic | Major complications for clip placement<br>Major complications for endovascular therapy | 0.541<br>0.606 |  |  | Goodness of fit was indicated by a non-significant p value determined with the Hosmer-Lemeshow test. The AUC of ASA was 0.606 (p = 0.219). |
| <u>Fu et al</u><br>(2018)<br>USA<br>10,527 | Research questions: (1) Which demographic/anthropometric variable among age, sex, and body mass index (BMI) has the best discriminative ability as measured by receiver operating characteristics (ROC) in its association with adverse events following Total Shoulder Arthroplasty? (2) Which comorbidity index, among the American Society of Anesthesiologists (ASA) classification, the mCCI, or the mFI, has the best ROC in its association with adverse events following Total Shoulder Arthroplasty?<br>Retrospective | Patients who underwent Total shoulder arthroplasty (TSA) from 2005 to 2015 were identified from the National Surgical Quality Improvement Program (NSQIP)<br>(Orthopaedic surgery) | AUC | Extended length of stay<br>Discharge to higher level of care | 0.630 (0.614-0.646)<br>0.630 (0.616-0.645) |  |  |  |
| <u>Hightower et al</u><br>2010<br>USA<br>32 | To compare the risk predictive value of preoperative physiological capacity (PC: defined by gas exchange measured during cardiopulmonary exercise testing) with the ASA physical status classification in the same patients (n=32) undergoing major abdominal cancer surgery. | Patients (18+) undergoing elective major abdominal cancer surgery (Gastrectomy, Pancreatectomy, Radical cystectomy, Radical nephrectomy, Radical transabdominal tumour debulking, Pelvic exenteration, Low anterior | AUC | Postoperative morbidity | 0.688 (0.52315 - 0.85185) |  |  | Three newly identified PC measures and the ASA rank were significantly associated with postoperative morbidity; none showed a statistically greater association compared with the others. |
|  | Prospective | resection,<br>Retroperitoneal<br>lymph node<br>dissection).<br>(General surgery) |  |  |  |  |  |  |
| <u>Lim et al</u><br>(2017)<br>USA<br>6,148 | This study aimed to<br>evaluate the predictive<br>value of ASA<br>classification system on<br>30- day morbidity<br>following single-level<br>elective anterior cervical<br>discectomy and fusion.<br>Retrospective | Patients who<br>underwent Single-level elective<br>anterior cervical<br>discectomy and<br>fusion (SLE-ACDF)<br>between 2011 and<br>2013 were selected<br>from the NSQIP<br>database.<br>(Orthopaedic<br>surgery) | C-statistic | Any complication<br>Catastrophic outcome<br>Airway complication<br>Pneumonia<br>Unplanned intubation<br>Ventilator >48hours | 0.706 (0.48–1.41)<br>0.793 (0.39–2.71)<br>0.719 (0.67–7.29)<br>0.700 (0.33–4.56)<br>0.678 (0.41–5.36)<br>0.825 (0.69–50.96) |  |  | C-statistics used in the<br>regression models<br>demonstrate sufficient<br>discriminability of<br>these models. |
| <u>McConaghy et al</u><br>(2021)<br>USA<br>202,488<br>(THA)<br>230,823<br>(TKA) | The purpose of this study<br>was to review the (1)<br>mCCI; (2)<br>ASA physical status<br>classification system; (3)<br>ECM; and (4) mFI-5 to<br>determine their ability to<br>accurately predict 30-day<br>mortality, 30-day rate of<br>major and minor<br>complications,<br>discharge disposition,<br>and extended length of<br>stay in the hospital<br>following Total hip<br>arthroplasty or Total knee<br>arthroplasty<br>Retrospective | Patients who<br>underwent elective<br>Total hip<br>arthroplasty or Total<br>knee arthroplasty<br>from January 2011<br>through December<br>2019 from the<br>NSQIP database<br>(Orthopaedic<br>surgery) | C-statistic | Any major<br>complications (30<br>days)<br>Any minor<br>complications (30<br>days)<br>Discharge to not home<br>Length of stay >1 day | THA 0.615 (0.609-0.621)<br>TKA 0.615 (0.607-0.617)<br>THA 0.608 (0.603-0.613)<br>TKA 0.608 (0.603-0.611)<br>THA 0.640 (0.636-0.642)<br>TKA 0.639 (0.632-0.636)<br>THA 0.580 (0.572-0.582)<br>TKA 0.561 (0.559-0.563) |  |  | ASA was not<br>predictive of major and<br>minor complications. |
| <u>Meng et al</u><br>(2018)<br>USA<br>1,516 | Primary study aim was to compare the discriminative ability of the ASA, CCI, and mFI with the occurrence of postoperative adverse outcomes in patients undergoing Radical Cystectomy.<br>Retrospective | Patients undergoing elective Radical cystectomy (RC) for bladder cancer were extracted from the 2005 to 2011 NSQIP database<br>(Urology surgery) | AUC | Any adverse event<br>Minor adverse event<br>Serious adverse event<br>Infectious adverse events<br>Length of stay<br>Discharge to a higher level of care | 0.510 (0.485-0.534)<br>0.514 (0.486-0.542)<br>0.519 (0.491-0.547)<br>0.517 (0.490-0.543)<br>0.536 (0.510-0.561)<br>0.590 (0.556-0.624) |  |  |  |
| <u>Ondeck et al</u><br>(2018)<br>USA<br>16,495 | This study aimed to compare the discriminative ability of ASA, mCCI, and mFI, as well as demographic factors including age, body mass index, and gender for perioperative adverse outcomes following Posterior lumbar fusion.<br>Retrospective | Patients undergoing elective posterior lumbar fusion (PLF) with or without interbody fusion were extracted from the 2011–2014 NSQIP database.<br>(Orthopaedic surgery) | AUC | Any adverse event<br>Severe adverse event<br>Minor adverse event<br>Infectious adverse event<br>Extended length of stay<br>Discharge to higher level of care | 0.567 (0.555–0.579)<br>0.571 (0.553–0.588)<br>0.571 (0.557–0.585)<br>0.580 (0.560–0.599)<br>0.564 (0.556–0.572)<br>0.631 (0.621–0.641) |  |  | The combination of the best overall performing comorbidity index across the board (ASA) and demographic factor across the board (age) for the occurrence of any adverse event following PLF led to an AUC of 0.603 (95% CI: 0.590–0.616), which was statistically better than ASA alone and age alone. |
| <u>Sankar et al</u><br>(2014)<br>Canada<br>10,864 | The primary objective was to evaluate the inter-rater agreement of ASA-PS scores assigned at outpatient preoperative assessment clinics vs operating theatres.<br>Retrospective | Adult patients (≥18 yr) who underwent elective non-cardiac surgery (ENT surgery, General surgery, Gynaecology, Neurosurgery, Ophthalmology, Orthopaedic surgery, Plastic surgery, Thoracic surgery, Urology,<br>Vascular surgery).<br>(Mixed surgery) | AUC | Myocardial injury (Ratings in the clinic)<br>Myocardial injury (Ratings in the operating theatre) | 0.700 (0.65 – 0.75)<br>0.750 (0.71–0.79) |  |  |  |
| <u>Wolters</u><br><u>et al</u><br><u>2006</u><br>Germany<br>107 | To prospectively apply<br>various scoring systems<br>in order to estimate<br>outcome in patients<br>undergoing<br>aortobifemoral surgery<br>due to arterial occlusive<br>disease at the aorto-iliac<br>level<br>Prospective | Patients who<br>received an<br>aortobifemoral or<br>aorto-iliac graft due<br>to arterial occlusive<br>disease between<br>1996 to 2000.<br>(Vascular surgery) | AUC | Morbidity | 0.518 |  |  | In this study there was<br>no significant<br>correlation between<br>the ASA classification<br>and the postoperative<br>mortality or morbidity. |

**Table 16.** Data Extraction Tables: RCRI.

| REF<br>Country<br>Sample<br>Size | Aim and<br>design | Population, surgery<br>and (surgical specialty<br>category) | Validation<br>measures | outcomes | C-statistic | O/E<br>ratio | Brier<br>score | Comments |
| --- | --- | --- | --- | --- | --- | --- | --- | --- |
| <u>Alphonsus</u><br><u>et al</u><br><u>(2021)</u><br>South<br>Africa<br>3,045 | Using RCRI to<br>quantify<br>perioperative<br>risk for<br>complications<br>in these<br>patients.<br>Retrospective | Patients aged ≥45 years<br>presenting for surgery in<br>the combined SASOS<br>and ASOS data set. Non-cardiac surgery<br>(Orthopaedic, Breast,<br>Obstetrics, Gynaecology,<br>Upper gastrointestinal,<br>Lower gastrointestinal,<br>Hepatobiliary, Urology,<br>Vascular, Head and<br>Neck, Plastics, Thoracic,<br>Neurosurgery, other)<br>(Mixed surgery) | AUC | Cardiac complications | **0.680 (0.57 -0.79)** | - | - | The discrimination of<br>the RCRI for cardiac<br>complications was<br>described using the<br>receiver operating<br>characteristic (ROC)<br>and area under the<br>curve (AUC). The<br>calibration between the<br>original derivation<br>cohort and the ASOS<br>cohort was compared<br>using the RR for<br>cardiac complications<br>in the African cohort for<br>each RCRI risk factor. |
| | | | | | | | | The likelihood ratios for low- (0 RCRI risk factors), intermediate- (1 - 2 RCRI risk factors) and high-risk ($\geq 3$ RCRI risk factors) categories were calculated. |
| <u>Alrezk et al (2017)</u><br>USA<br>172,905 | To investigate the performance of the RCRI and Gupta MICA perioperative cardiac risk models in a geriatric population. Retrospective | NSQIP geriatric cohort >65 non-cardiac surgery (Hernia, Anorectal, Aortic, Bariatric, Brain, Breast, ENT, Foregut/ hepatopancreatobiliary, GBAAS/intestinal, Neck, Obstetric/gynecologic, Orthopedic, Other abdomen, Peripheral vascular, Skin, Spine, Thoracic, Vein, Urology)<br>(Mixed surgery) | AUC | Cardiac risk | 0.680 (0.67-0.69) |  |  | Although the RCRI underestimates risk for low-risk patients, it is well calibrated for the highest-risk group (2.4% to 5.4% in RCRI).<br>The RCRI was not derived to predict the cardiac risk within 30 days of surgery but is aimed solely at predicting the risk during a hospital stay. In the modern world, using equations developed so long ago and on unique populations (i.e., RCRI) is of questionable value, particularly in an era when curated data sources such as the NSQIP and other large data sets are readily available. |
| <u>Andersson et al (2015)</u> | To assess the relationship of RCRI with | Patients >25 undergoing major elective, non-cardiac surgery (Ear- | C-Statistic | Major adverse cardiovascular events | 0.761 | - | - | Modelling RCRI classes as a continuous variable, C |
| Denmark<br>447,352 | major adverse cardiovascular events in an unselected cohort of patients undergoing elective, non-cardiac surgery.<br>Retrospective | nose-throat, Major orthopedic, Minor orthopedic, Abdominal (bowel) Abdominal (nonbowel), Breast, Plastic, Endocrine, Eye, Female reproductive, Male reproductive, Intracranial, Neuro, Nonarterial vessels, Thoracic, Urologic, Vascular)<br>**(Mixed surgery)** | | | | | | statistic was highest among age group 56 to 65 years (0.772) and lowest for those aged >85 years (0.683). In a nationwide unselected cohort, the performance of the RCRI was similar to that of the original cohort. Having $\geq 1$ risk factor was of moderate sensitivity, but high negative predictive value for all ages. |
| <u>Archan et al (2010)</u><br>Austria<br>225 | To determine if the Revised Cardiac Risk Index (Lee) is useful for stratification of patients by risk of both perioperative cardiac morbidity and long-term all-cause mortality<br>Retrospective | Patients with abdominal aortic aneurysms admitted to the authors' institution from 1999 to 2006. All patients underwent endovascular aortic aneurysm repair<br>(Vascular surgery) | AUC | Postoperative cardiac events | 0.730 | | | The AUCs for the prediction of postoperative cardiac events by age and GAS (Glasgow Aneurysm Score) were 0.60 and 0.67, respectively.<br>In the present study, the authors found a highly significant association between a Lee index $\geq 3$ and postoperative cardiac events (p 0.004). These results indicate that the Lee index, even if its overall discriminatory ability is fair at best, is a useful tool for the identification of high- risk patients from both a short- and long-term perspective. The Lee index was developed for the prediction of prospectively detected major cardiac complications. |
| <u>Bronheim et al 2018</u><br>USA<br>52066 | To determine the ability of RCRI to predict non-cardiac adverse events after Posterior Lumbar Decompression (PLD)<br>Retrospective | Adult (≥18 years), undergoing Posterior Lumbar Decompression (PLD) between 2006 and 2014<br>(Orthopaedic surgery) | AUC | Unplanned intubation<br>Pulmonary embolism<br>Ventilated >48 hours<br>Acute renal failure<br>Cerebrovascular accident/stroke with neurologic deficit<br>Coma >24 hours<br>Sepsis<br>Septic shock<br>Reoperation<br>Superficial Surgical site infection (SSI)<br>Deep incisional SSI<br>Organ space SSI<br>Wound dehiscence<br>Pneumonia<br>Progressive renal insufficiency<br>Urinary tract infection<br>Peripheral nerve injury<br>Bleeding transfusions<br>Deep vein thrombosis/thrombophlebitis<br>Readmission | 0.837 (0.833-0.842)<br>0.409 (0.403 -0.415)<br>0.845 (0.841 -0.850)<br>0.881 (0.876 -0.887)<br>0.747 (0.742 -0.753)<br>0.900 (0.868 -0.933)<br>0.826 (0.821 -0.830)<br>0.845 (0.840 -0.850)<br>0.850 (0.845 -0.855)<br>0.717 (0.711 -0.722)<br>0.879 (0.875 -0.883)<br>0.876 (0.872 -0.879)<br>0.717 (0.712 -0.723)<br>0.739 (0.733 -0.744)<br>0.845 (0.839 -0.851)<br>0.738 (0.733 -0.744)<br>0.073 (0.071 -0.076)<br>0.712 (0.706 -0.717)<br>0.707 (0.701 -0.713)<br>0.835 (0.829 -0.840) |  |  | RCRI had a poor discriminative ability (AUC = 0.623) for a composite of any non-cardiac complication. |
| <u>Bronheim et al 2019</u><br>USA<br>52066 | To determine the ability of Revised Cardiac Risk Index (RCRI) to predict adverse | Adult (≥18 years), undergoing Posterior Lumbar Decompression (PLD) between 2006 and 2015 | AUC | Cardiac arrest requiring CPR<br>Myocardial Infarction | 0.855 (0.851-0.860)<br>0.876 (0.872-0.880) |  |  | RCRI had a good discriminative ability to predict both MI [area under the curve (AUC) = 0.876] and cardiac |
|  | cardiac events<br>following<br>posterior<br>lumbar<br>decompression<br>Retrospective | (Orthopaedic surgery) |  |  |  |  |  | arrest requiring CPR<br>(AUC = 0.855). |
| <u>Carabini et al</u><br>(2014)<br>Lebanon<br>547 | To determine<br>the accuracy of<br>the RCRI in<br>predicting<br>MACE in<br>patients<br>undergoing<br>spine fusion<br>surgery of 3 or<br>more bony<br>levels.<br>Retrospective | Patients undergoing<br>spine fusion surgery of 3<br>levels or more<br>(Orthopaedic surgery) | AUC | Major adverse cardiac events | 0.540 (0.47-0.61) |  |  | The RCRI did not<br>predict cardiac<br>morbidity in our<br>patients undergoing<br>major spine fusion<br>surgery, despite being<br>extensively validated in<br>low-risk non-cardiac<br>surgical patients. |
| <u>Chrisant et al</u> 2024<br>Tanzania<br>184 | To assess the<br>accuracy of<br>RCRI<br>compared to<br>the ASA-PS<br>classification<br>system in<br>preoperative<br>risk<br>assessment for<br>pulmonary and<br>cardiac<br>problems<br>among non-cardiothoracic<br>surgery<br>patients<br>Prospective | Patients ≥18 admitted for<br>elective non-cardiothoracic surgery<br>(Mixed surgery) | AUC | Pulmonary complications<br>Cardiac complications | 0.710 (0.59 - 0.83)<br>0.730 (0.56 – 0.91) |  |  | There was no<br>significant difference in<br>the predictive ability of<br>the two tools. Both<br>RCRI and ASA-PS<br>classification systems<br>were equally effective<br>in predicting these<br>complications. |
| <u>Cohn and</u><br><u>Ros</u><br><u>2018</u><br>USA<br>663 | This study compared these risk calculators and a reconstructed Revised Cardiac Risk Index in predicting postoperative cardiac complications, both during hospitalization and 30 days after operation, in a patient cohort who underwent select surgical procedures in various risk categories.<br>Retrospective | A patient cohort who underwent select surgical procedures in various risk categories Non cardiac surgery (15.7% high risk; 62.9% Intermediate risk; 21.4% low risk)<br>(Mixed surgery) | ROC | All cardiac complications (in hospital)<br>All cardiac complications (-30 days)<br>Major cardiac complications | 0.850<br>0.780<br>0.550 |  |  | In this study, the C-statistics (95% confidence interval) for RCRI and R-RCRI showed good discrimination for cardiac complications. |
| <u>Davis et al</u><br><u>(2013)</u><br>Canada<br>9,519 | To re-examine the validity of the inclusion of the two predictors, diabetes and chronic renal failure, in the RCRI using a large modern prospectively collected data set. | Patients over the age of 50 undergoing elective non-cardiac surgery with an expected length of stay > two days<br>(Abdominal, Orthopaedic, Thoracic, Vascular, genitourinary, neurosurgery, and ear nose and throat surgeries)<br>(Mixed surgery) | AUC | Major cardiac complications | 0.790 (0.76 - 0.83) | | | A simplified 5-Factor model using a high-risk type of surgery, a history of ischemic heart disease, congestive heart failure, cerebrovascular disease, and a preoperative GFR $\geq 30$ mLmin<sup>-1</sup> results in superior prediction of major cardiac complications following |
|  | Prospective |  |  |  |  |  |  | elective non-cardiac surgery compared to RCRI. |
| <u>Dunn et al (2019)</u><br>USA<br>503 | To compare the utility of the three different CV risk calculators using data from our own transplant center, to assess which of the models most accurately predicts long-term and short term Major Adverse Cardiac Events (MACE)<br>Retrospective | Transplant database, all adult patients who underwent kidney transplant from 2005 to 2010<br>(Urology / Transplant) | AUC | 30 day major adverse cardiac event<br>1 year major adverse cardiac event | 0.633<br>0.661 |  |  | This study focussed on which of the three models performed best: Of the three calculators, PORT performed best when the sensitivity was set at a clinically relevant level. |
| <u>Moses et al 2019</u><br>USA<br>856 | To compare and further validate online calculators with actual adverse cardiac outcomes at a single institution<br>Retrospective | All patients from January 2011 through December 2015 on the vascular surgical service. Vascular operations: carotid endarterectomy (CEA), infrainguinal lower extremity bypass, open abdominal aortic aneurysm (AAA) repair, and endovascular aneurysm repair (EVAR)<br>(Vascular surgery) | O/E | Adverse cardiac events | - | 2.231 | - | For open AAA repair RCRI strongly underpredicted the adverse events ($P \leq 0.0001$). Pooled data showed adverse cardiac events (ACEs) were underpredicted by RCRI ($P \leq .001$). |
| <u>Pandey et al</u><br>(2015)<br>USA<br>1,568 | To evaluate the association between the presence of stable anginal symptoms and risk for adverse postoperative cardiovascular outcomes in patients with a history of MI scheduled to undergo a major non-cardiac surgery.<br>Retrospective | Patients who underwent 1 of the 15 elective non-cardiac procedures who had a concomitant documented history of a recent MI from the NSQIP database (2005-2011). (Carotid Endarterectomy, Lower extremity bypass, Abdominal aortic aneurysm repair, Total hip replacement, Total knee replacement, Bariatric surgery, Esophagectomy, Gastrectomy, Pancreatectomy, Colectomy, Nephrectomy, Cystectomy, Prostatectomy, Hysterectomy, Pneumonectomy<br>(Mixed surgery) | AUC | Adverse cardiac event | 0.590 | - | - | We quantified the predictive accuracy of the RCRI and the modified RCRI (treating preoperative angina and MI as separate inputs in the score) in predicting an adverse cardiac outcome in the present patient population using the area under the curve (AUC) analysis. All input variables were given a score of 0 or 1 (0 = no, 1 = yes), in accordance with the original RCRI described. The Mantel-Haenszel test was used to assess the statistical significance of the difference in the discriminatory ability between the 2 models. |
| <u>Peterson et al</u><br>(2016)<br>USA<br>1,096 | To determine whether the NSQIP MICA risk calculator could accurately discriminate perioperative MICA in patients undergoing elective hip and knee<br>replacement<br>surgery and<br>compare with<br>RCRI.<br>Retrospective | Adult patients at Penn State Milton S. Hershey Medical Center from January 1, 2013, to December 31, 2014, Elective total hip replacement and total knee replacement surgeries<br>(Orthopaedic surgery) | C-Statistic | Adverse cardiac events | 0.90 (0.75-1) |  |  | The NSQIP and RCRI calculators perform comparably in discriminating adverse cardiac events in patients undergoing elective hip and knee surgery. |
| <u>Schmidt et al</u><br>(2024)<br>Germany<br>199 | To compare<br>the predictive<br>value of the<br>RCRI, AUB-HAS2, and<br>Andersson<br>scores with<br>preoperative<br>NT-proBNP for<br>the<br>postoperative<br>30-day<br>morbidity in an<br>observational<br>non-cardiac<br>and non-vascular<br>surgery cohort<br>Retrospective | Patients aged>65 years<br>scheduled for elective<br>non-cardiac non-vascular<br>surgery in general<br>anaesthesia with<br>intermediate or high<br>surgical risk (elective<br>intracranial, thoracic,<br>head and neck, trauma<br>and<br>orthopaedic, or<br>abdominal surgery,<br>including visceral,<br>urological, and<br>gynaecological<br>operations)<br>(Mixed surgery) | AUC/ROC | Morbidity<br>Rehospitalisation<br>Acute kidney injury<br>Infection<br>Acute decompensated heart<br>failure | 0.560<br>0.457<br>0.687<br>0.573<br>0.590 | - | - | AUB-HAS2, but not<br>RCRI or Andersson<br>score, significantly<br>predicted the CME<br>(AUB-HAS2: AUCROC<br>0.646,<br>p<0.001; RCRI:<br>AUCROC 0.560,<br>p=0.126; Andersson:<br>AUCROC 0.487,<br>p=0.760).<br>In our analysis, the<br>modified RCRI,<br>including the weighted<br>preoperative NT-proBNP cut-of and<br>original RCRI<br>components, showed<br>improved predictively<br>compared to the<br>original RCRI. |
| <u>Yap et al</u><br>(2018)<br>Philippines<br>3,045 | To validate the<br>ACS-NSQIP<br>calculator in a<br>Filipino<br>population and<br>compare its<br>predictive<br>ability with the<br>Revised | All patients aged 19<br>years and older admitted<br>from January 2016 to<br>March 2017, referred for<br>preoperative evaluation<br>and cardiopulmonary risk<br>stratification before non-cardiac surgery. Open,<br>laparoscopic and | AUC/ Brier | Major adverse cardiac event | 0.930 |  |  | Primary outcome:<br>MACE—cardiac arrest,<br>acute myocardial<br>infarction (ST elevation<br>or non-elevation acute<br>coronary syndrome),<br>heart failure.<br>Secondary outcomes:<br>2. Morbidity—any |
|  | Cardiac Risk<br>Index.<br>Prospective | percutaneous abdominal<br>surgeries, anorectal<br>surgeries, breast<br>surgeries, thyroid<br>surgeries, head and neck<br>surgeries, orthopaedic<br>surgeries, urologic<br>surgeries, excision and<br>incision<br>biopsies of superficial<br>masses, wound<br>debridement, vascular<br>surgeries, and<br>neurosurgical procedures<br>(Mixed surgery) |  |  |  |  |  | postoperative<br>complication |

**Table 17.** Data Extraction Tables: P-POSSUM.

| REF<br>Country<br>Sample<br>Size | Aim and design | Population, surgery and<br>(surgical specialty<br>category) | Validation<br>measures | Outcomes | C-statistic | O/E<br>ratio | Brier<br>score | Comments |
| --- | --- | --- | --- | --- | --- | --- | --- | --- |
| Angelucci<br>et al<br>2024<br>Italy<br>567 | To investigate the<br>accuracy of ACS-<br>NSQIP and P-POSSUM risk<br>calculators in<br>predicting<br>postoperative<br>outcomes for<br>patients<br>undergoing<br>retroperitoneal<br>sarcoma surgery | Adult patients (≥18),<br>undergoing elective<br>comprehensive resection<br>for primary or persistent<br>adult-type retroperitoneal<br>sarcoma with ASA score I–<br>IV<br>(General surgery) | AUC,<br>Brier<br>score | Any complication<br>Severe complication | 0.660 (0.61–0.70)<br>0.630 (0.58–0.68) |  | 0.229<br>0.205 |  |
|  | Retrospective |  |  |  |  |  |  |  |
| <u>Bodea et al</u><br>(2018)<br>Romania<br>113 | To evaluate the capacity of P-POSSUM risk scores concerning the morbidity and mortality following standard pancreaticoduodenectomy and pancreaticoduodenectomy associated with portal vein<br>Retrospective | Consecutive pancreaticoduodenectomy performed for perampullary malignant tumors during July 2013-December 2015.<br>(General surgery) | ROC | All complications<br>Pancreaticoduodenectomy specific complications | 0.610<br>0.640 | 1.660<br>0.890 |  | The predictive power of P-POSSUM score was higher in PD specific complications and subsequent mortality compared to the whole morbidity and mortality rates. |
| <u>He et al</u><br>2023<br>China<br>349 | To assess the suitability of POSSUM and its modified versions, E-PASS and its modified score, SRS, and SORT scores for predicting postoperative complications and mortality in patients undergoing laparoscopic radical gastrectomy for gastric cancer<br>Retrospective | Patients who were histopathologically diagnosed with gastric adenocarcinoma and underwent R0 resection (Laparoscopic radical gastrectomy)<br>(General surgery) | AUC | Early complications after laparoscopic radical gastrectomy | 0.781 |  |  | P-POSSUM performed moderately for the prediction of postoperative complication morbidity in laparoscopic surgery. P-POSSUM underestimated post-operative mortality. |
| <u>Jones et al</u><br>(2022)<br>Ireland<br>67 | To determine whether the risk estimates for the Portsmouth scoring system could be used in major head and neck reconstructive surgery.<br>Retrospective | Patients undergoing resection for a temporal bone malignancy in a single head and neck centre<br>(ENT surgery) | AUC | Complications<br>Morbidity | 0.766 | 1.013 |  | The optimal cut-off for the scoring system was then calculated using the Youden index from the receiver operating characteristic curve, which was 40.5 % in this case. Although it was a valid predictor of morbidity overall, we found that the Portsmouth Physiological and Operative Severity Score for the Enumeration of Mortality and Morbidity was most useful in higher-risk patients, with an optimal cut-off score of 40.5 per cent. |
| <u>Karabulut et al Turkey 2024</u><br>300 | To investigate whether these two risk calculators (ACS surgical risk calculator and P-POSSUM) accurately reflect actual mortality and morbidity outcomes when retrospectively assessed based on preoperative risk scores, to compare their predictive superiority against each other, and to assess their applicability to major hepatobiliary surgery<br>Retrospective | Patients undergoing major hepatobiliary surgeries between August 2016 and December 2021<br>Major hepatopancreaticobiliary surgery, pancreaticoduodenectomy, pylorus-preserving pancreaticoduodenectomy, total pancreatectomy, distal pancreatectomy, hepatectomy (right, left, partial, trisegmentectomy), and hepaticojejunostomy surgeries<br>(General surgery) | C-statistic, O/E, Brier | Morbidity | 0.672 (0.611-0.732) | 1.480 | 0.257 |  |
| <u>Sahiner et al</u> | The sensitivity and specificity of the | Patients who were admitted to the general | AUC | Morbidity | 0.734 (0.635-0.833) |  |  | The AUC result of e CRP/ALB was found |
| (2020)<br>Turkey<br>119 | CRP/ALB ratio in predicting postoperative morbidity and mortality in patients undergoing colorectal surgery were investigated by comparing it with P-POSSUM, Cr-POSSUM, and ACPGBICRC scores.<br>Retrospective | surgery outpatient clinic for colorectal malignancy, operated and followed up in the intensive care unit between January 2015 and November 2018<br>Surgery for colorectal cancer (Right hemicolectomy, Left hemicolectomy, Transverse colectomy, Anterior resection, Low anterior resection, Total colectomy).<br>(General surgery) |  |  |  |  |  | to be higher (0.817) than the value of 0.734 calculated for P-POSSUM. P-POSSUM appears to be more valuable than CRP/ALB in predicting morbidity |
| <u>Sevinyan et al</u><br><u>UK</u><br>153 | To evaluate the accuracy and reliability of the P-POSSUM and ACS-NSQIP surgical risk calculators in predicting postoperative complications in gynaecological–oncological robotic surgery<br>Retrospective | Patients who had undergone Da Vinci-assisted Robotic Surgery for suspected or confirmed gynaecological malignancies<br>(Gynaecology surgery) | AUC, Brier score | Morbidity | 0.551 | - | 0.183 | Morbidity was much better predicted by ACS-NSQIP than by P-POSSUM (AUC-0.608 vs. AUC-0.551) with the same result in mortality prediction (Brier score 0.0000). Moreover, a statistically significant overestimation of morbidity has been shown by the P-POSSUM calculator (p = 0.018). |

### 6.3 Information available on request

The protocol for this review, and a clinical summary are available on request.

## Data Availability

All data produced in the present study are available upon reasonable request to the authors

## Acknowledgements

The Public Health Wales team would like to thank Dr Claire Dunstan, Dr Meredith Graham, Dr Linda Warnock, Meredith Graham, the Clinical Implementation Network, and Praveena Pemmasani for their contributions during stakeholder meetings in guiding the focus of the review and interpretation of findings.

## APPENDIX

### 7.2 Appendix 1 Data extraction table of studies that include mortality

**Table 18.** Data extraction table of studies that include mortality.

| Authors | Country | No. of Participants/age group | Surgery/Category | Outcomes of interest | Accuracy Scores |
| --- | --- | --- | --- | --- | --- |
| **P-POSSUM** |  |  |  |  |  |
| Parmar et al. (2010) | UK | 344/mean (SD) age 70 (9.9) years. | Consecutive reconstructive vascular surgical procedures/Vascular category. | Major adverse cardiac event (MACE) within 30 days of surgery, defined as 1 or more of the following: myocardial infarction, coronary revascularization, sudden death, and left ventricular failure. | AUC 0.82 (95% CI: 0.73 to 0.91).<br>Good level of accuracy in predicting MACE after vascular surgery. |
| **ACS-NSQIP Surgical Risk Calculator tool** |  |  |  |  |  |
| Baker et al. (2018) | USA | 283/ above 18 years old. | An abdominal operation for oncology indications. These included open and laparoscopic operations in the following subcategories: 78 colorectal (27%), 46 pancreatic (16%), 35 gastro esophageal (12%), 30 liver (11%), 29 small bowel (10%), 29 retroperitoneal (10%), and 36 other (13%). General Category. | Composite<br>Grade 1 complications: minor (bedside procedures such as an nasogastric tube insertion)<br>Grade 2 complications: moderate (blood transfusions or need for total parenteral nutrition),<br>Grade 3 complications: severe (interventions without general anesthesia).<br>Grade 4 complications: severe (requiring interventions with general anesthesia).<br>Grade 5 complications: severe (involve organ system failure).<br>Grade 6 complications (post-operative death). | ACSNSQIP's AUC is reported for overall post-operative morbidity.<br>AUC 0.606<br>Poor level of accuracy in predicting complications across a wide variety of abdominal surgical procedures. |
| Suresh et al.(2019) | USA | 264/ average age 50.8 ± 12.6 years | Panniculectomy/ Plastic and Reconstructive | Postoperative complications were defined as major (return to the operating room within 30 days or death) or minor (surgical site infection, deep vein thrombosis/pulmonary embolism, wound dehiscence, use of postoperative antibiotics, postoperative blood transfusion, prolonged hospital length of stay, or seroma/hematoma formation). | AUC for major complication 0.514<br>Very poor level of accuracy in predicting major complication after Panniculectomy surgery.<br>AUC for any outcome 0.619. |
|  |  |  |  |  | Poor level of accuracy in predicting any complication after Panniculectomy surgery. |
| **ASA** |  |  |  |  |  |
| Parmar et al. (2010) | UK | 344/mean (SD) age 70 (9.9) years. | Consecutive reconstructive vascular surgical procedures/ Vascular Category. | MACE within 30 days of surgery, defined as 1 or more of the following: myocardial infarction, coronary revascularization, sudden death, and left ventricular failure. | AUC 0.67 (95% CI: 0.56 to 0.78).<br>Poor level of accuracy in predicting MACE after vascular surgery. |
| Huisman et al.(2014) | UK | 263/ aged≥70 years | Laparotomies (Colorectal cancer, gastric cancer and pancreatic cancer) and breast surgery/ General surgery. | Endpoint was morbidity during the first 30 days after surgery. Morbidity was registered using the Clavien-Dindo classification, a scale ranking severity of complications from 'any deviation from the normal post-operative course without the need for pharmacological treatment or surgical, endoscopic and radiological interventions' (grade one) to 'death of a patient' (grade five). Morbidity was dichotomized into minor (Clavien-Dindo grade one and two) and major complications (Clavien-Dindo grade three to five). | AUC is only for major complication. AUC 0.58, (95%CI:0.49 to 0.67). Very poor level of accuracy in predicting post-operative complications among onco-geriatric patients. |
| **RCRI** |  |  |  |  |  |
| Bryce et al. (2012) | UK | 106/66 to 77 years old. | Patients undergoing open elective abdominal aortic aneurysm (AAA)/ Vascular surgery. | MACE (major adverse cardiac event) defined as non-fatal myocardial infarction and cardiac death.<br>Major adverse cardiac event included myocardial infarction, cerebrovascular disease, congestive cardiac failure, chronic renal failure, chronic obstructive pulmonary disease, white cell count, prothrombin time. | AUC 0.525 (95% CI: 0.364 to 0.687)<br>Very poor predictive accuracy for MACE in those patients receiving open elective abdominal aortic aneurysm surgery. |
| Che et al. (2017) | China | 1202/aged 69.5±5.3 years | Patients who underwent noncardiac surgery. | Postoperative major cardiac event (PoMCE) within 30 days. PoMCEs defined as cardiac death, nonfatal myocardial infarction, nonfatal cardiac arrest, and heart failure. | AUC 0.53 (95% CI: 0.45 to 0.61).<br>Very poor predictive ability in identifying patients' cardiac risk among Chinese patients with CAD undergoing noncardiac surgery. |
| Cho et al. (2021) | South Korea | 256/68.7±12.1 years | Patients who underwent both a nuclear stress test and noncardiac surgery. | Postoperative major adverse cardiac event (MACE) as a composite of cardiac death, nonfatal myocardial infarction, and pulmonary edema. | AUC 0.612 (95% CI 0.549 to 0.672).<br>Poor predictive accuracy for MACE in those patients receiving non cardiac surgery. |
| Fronczek et al. (2019) | Poland | 870/aged≥45 years old | Patients undergoing in-hospital noncardiac surgery | Major cardiac complications defined as a composite of a nonfatal MI, a nonfatal cardiac arrest, or a cardiac death within 30 days after surgery. | C-statistic 0.60 (95% CI 0.54 to 0.65).<br>Poor predictive ability for major cardiac complications within 30 days of noncardiac surgery. |
| Palamuthusingam et al.(2024) | Australia and New Zealand | 5094/aged≥18 years old. | Patients receiving Chronic Kidney Replacement Therapy (KRT) who underwent elective abdominal surgery, /Urology. | Major adverse cardiovascular event (MACE), defined as nonfatal myocardial infarction, nonfatal stroke, non-fatal cardiac arrest and cardiovascular mortality at 30 days. | AUC 0.67 (95% CI 0.63 to 0.70).<br>Poor predictive ability for MACE at 30 days among the patients on chronic KRT undergoing elective surgery. |
| Parmar et al.(2010) | UK | 344/mean (SD) age 70 (9.9) years. | Consecutive reconstructive vascular surgical procedures/ Vascular surgery category. | MACE within 30 days of surgery, defined as 1 or more of the following: myocardial infarction, coronary revascularization, sudden death, and left ventricular failure. | AUC 0.68 (95% CI 0.57 to 0.83).<br>Poor ability in predicting MACE after vascular surgery. |
| Payne et al.(2013) | UK | 252/ aged≥70 years old. | Patients undergoing major vascular surgery (aortic surgery, infra-inguinal bypass surgery, amputation)/ Vascular surgery category. | Major adverse cardiac event (MACE), defined as non-fatal myocardial infarction and cardiac mortality. | AUC 0.538. Very poor predictive accuracy for MACE. |
| Roshanov et al.(2021) | 14 Countries, name not reported. | 35,815/ aged≥45 years old. | Major elective noncardiac surgery. | Major cardiac complication including death up to 30 days after surgery. Primary outcome as a composite of Myocardial injury after noncardiac surgery (MINS), myocardial infarction, nonfatal cardiac arrest, or cardiac death. secondary outcome as a composite of myocardial infarction, nonfatal cardiac arrest, or cardiac death. | Primary Outcome: C-statistic 0.65 (95% CI 0.62 to 0.68).<br>Secondary Outcome: C-statistics 0.69 (95% CI 0.65 to 0.72). Poor ability to predict both primary and secondary outcomes for noncardiac surgery. |
| Welten et al. (2007) | Netherlands | 2642/above 18 years. ≤55 years (396), 56 to 65 years (650), 66 to 75 years (1058) | Patients underwent open non-cardiac vascular surgical procedures/Vascular surgery. | Major adverse cardiac events (MACE) within 30 days after surgery. MACE within 30-days after surgery, was defined as cardiac death (which was defined | AUC 0.65. Poor predictive accuracy for MACE. |
|  |  | and above 75 years (538). |  | as any death with a cardiac complication as the primary or secondary cause, including deaths following myocardial infarction, cardiac arrhythmia and heart failure), myocardial infarction or coronary revascularization (PCI or CABG). |  |
| Wotton et al. (2013) | UK | 703/median age 68 years (range 14 to 89) | Patients underwent thoracotomy and lung resection/Cardiothoracic surgery. | Post-operative cardiac complication including death within 30 days. Post-operative major cardiac complications included pulmonary oedema, myocardial infarction (MI), ventricular fibrillation (VF) arrest, supraventricular arrhythmia, atrial fibrillation (AF) and mortality occurring within 30 days. | AUC 0.59 (95% CI 0.51 to 0.67). Very Poor discriminative ability for predicting post-operative cardiac complications. |
| Yao et al. (2021) | Australia | 1366/ aged≥45 years old. | Patients undergoing elective non-cardiac surgery. | Major adverse cardiac events (MACE) within 30 days of surgery, including myocardial infarction, pulmonary oedema, complete heart block or cardiac death. | AUC 0.73 (95% CI 0.60 to 0.86). Fair level of accuracy in predicting MACE. |
| Zou et al. (2024) | China | 453/aged over 60 years. | Elderly patients who underwent endovascular aortic aneurysm repair (EVAR)/ Vascular Surgery. | Major adverse cardiac and cerebrovascular events (MACCE). MACCE was defined as a composition of death, myocardial infarction, stroke, chronic cardiac failure and repeat revascularization. | AUC 0.588. Very poor level of accuracy in predicting MACCE. |

### 7.3 Appendix 2. Modified versions of surgical risk prediction tools

Below are references identified through our screening process that satisfied our eligibility criteria, but that used modified versions of included tools.

#### ARISCAT score (for Postoperative Pulmonary Complications)

ARISCAT plus predicted postoperative forced expiratory volume (ppoFEV1)

Zorrilla-Vaca, A., et al. (2023) Performance Comparison of Pulmonary Risk Scoring Systems in Lung Resection. *Journal of Cardiothoracic and Vascular Anesthesia* - Volume 37, Issue 9, pp.1734 EP – 1743.

#### ASA Physical Status/ ASA Classification

No modified tools identified meeting out eligibility criteria

#### Carlisle Risk/Carlisle Calculator

No modified tools identified meeting out eligibility criteria

#### CFS (Clinical Frailty Scale, also known as Rockwood)

No modified tools identified meeting out eligibility criteria

#### CPET (Cardiopulmonary exercise testing)

No modified tools identified meeting out eligibility criteria

#### DASI (Duke Activity Status Index)

Modified DASI

Li, M. H. G., Rosser, M., Blitz, J. (2024). A Retrospective Cohort Study Examining the Validation of the Modified Duke Activity Status Index in the Non-cardiac Surgical Population. Journal of PeriAnesthesia Nursing. S1089-9472(24)00361-7

#### NELA PRS (National Emergency Laparotomy Audit (Parsimonious Risk Score)

No modified tools identified meeting out eligibility criteria

#### NRS-2002 (Nutrition Risk Screening 2002)

No modified tools identified meeting out eligibility criteria

#### NSQIP (National Surgical Quality Improvement Program) universal surgical risk calculator)

Adapted model

Karabulut, A., Umman, V., Oral, G., et al. (2024) Assessing the effectiveness of ACS surgical risk calculator versus P-POSSUM in predicting mortality and morbidity for major hepatobiliary surgery: An observational study. Medicine - Volume 103, Issue 28, pp. 1-6.

#### P-POSSUM score (Physiological and Operative Severity Score for the enumeration of Mortality and Morbidity)

Modified/ improved POSSUM

Nie, Y., Li, Z., Su, T., et al. (2019) Application of Improved POSSUM Score Combined with Clavien-Dindo Classification in Predicting the Incidence of Severe Complications After Thoracoscopic Lung Surgery. *Indian Journal of Surgery* - Volume 82, Issue 6, pp. 1031-1037.

Adapted model

Karabulut, A., Umman, V., Oral, G., et al. (2024) Assessing the effectiveness of ACS surgical risk calculator versus P-POSSUM in predicting mortality and morbidity for major hepatobiliary surgery: An observational study. Medicine - Volume 103, Issue 28, pp. 1-6.

Cr-POSSUM

Şahiner, Y., Yıldırım, M. B. (2020). Can the c-reactive protein-to-plasma albumin ratio be an alternative scoring to show mortality and morbidity in patients with colorectal cancer? Ulusal Travma ve Acil Cerrahi Dergisi - Volume 26, Issue 4, pp. 580-585.

Prabakaran, V., Thangaraju, T., Mathew, A. C., et al. (2019). *Indian Journal of Surgical Oncology* - Volume 10, Issue 1, pp. 174 EP – 179.

E-POSSUM

Kim, S. Y., Kim, J. H., Chin, H., et al. (2020). Prediction of postoperative mortality and morbidity in octogenarians with gastric cancer - Comparison of P-POSSUM, O-POSSUM, and E-POSSUM: A retrospective single-center cohort study. International Journal of Surgery - Volume 77, Issue 0, pp. 64 EP – 68.

O-POSSUM

He, H., Liu, Y., Liu, X., et al. (2023). Evaluation of different scoring systems in the prediction of complications, morbidity, and mortality after laparoscopic radical gastrectomy. World Journal of Surgical Oncology - Volume 21, Issue 1.

Kim, S. Y., Kim, J. H., Chin, H., et al. (2020). Prediction of postoperative mortality and morbidity in octogenarians with gastric cancer - Comparison of P-POSSUM, O-POSSUM, and E-POSSUM: A retrospective single-center cohort study. International Journal of Surgery - Volume 77, Issue 0, pp. 64 EP – 68.

PD-POSSUM

Zhang, Z. L., Chen, L., Peng, L., et al. (2020). A newly improved POSSUM scoring system for prediction of morbidity in patients with pancreaticoduodenectomy. *Translational Cancer Research* - Volume 9, Issue 9, pp. 5517 EP – 5527.

#### PONV (Apfel Score for Postoperative Nausea and Vomiting)

No modified tools identified meeting out eligibility criteria

#### RCRI (Revised Cardiac Risk Index for Pre-Operative Risk)

Modified RCRI

Ackland, G. L., Harris, S., Ziabari, Y., et al. (2020). Revised cardiac risk index and postoperative morbidity after elective orthopaedic surgery: a prospective cohort study. British journal of anaesthesia - Volume 105, Issue 6, pp. 744-52 - published 2010-01-01.

Pandey, A., Sood, A., Sammon, J. D., et al. (2015). Effect of preoperative angina pectoris on cardiac outcomes in patients with previous myocardial infarction undergoing major noncardiac surgery (data from ACS-NSQIP). *American Journal of Cardiology* - Volume 115, Issue 8, pp. 1080-1084.

Schmidt, G., Frieling, N., Schneck, E., et al. (2024). Comparison of preoperative NT-proBNP and simple cardiac risk scores for predicting postoperative morbidity after non-cardiac surgery with intermediate or high surgical risk. *Perioperative Medicine* - Volume 13, Issue 1, pp. 44.

Reconstructed RCRI

Cohn, S. L., Fernandez Ros, N. (2018). Comparison of 4 Cardiac Risk Calculators in Predicting Postoperative Cardiac Complications After Noncardiac Operations. American Journal of Cardiology - Volume 121, Issue 1, pp. 125 EP – 130.

Davis, C., Tait, G., Carroll, J., et al. (2013). The Revised Cardiac Risk Index in the new millennium: a single-centre prospective cohort re-evaluation of the original variables in 9,519 consecutive elective surgical patients. Canadian journal of anaesthesia - Volume 60, Issue 9, pp. 855-63.

#### ThRCRI

No modified tools identified meeting out eligibility criteria

#### SORT (Surgical Outcome Risk Tool)

No modified tools identified meeting out eligibility criteria

### 7.4 Appendix 3. Breakdown of all outcomes reported across studies by tool and surgical specialty

Below is a breakdown of the different outcomes reported for ACS NSQIP, P-POSSUM, the RCRI and the ASA classification system. Where more than one study was reported for a particular surgical specialty, the findings have also been tabulated below. Please note some of the composite complications may include mortality as the composites were not always clearly defined within the studies.

**Table 19.**
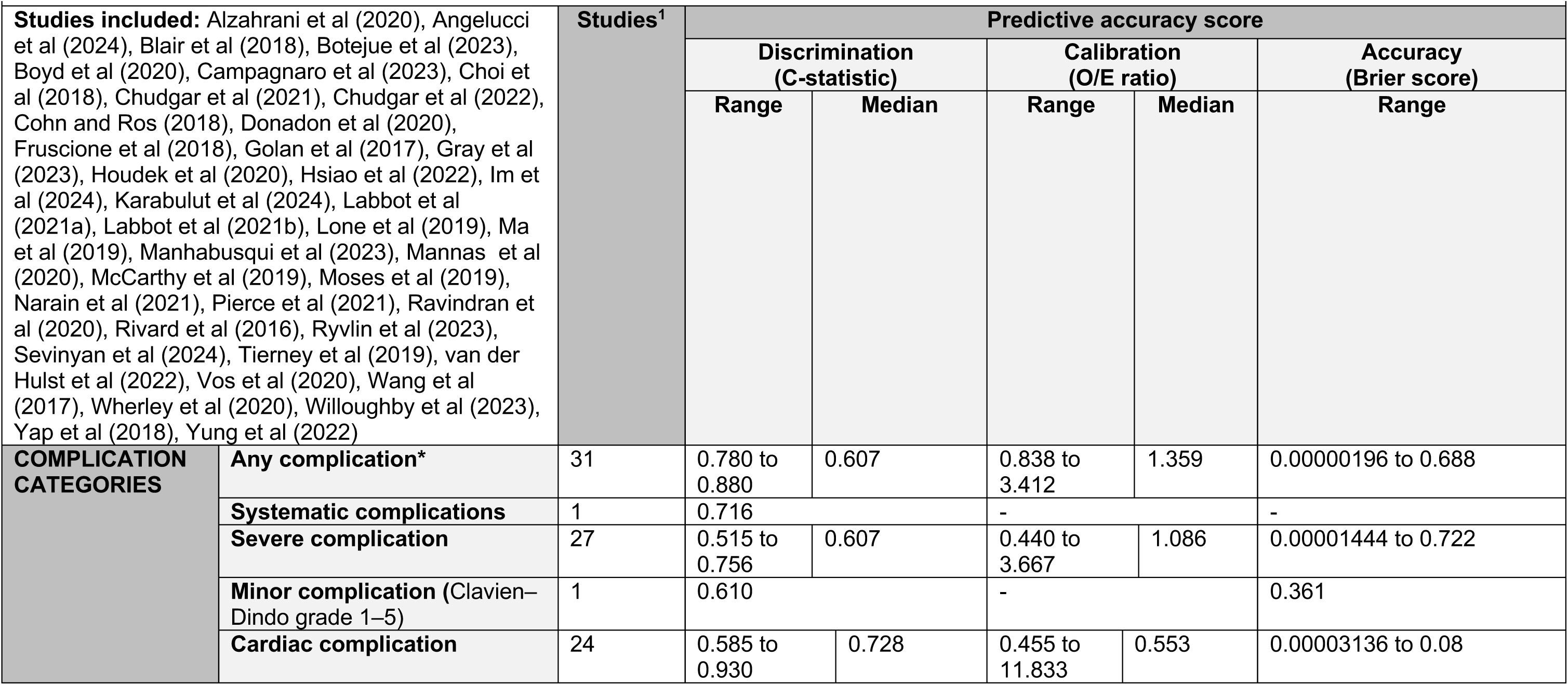
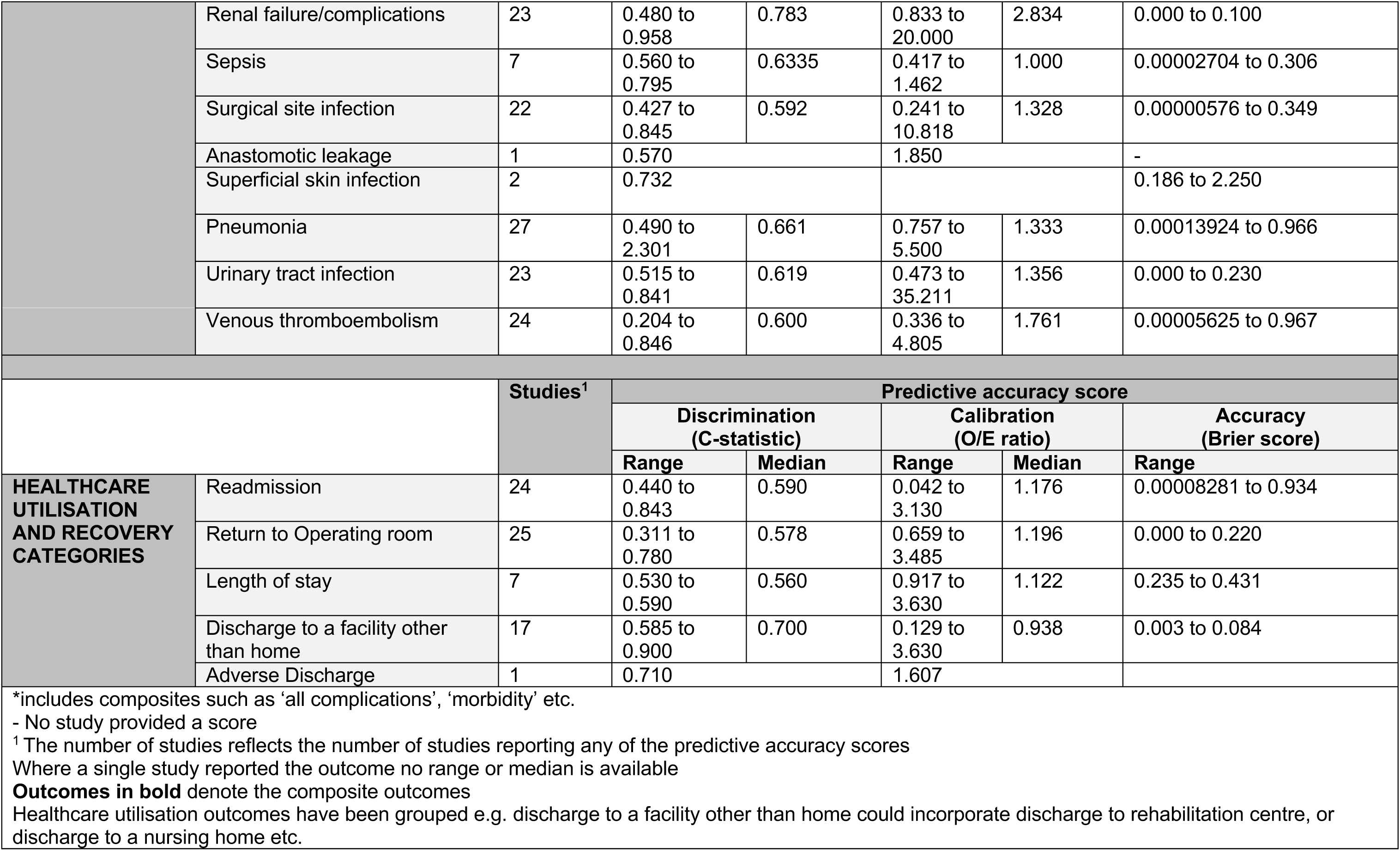
ACS NSQIP (40 studies) all studies.

| Table 19 – ACS NSQIP (40 studies) all studies |  |  |  |  |  |  |  |
| --- | --- | --- | --- | --- | --- | --- | --- |
| **Studies included:** Alzahrani et al (2020), Angelucci et al (2024), Blair et al (2018), Botejue et al (2023), Boyd et al (2020), Campagnaro et al (2023), Choi et al (2018), Chudgar et al (2021), Chudgar et al (2022), Cohn and Ros (2018), Donadon et al (2020), Fruscione et al (2018), Golan et al (2017), Gray et al (2023), Houdek et al (2020), Hsiao et al (2022), Im et al (2024), Karabulut et al (2024), Labbot et al (2021a), Labbot et al (2021b), Lone et al (2019), Ma et al (2019), Manhabusqui et al (2023), Mannas et al (2020), McCarthy et al (2019), Moses et al (2019), Narain et al (2021), Pierce et al (2021), Ravindran et al (2020), Rivard et al (2016), Ryvlin et al (2023), Sevinyan et al (2024), Tierney et al (2019), van der Hulst et al (2022), Vos et al (2020), Wang et al (2017), Wherley et al (2020), Willoughby et al (2023), Yap et al (2018), Yung et al (2022) |  | **Studies**<sup>1</sup> | **Predictive accuracy score** |  |  |  |  |
|  |  |  | **Discrimination (C-statistic)** |  | **Calibration (O/E ratio)** |  | **Accuracy (Brier score)** |
|  |  |  | **Range** | **Median** | **Range** | **Median** | **Range** |
| **COMPLICATION CATEGORIES** | **Any complication\*** | 31 | 0.780 to 0.880 | 0.607 | 0.838 to 3.412 | 1.359 | 0.00000196 to 0.688 |
|  | **Systematic complications** | 1 | 0.716 |  | - |  | - |
|  | **Severe complication** | 27 | 0.515 to 0.756 | 0.607 | 0.440 to 3.667 | 1.086 | 0.00001444 to 0.722 |
|  | **Minor complication (Clavien–Dindo grade 1–5)** | 1 | 0.610 |  | - |  | 0.361 |
|  | **Cardiac complication** | 24 | 0.585 to 0.930 | 0.728 | 0.455 to 11.833 | 0.553 | 0.00003136 to 0.08 |
|  | Renal failure/complications | 23 | 0.480 to 0.958 | 0.783 | 0.833 to 20.000 | 2.834 | 0.000 to 0.100 |
|  | Sepsis | 7 | 0.560 to 0.795 | 0.6335 | 0.417 to 1.462 | 1.000 | 0.00002704 to 0.306 |
|  | Surgical site infection | 22 | 0.427 to 0.845 | 0.592 | 0.241 to 10.818 | 1.328 | 0.00000576 to 0.349 |
|  | Anastomotic leakage | 1 | 0.570 |  | 1.850 |  | - |
|  | Superficial skin infection | 2 | 0.732 |  |  |  | 0.186 to 2.250 |
|  | Pneumonia | 27 | 0.490 to 2.301 | 0.661 | 0.757 to 5.500 | 1.333 | 0.00013924 to 0.966 |
|  | Urinary tract infection | 23 | 0.515 to 0.841 | 0.619 | 0.473 to 35.211 | 1.356 | 0.000 to 0.230 |
|  | Venous thromboembolism | 24 | 0.204 to 0.846 | 0.600 | 0.336 to 4.805 | 1.761 | 0.00005625 to 0.967 |
|  |  | Studies<sup>1</sup> | Predictive accuracy score |  |  |  |  |
|  |  |  | Discrimination (C-statistic) |  | Calibration (O/E ratio) |  | Accuracy (Brier score) |
|  |  |  | Range | Median | Range | Median | Range |
| HEALTHCARE UTILISATION AND RECOVERY CATEGORIES | Readmission | 24 | 0.440 to 0.843 | 0.590 | 0.042 to 3.130 | 1.176 | 0.00008281 to 0.934 |
|  | Return to Operating room | 25 | 0.311 to 0.780 | 0.578 | 0.659 to 3.485 | 1.196 | 0.000 to 0.220 |
|  | Length of stay | 7 | 0.530 to 0.590 | 0.560 | 0.917 to 3.630 | 1.122 | 0.235 to 0.431 |
|  | Discharge to a facility other than home | 17 | 0.585 to 0.900 | 0.700 | 0.129 to 3.630 | 0.938 | 0.003 to 0.084 |
|  | Adverse Discharge | 1 | 0.710 |  | 1.607 |  |  |
| *includes composites such as 'all complications', 'morbidity' etc.<br>- No study provided a score<br><sup>1</sup> The number of studies reflects the number of studies reporting any of the predictive accuracy scores<br>Where a single study reported the outcome no range or median is available<br>**Outcomes in bold** denote the composite outcomes<br>Healthcare utilisation outcomes have been grouped e.g. discharge to a facility other than home could incorporate discharge to rehabilitation centre, or discharge to a nursing home etc. |  |  |  |  |  |  |  |

**Table 20.**
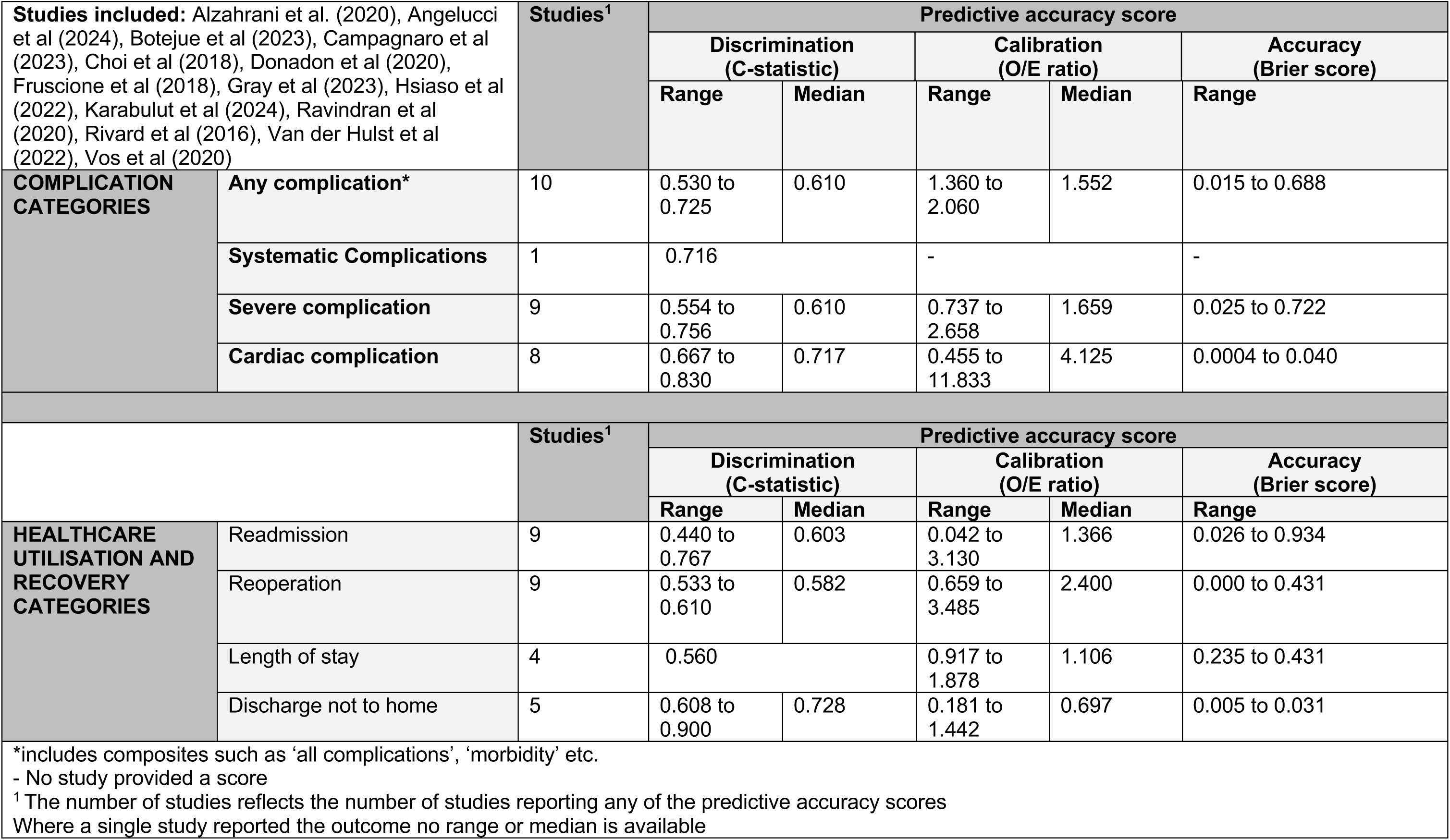
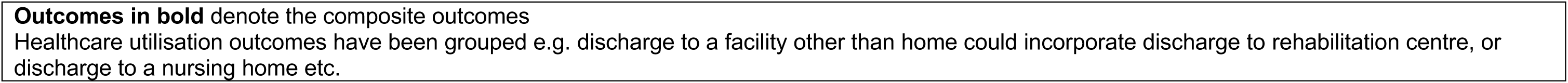
ACS NSQIP (14 studies) General Surgery.

| Table 20 – ACS NSQIP (14 studies) General Surgery |  |  |  |  |  |  |  |
| --- | --- | --- | --- | --- | --- | --- | --- |
| **Studies included:** Alzahrani et al. (2020), Angelucci et al (2024), Botejue et al (2023), Campagnaro et al (2023), Choi et al (2018), Donadon et al (2020), Fruscione et al (2018), Gray et al (2023), Hsiao et al (2022), Karabulut et al (2024), Ravindran et al (2020), Rivard et al (2016), Van der Hulst et al (2022), Vos et al (2020) |  | Studies<sup>1</sup> | Predictive accuracy score |  |  |  |  |
|  |  |  | Discrimination (C-statistic) |  | Calibration (O/E ratio) |  | Accuracy (Brier score) |
|  |  |  | Range | Median | Range | Median | Range |
| **COMPLICATION CATEGORIES** | **Any complication\*** | 10 | 0.530 to 0.725 | 0.610 | 1.360 to 2.060 | 1.552 | 0.015 to 0.688 |
|  | **Systematic Complications** | 1 | 0.716 |  | - |  | - |
|  | **Severe complication** | 9 | 0.554 to 0.756 | 0.610 | 0.737 to 2.658 | 1.659 | 0.025 to 0.722 |
|  | **Cardiac complication** | 8 | 0.667 to 0.830 | 0.717 | 0.455 to 11.833 | 4.125 | 0.0004 to 0.040 |
|  |  | Studies<sup>1</sup> | Predictive accuracy score |  |  |  |  |
|  |  |  | Discrimination (C-statistic) |  | Calibration (O/E ratio) |  | Accuracy (Brier score) |
|  |  |  | Range | Median | Range | Median | Range |
| **HEALTHCARE UTILISATION AND RECOVERY CATEGORIES** | Readmission | 9 | 0.440 to 0.767 | 0.603 | 0.042 to 3.130 | 1.366 | 0.026 to 0.934 |
|  | Reoperation | 9 | 0.533 to 0.610 | 0.582 | 0.659 to 3.485 | 2.400 | 0.000 to 0.431 |
|  | Length of stay | 4 | 0.560 |  | 0.917 to 1.878 | 1.106 | 0.235 to 0.431 |
|  | Discharge not to home | 5 | 0.608 to 0.900 | 0.728 | 0.181 to 1.442 | 0.697 | 0.005 to 0.031 |
| *includes composites such as 'all complications', 'morbidity' etc.<br>- No study provided a score<br><sup>1</sup> The number of studies reflects the number of studies reporting any of the predictive accuracy scores<br>Where a single study reported the outcome no range or median is available |  |  |  |  |  |  |  |

**Outcomes in bold** denote the composite outcomes Healthcare utilisation outcomes have been grouped e.g. discharge to a facility other than home could incorporate discharge to rehabilitation centre, or discharge to a nursing home etc.

**Table 21.**
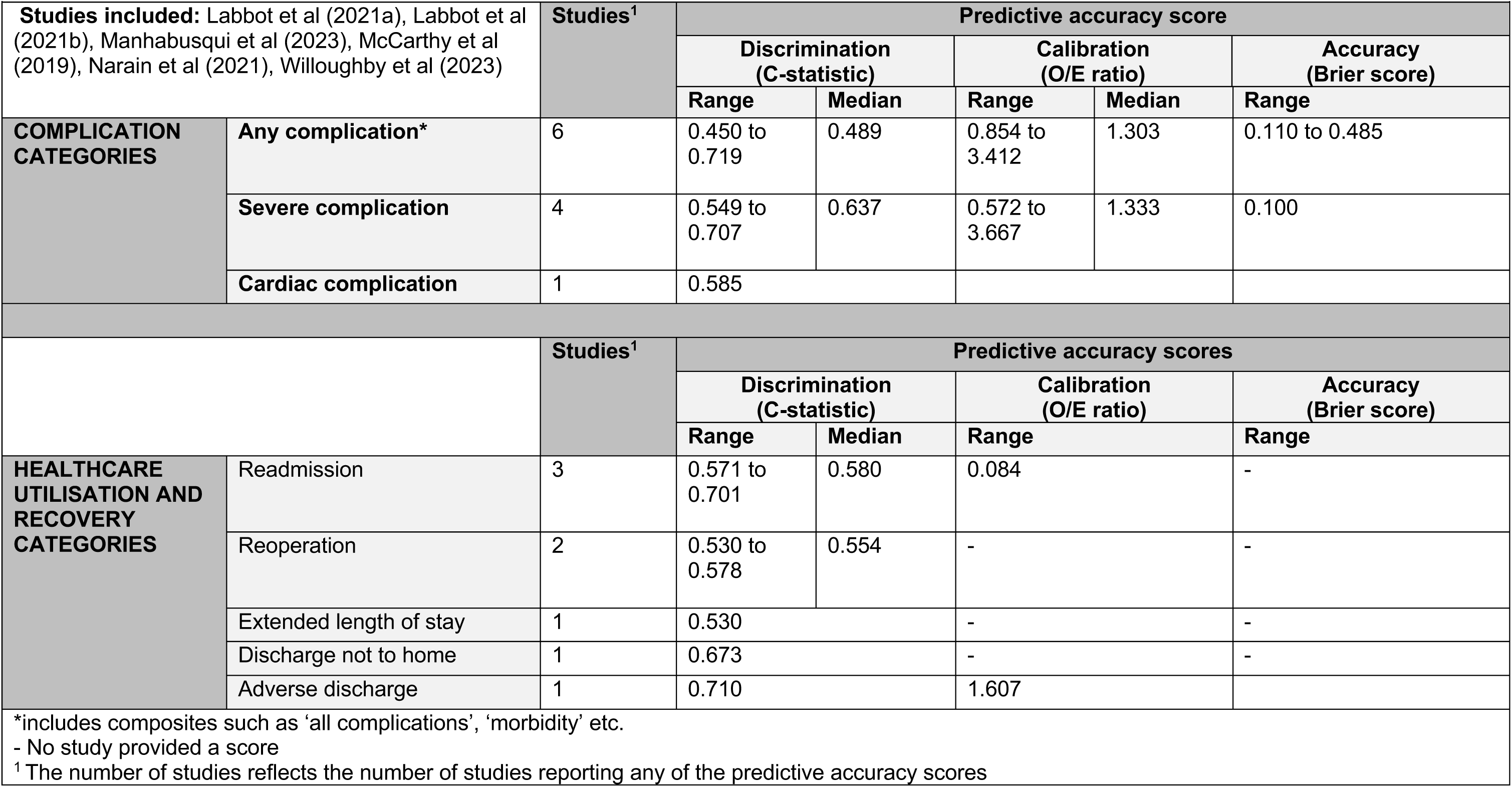
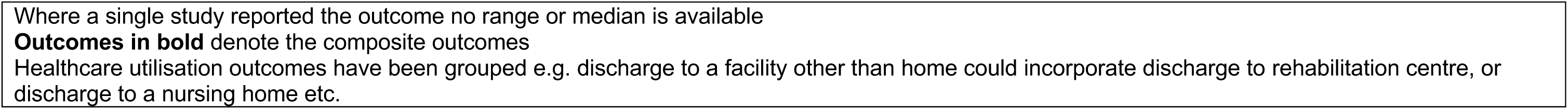
ACS NSQIP (6 studies) Orthopaedic Surgery.

| Studies included: Labbot et al (2021a), Labbot et al (2021b), Manhabusqui et al (2023), McCarthy et al (2019), Narain et al (2021), Willoughby et al (2023) |  | Studies<sup>1</sup> | Predictive accuracy score |  |  |  |  |
| --- | --- | --- | --- | --- | --- | --- | --- |
|  |  |  | Discrimination (C-statistic) |  | Calibration (O/E ratio) |  | Accuracy (Brier score) |
|  |  |  | Range | Median | Range | Median | Range |
| COMPLICATION CATEGORIES | Any complication* | 6 | 0.450 to 0.719 | 0.489 | 0.854 to 3.412 | 1.303 | 0.110 to 0.485 |
|  | Severe complication | 4 | 0.549 to 0.707 | 0.637 | 0.572 to 3.667 | 1.333 | 0.100 |
|  | Cardiac complication | 1 | 0.585 |  |  |  |  |
|  |  | Studies<sup>1</sup> | Predictive accuracy scores |  |  |  |  |
|  |  |  | Discrimination (C-statistic) |  | Calibration (O/E ratio) |  | Accuracy (Brier score) |
|  |  |  | Range | Median | Range |  | Range |
| HEALTHCARE UTILISATION AND RECOVERY CATEGORIES | Readmission | 3 | 0.571 to 0.701 | 0.580 | 0.084 |  | - |
|  | Reoperation | 2 | 0.530 to 0.578 | 0.554 | - |  | - |
|  | Extended length of stay | 1 | 0.530 |  | - |  | - |
|  | Discharge not to home | 1 | 0.673 |  | - |  | - |
|  | Adverse discharge | 1 | 0.710 |  | 1.607 |  |  |
| *includes composites such as ‘all complications’, ‘morbidity’ etc.<br>- No study provided a score<br><sup>1</sup> The number of studies reflects the number of studies reporting any of the predictive accuracy scores |  |  |  |  |  |  |  |

Where a single study reported the outcome no range or median is available
**Outcomes in bold** denote the composite outcomes
Healthcare utilisation outcomes have been grouped e.g. discharge to a facility other than home could incorporate discharge to rehabilitation centre, or discharge to a nursing home etc.

**Table 22.**
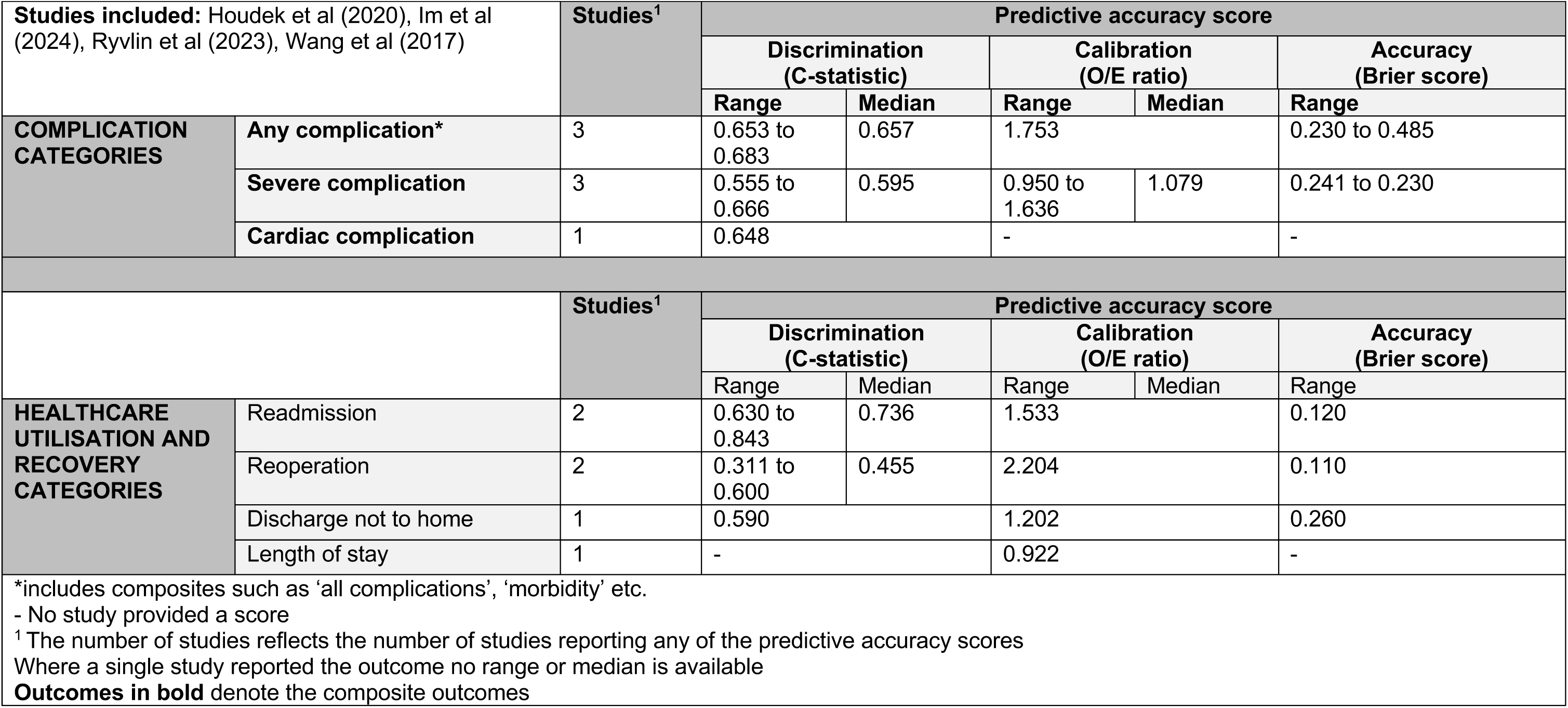
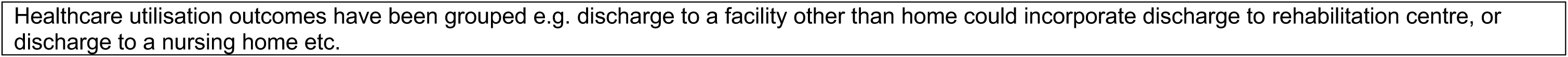
ACS NSQIP (4 studies) Neurosurgery.

| Table 22 – ACS NSQIP (4 studies) Neurosurgery |  |  |  |  |  |  |  |
| --- | --- | --- | --- | --- | --- | --- | --- |
| **Studies included:** Houdek et al (2020), Im et al (2024), Rylvlin et al (2023), Wang et al (2017) |  | **Studies<sup>1</sup>** | **Predictive accuracy score** |  |  |  |  |
|  |  |  | **Discrimination (C-statistic)** |  | **Calibration (O/E ratio)** |  | **Accuracy (Brier score)** |
|  |  |  | **Range** | **Median** | **Range** | **Median** | **Range** |
| **COMPLICATION CATEGORIES** | **Any complication\*** | 3 | 0.653 to 0.683 | 0.657 | 1.753 |  | 0.230 to 0.485 |
|  | **Severe complication** | 3 | 0.555 to 0.666 | 0.595 | 0.950 to 1.636 | 1.079 | 0.241 to 0.230 |
|  | **Cardiac complication** | 1 | 0.648 |  | - |  | - |
|  |  | **Studies<sup>1</sup>** | **Predictive accuracy score** |  |  |  |  |
|  |  |  | **Discrimination (C-statistic)** |  | **Calibration (O/E ratio)** |  | **Accuracy (Brier score)** |
|  |  |  | **Range** | **Median** | **Range** | **Median** | **Range** |
| **HEALTHCARE UTILISATION AND RECOVERY CATEGORIES** | Readmission | 2 | 0.630 to 0.843 | 0.736 | 1.533 |  | 0.120 |
|  | Reoperation | 2 | 0.311 to 0.600 | 0.455 | 2.204 |  | 0.110 |
|  | Discharge not to home | 1 | 0.590 |  | 1.202 |  | 0.260 |
|  | Length of stay | 1 | - |  | 0.922 |  | - |
| *includes composites such as 'all complications', 'morbidity' etc.<br>- No study provided a score<br><sup>1</sup> The number of studies reflects the number of studies reporting any of the predictive accuracy scores<br>Where a single study reported the outcome no range or median is available<br>**Outcomes in bold** denote the composite outcomes |  |  |  |  |  |  |  |

Healthcare utilisation outcomes have been grouped e.g. discharge to a facility other than home could incorporate discharge to rehabilitation centre, or discharge to a nursing home etc.

**Table 23.** ACS NSQIP (4 studies) Urology.

| Studies included: Blair et al (2018), Golan et al (2017), Lone et al (2019), Mannas et al (2020) |  | Studies<sup>1</sup> | Predictive accuracy score |  |  |  |  |
| --- | --- | --- | --- | --- | --- | --- | --- |
|  |  |  | Discrimination (C statistic) |  | Calibration (O/E ratio) |  | Accuracy (Brier score) |
|  |  |  | Range | Median | Range | Median | Range |
| COMPLICATION CATEGORIES | Any complication* | 4 | 0.500 to 0.610 | 0.590 | 1.108 to 1.833 | 1.501 | 0.189 to 0.361 |
|  | Severe complication | 4 | 0.530 to 06.00 | 0.565 | 0.791to 1.365 | 0.938 | 0.121 to 0.240 |
|  | Cardiac complication | 4 | 0.590 to 0.800 | 0.690 | 0.955 to 2.00 | 1.381 | 0.020 to 0.38 |
|  |  | Studies<sup>1</sup> | Predictive accuracy score |  |  |  |  |
|  |  |  | Discrimination (C statistic) |  | Calibration (O/E ratio) |  | Accuracy (Brier score) |
|  |  |  | Range | Median | Range | Median | Range |
| HEALTHCARE UTILISATION AND RECOVERY CATEGORIES | Readmission | 3 | 0.520 to 0.600 | 0.550 | 0.667 to 1.296 | 1.191 | 0.111 to 0.180 |
|  | Reoperation | 4 | 0.520 to 0.590 | 0.580 | 0.841 to 1.400 | 1.260 | 0.034 to 0.67 |
|  | Discharge not to home | 3 | 0.690 to 0.750 | 0.720 | 0.938 to 3.630 | 0.990 | 0.034 to 0.090 |
|  | Length of stay | 2 | 0.590 |  | 1.148 to 3.630 | 2.389 | 0.419 |

\*includes composites such as ‘all complications’, ‘morbidity’ etc. - No study provided a score
<sup>1</sup> The number of studies reflects the number of studies reporting any of the predictive accuracy scores Where a single study reported the outcome no range or median is available
**Outcomes in bold** denote the composite outcomes
Healthcare utilisation outcomes have been grouped e.g. discharge to a facility other than home could incorporate discharge to rehabilitation centre, or discharge to a nursing home etc.

**Table 24.**
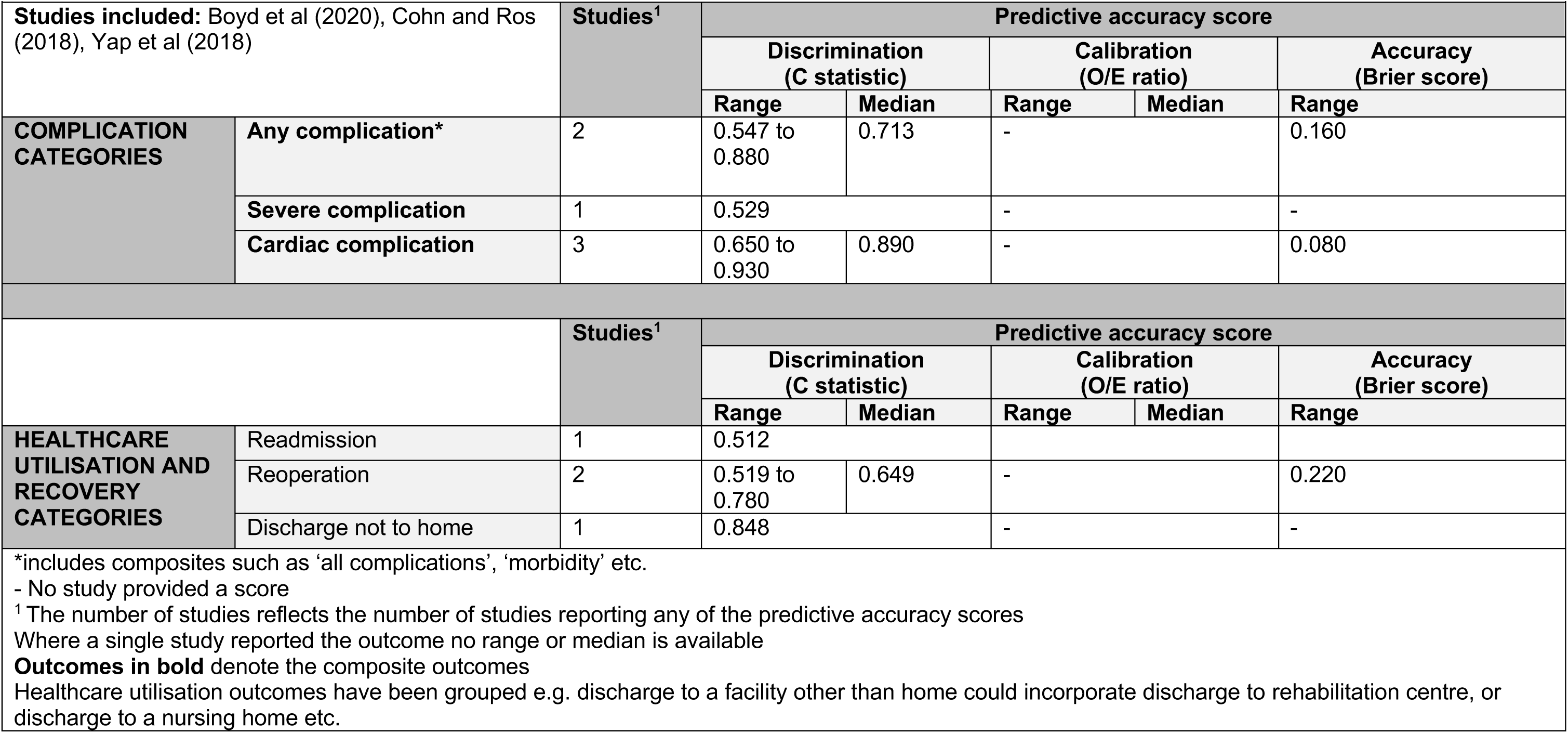
ACS NSQIP (3 studies) mixed Surgery.

| Table 24 – ACS NSQIP (3 studies) mixed Surgery |  |  |  |  |  |  |  |
| --- | --- | --- | --- | --- | --- | --- | --- |
| **Studies included:** Boyd et al (2020), Cohn and Ros (2018), Yap et al (2018) |  | **Studies<sup>1</sup>** | **Predictive accuracy score** |  |  |  |  |
|  |  |  | **Discrimination (C statistic)** |  | **Calibration (O/E ratio)** |  | **Accuracy (Brier score)** |
|  |  |  | **Range** | **Median** | **Range** | **Median** | **Range** |
| **COMPLICATION CATEGORIES** | **Any complication\*** | 2 | 0.547 to 0.880 | 0.713 | - |  | 0.160 |
|  | **Severe complication** | 1 | 0.529 |  | - |  | - |
|  | **Cardiac complication** | 3 | 0.650 to 0.930 | 0.890 | - |  | 0.080 |
|  |  | **Studies<sup>1</sup>** | **Predictive accuracy score** |  |  |  |  |
|  |  |  | **Discrimination (C statistic)** |  | **Calibration (O/E ratio)** |  | **Accuracy (Brier score)** |
|  |  |  | **Range** | **Median** | **Range** | **Median** | **Range** |
| **HEALTHCARE UTILISATION AND RECOVERY CATEGORIES** | Readmission | 1 | 0.512 |  |  |  |  |
|  | Reoperation | 2 | 0.519 to 0.780 | 0.649 | - |  | 0.220 |
|  | Discharge not to home | 1 | 0.848 |  | - |  | - |
| *includes composites such as ‘all complications’, ‘morbidity’ etc.<br>- No study provided a score<br><sup>1</sup> The number of studies reflects the number of studies reporting any of the predictive accuracy scores<br>Where a single study reported the outcome no range or median is available<br>**Outcomes in bold** denote the composite outcomes<br>Healthcare utilisation outcomes have been grouped e.g. discharge to a facility other than home could incorporate discharge to rehabilitation centre, or discharge to a nursing home etc. |  |  |  |  |  |  |  |

**Table 25.**
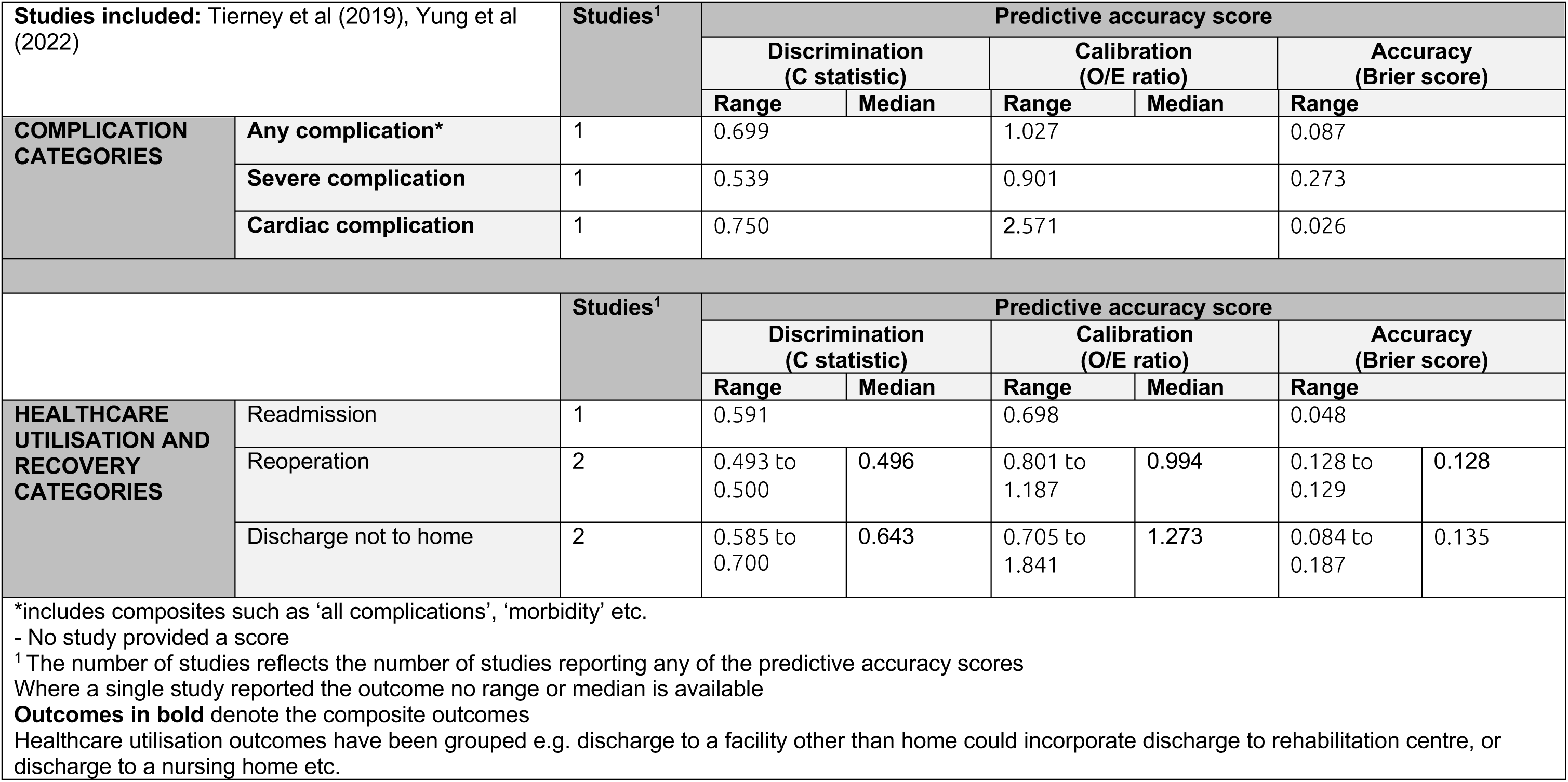
ACS NSQIP (2 studies) Plastic.

| Studies included: Tierney et al (2019), Yung et al (2022) |  | Studies<sup>1</sup> | Predictive accuracy score |  |  |  |  |  |
| --- | --- | --- | --- | --- | --- | --- | --- | --- |
|  |  |  | Discrimination (C statistic) |  | Calibration (O/E ratio) |  | Accuracy (Brier score) |  |
|  |  |  | Range | Median | Range | Median | Range |  |
| COMPLICATION CATEGORIES | Any complication* | 1 | 0.699 |  | 1.027 |  | 0.087 |  |
|  | Severe complication | 1 | 0.539 |  | 0.901 |  | 0.273 |  |
|  | Cardiac complication | 1 | 0.750 |  | 2.571 |  | 0.026 |  |
|  |  | Studies<sup>1</sup> | Predictive accuracy score |  |  |  |  |  |
|  |  |  | Discrimination (C statistic) |  | Calibration (O/E ratio) |  | Accuracy (Brier score) |  |
|  |  |  | Range | Median | Range | Median | Range |  |
| HEALTHCARE UTILISATION AND RECOVERY CATEGORIES | Readmission | 1 | 0.591 |  | 0.698 |  | 0.048 |  |
|  | Reoperation | 2 | 0.493 to 0.500 | 0.496 | 0.801 to 1.187 | 0.994 | 0.128 to 0.129 | 0.128 |
|  | Discharge not to home | 2 | 0.585 to 0.700 | 0.643 | 0.705 to 1.841 | 1.273 | 0.084 to 0.187 | 0.135 |
| *includes composites such as ‘all complications’, ‘morbidity’ etc.<br>- No study provided a score<br><sup>1</sup> The number of studies reflects the number of studies reporting any of the predictive accuracy scores<br>Where a single study reported the outcome no range or median is available<br>**Outcomes in bold** denote the composite outcomes<br>Healthcare utilisation outcomes have been grouped e.g. discharge to a facility other than home could incorporate discharge to rehabilitation centre, or discharge to a nursing home etc. |  |  |  |  |  |  |  |  |

**Table 26.** ACS NSQIP (2 studies) Vascular.

| Table 26 – ACS NSQIP (2 studies) Vascular |  |  |  |  |
| --- | --- | --- | --- | --- |
| Studies included: Ma et al (2018), Moses et al (2019) | Studies<sup>1</sup> | Predictive accuracy score |  |  |
|  |  | Discrimination (C statistic) | Calibration (O/E ratio) | Accuracy (Brier score) |

|  |  |  | Range | Median | Range | Median | Range |
| --- | --- | --- | --- | --- | --- | --- | --- |
| COMPLICATION CATEGORIES | Any complication* | 1 | 0.599 |  | 1.552 |  | - |
|  | Severe complication | 1 | 0.588 |  | 1.618 |  | - |
|  | Cardiac complication | 2 | 0.601 |  | 1.605 to 1.795 | 1.700 | - |
|  |  | Studies<sup>1</sup> | Predictive accuracy score |  |  |  |  |
|  |  |  | Discrimination (C statistic) |  | Calibration (O/E ratio) |  | Accuracy (Brier score) |
|  |  |  | Range | Median | Range | Median | Range |
| HEALTHCARE UTILISATION AND RECOVERY CATEGORIES | Reoperation | 1 | 0.535 |  | 1.884 |  | - |
|  | Discharge not to home | 1 | 0.693 |  | 1.058 |  | - |

\*includes composites such as ‘all complications’, ‘morbidity’ etc. - No study provided a score <sup>1</sup> The number of studies reflects the number of studies reporting any of the predictive accuracy scores Where a single study reported the outcome no range or median is available **Outcomes in bold** denote the composite outcomes Healthcare utilisation outcomes have been grouped e.g. discharge to a facility other than home could incorporate discharge to rehabilitation centre, or discharge to a nursing home etc.

#### P-POSSUM

**Table 27.** P-POSSUM (7 studies) all studies.

| Table 27 – P-POSSUM (7 studies) all studies |  |  |  |  |  |  |  |
| --- | --- | --- | --- | --- | --- | --- | --- |
| Studies included: Angelucci et al (2024); Bodea et al (2018); He et al (2023); Jones et al (2022); Karabulut et al (2024); Sahiner et al (2020); Sevinyan et al (2024) |  | Studies<sup>1</sup> | Predictive accuracy scores |  |  |  |  |
|  |  |  | Discrimination (C statistic) |  | Calibration (O/E ratio) |  | Accuracy (Brier score) |
|  |  |  | Range | Median | Range | Median | Range |
| COMPLICATION CATEGORIES | Any complication* | 7 | 0.551 to 0.781 | 0.567 | 1.48 to 1.66 | 1.57 | 0.186 to 0.257 |
|  | Severe complication | 1 | 0.63 |  | - |  | 0.205 |
|  | Pancreaticoduodenectomy specific complications | 1 | 0.64 |  | 0.89 |  | - |
| *includes composites such as 'all complications', 'morbidity' etc.<br>- No study provided a score<br><sup>1</sup> The number of studies reflects the number of studies reporting any of the predictive accuracy scores<br>Where a single study reported the outcome no range or median is available<br>**Outcomes in bold** denote the composite outcomes |  |  |  |  |  |  |  |

**Table 28.**
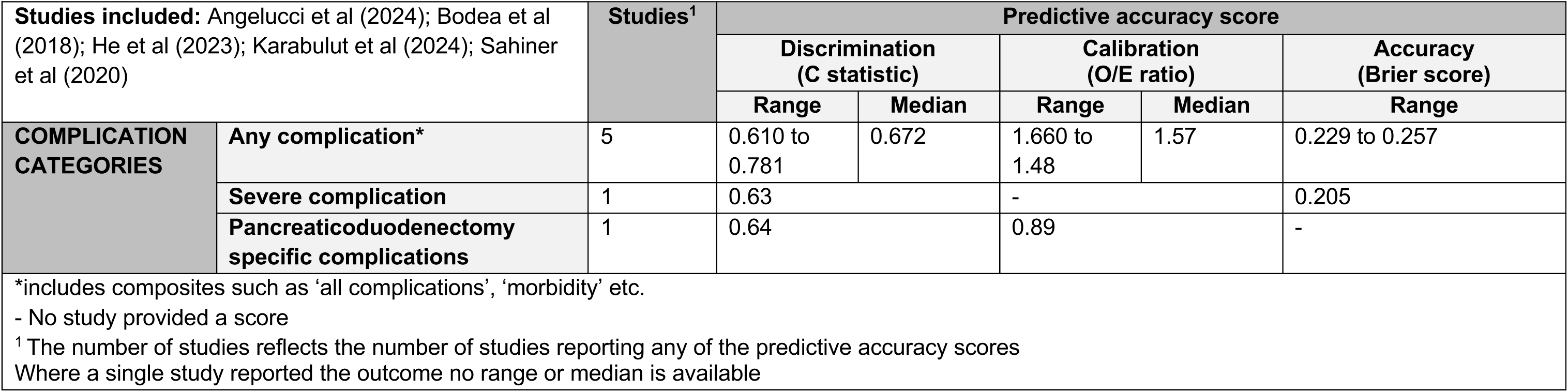
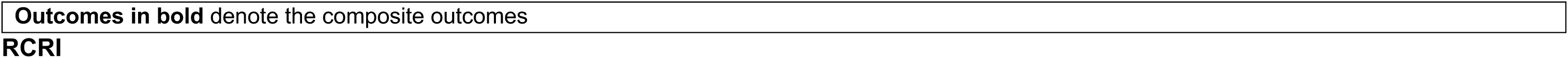
P-POSSUM (5 studies) General Surgery.

#### RCRI

**Table 29.**
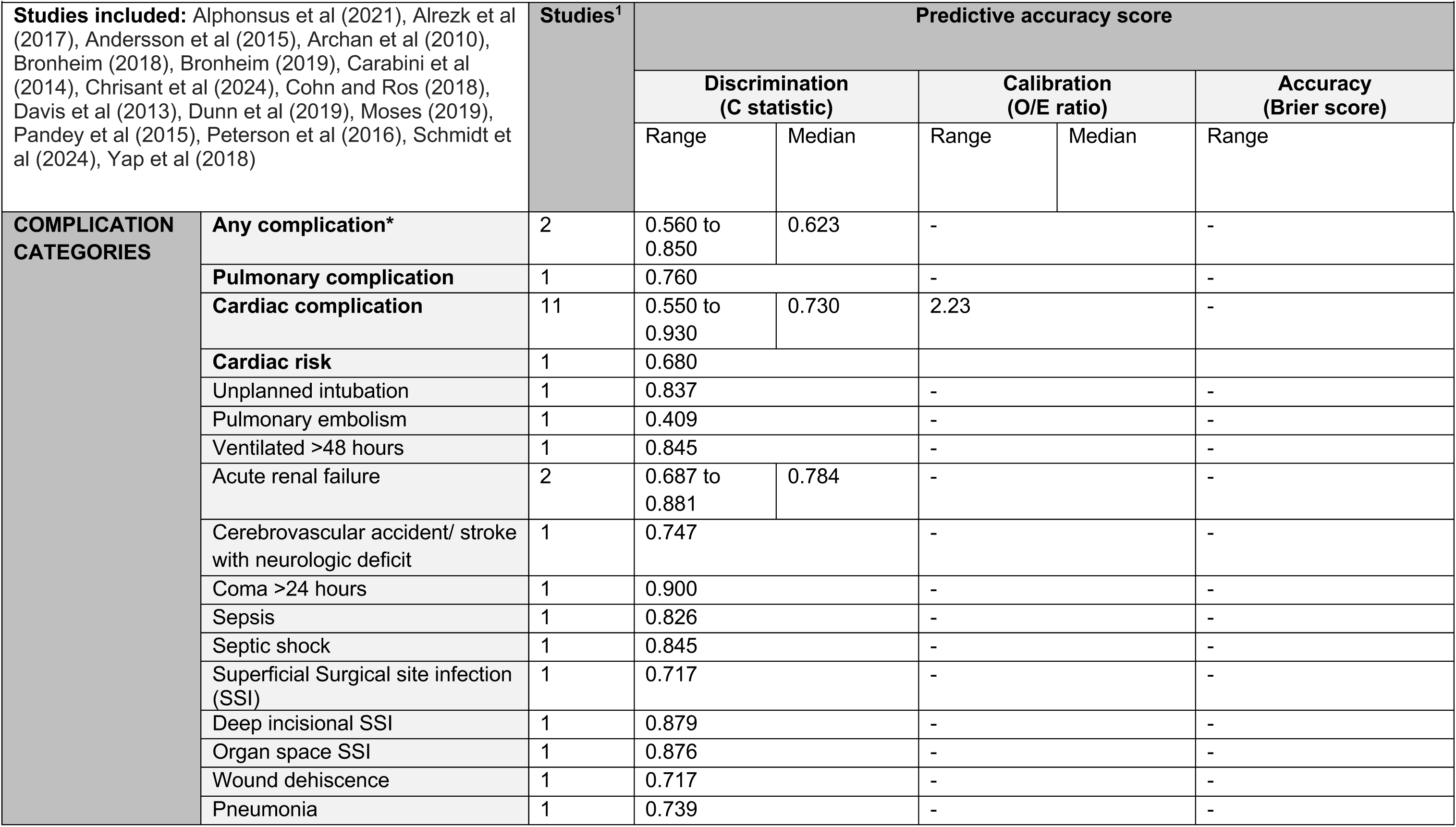
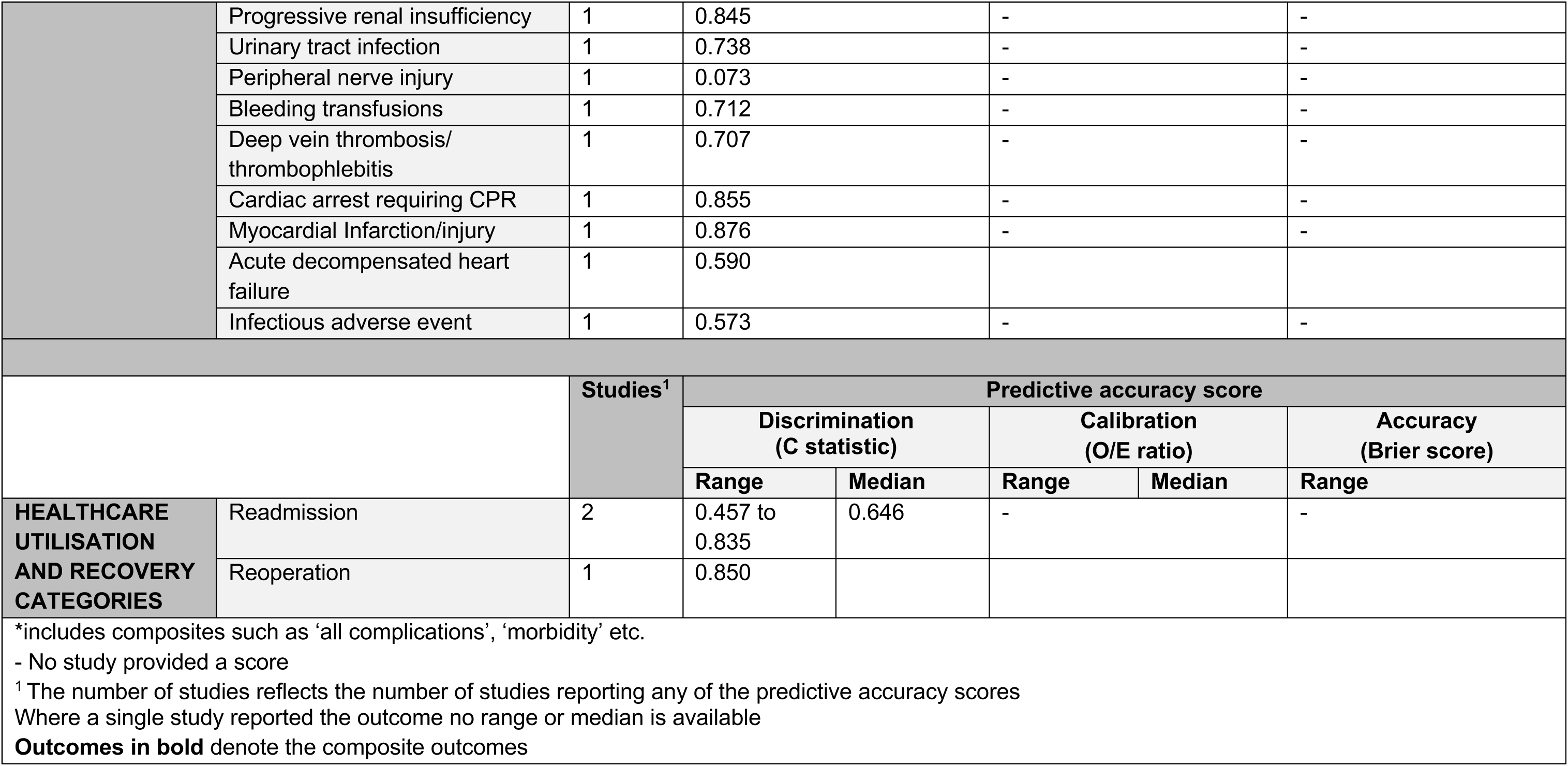
RCRI (16 studies) all studies.

**Table 30.**
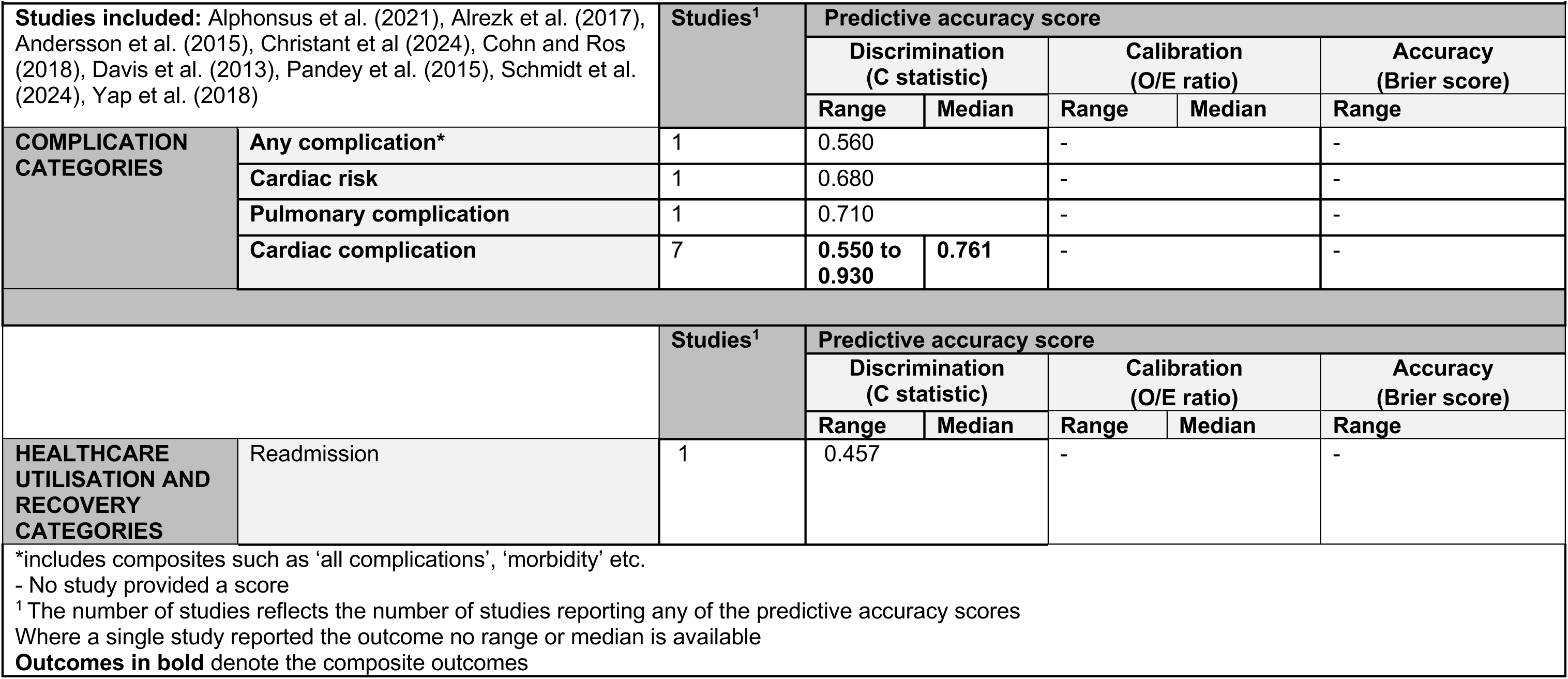
RCRI (9 studies) Mixed Surgery.

**Table 31.** RCRI (4 studies) Orthopaedic Surgery.

| Table 31 – RCRI (4 studies) Orthopaedic Surgery |  |  |  |  |  |  |  |
| --- | --- | --- | --- | --- | --- | --- | --- |
| **Studies included:** Bronheim et al. (2018), Bronheim et al. (2019), Carabini et al. (2014), Peterson et al. (2016) |  | **Studies<sup>1</sup>** | **Predictive accuracy score** |  |  |  |  |
|  |  |  | **Discrimination (C statistic)** |  | **Calibration (O/E ratio)** |  | **Accuracy (Brier score)** |
|  |  |  | **Range** | **Median** | **Range** | **Median** | **Range** |

| COMPLICATION CATEGORIES | Any complication* | 1 | 0.623 |  | - |  | - |
| --- | --- | --- | --- | --- | --- | --- | --- |
|  | Cardiac complication | 2 | 0.540 to 0.900 | 0.72 | - |  | - |
|  |  | Studies<sup>1</sup> | Predictive accuracy score |  |  |  |  |
|  |  |  | Discrimination (C statistic) |  | Calibration (O/E ratio) |  | Accuracy (Brier score) |
|  |  |  | Range | Median | Range | Median | Range |
| HEALTHCARE UTILISATION AND RECOVERY CATEGORIES | Readmission | 1 | 0.835 |  | - |  | - |

\*includes composites such as ‘all complications’, ‘morbidity’ etc.
- No study provided a score
<sup>1</sup> The number of studies reflects the number of studies reporting any of the predictive accuracy scores
Where a single study reported the outcome no range or median is available
**Outcomes in bold** denote the composite outcomes

**Table 32.**
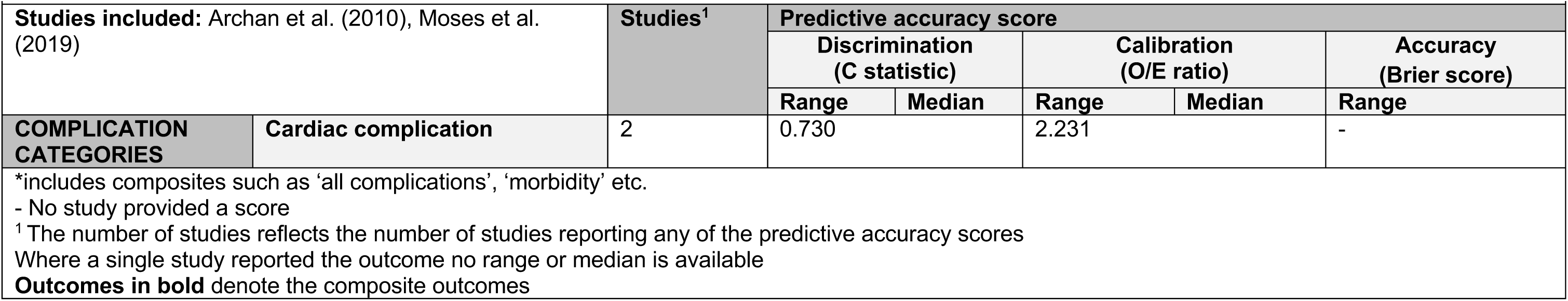
RCRI (2 studies) Vascular surgery.

| Table 32 – RCRI (2 studies) Vascular surgery |  |  |  |  |  |  |  |
| --- | --- | --- | --- | --- | --- | --- | --- |
| Studies included: Archan et al. (2010), Moses et al. (2019) |  | Studies<sup>1</sup> | Predictive accuracy score |  |  |  |  |
|  |  |  | Discrimination (C statistic) |  | Calibration (O/E ratio) |  | Accuracy (Brier score) |
|  |  |  | Range | Median | Range | Median | Range |
| COMPLICATION CATEGORIES | Cardiac complication | 2 | 0.730 |  | 2.231 |  | - |

\*includes composites such as ‘all complications’, ‘morbidity’ etc.
- No study provided a score
<sup>1</sup> The number of studies reflects the number of studies reporting any of the predictive accuracy scores
Where a single study reported the outcome no range or median is available
**Outcomes in bold** denote the composite outcomes

#### ASA classification system

**Table 33.**
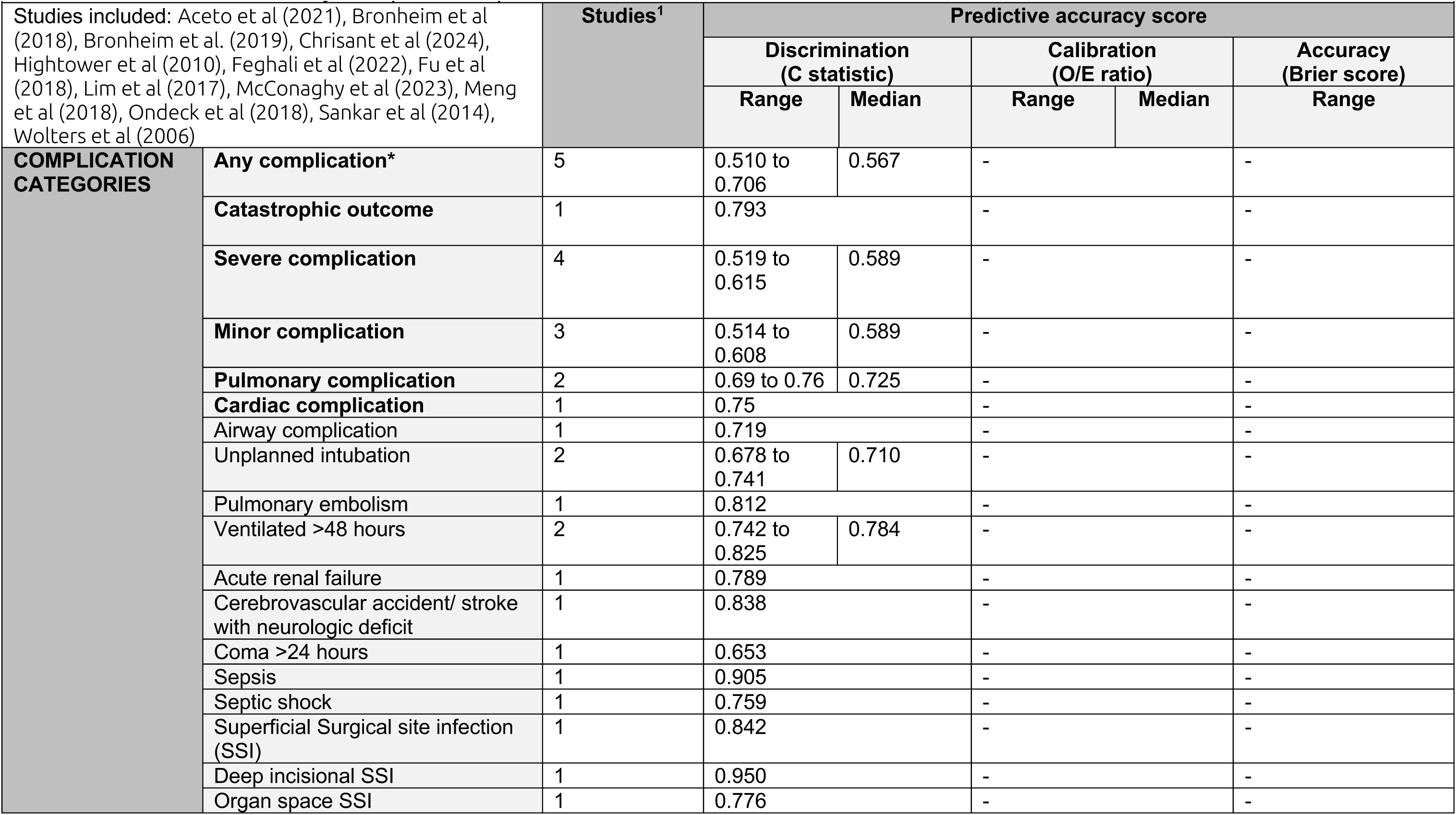
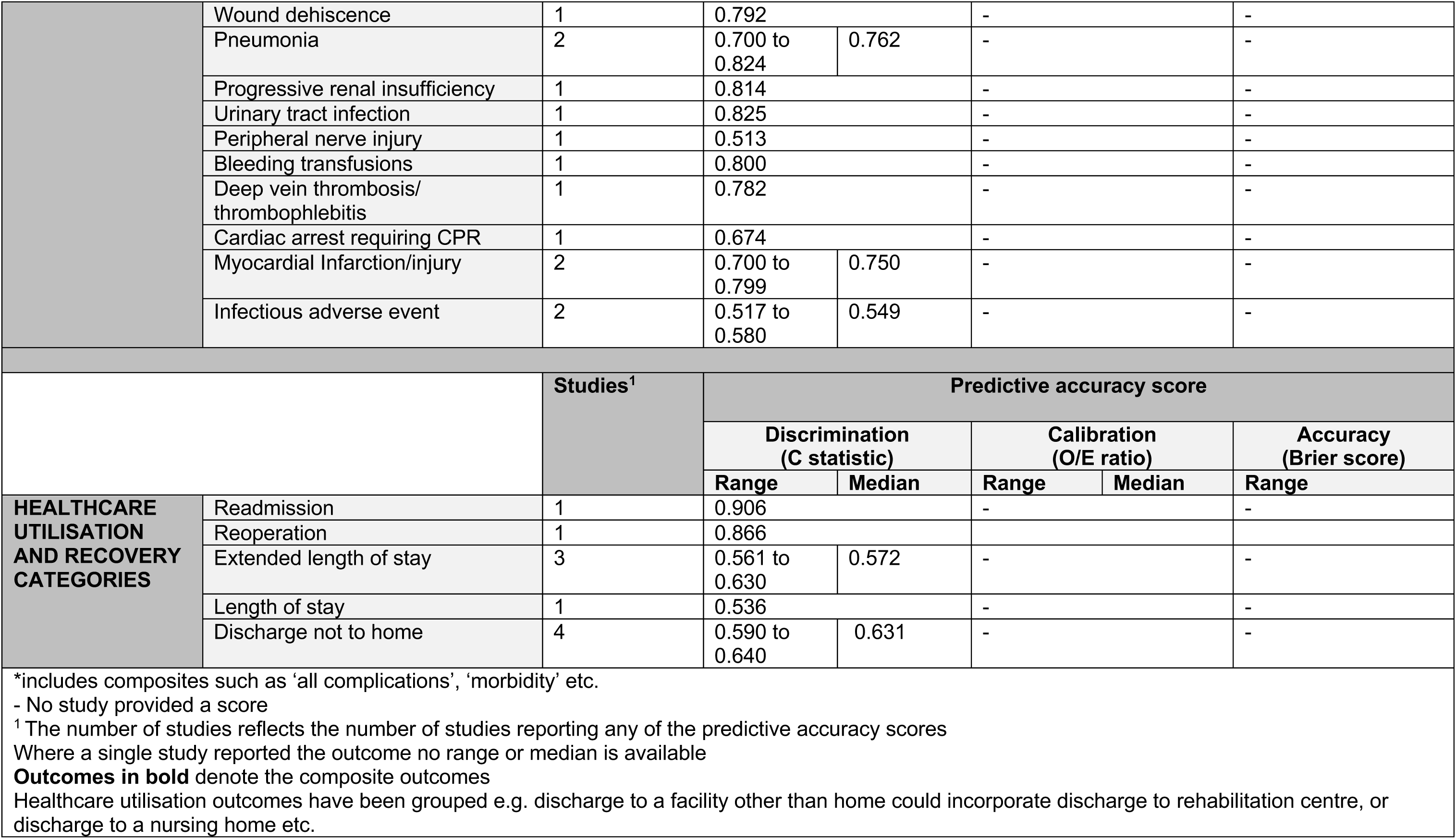
ASA Classification system (13 studies) All studies.

**Table 34.** ASA Classification system (6 studies) Orthopaedic Surgery.

| Table 34 – ASA Classification system (6 studies) Orthopaedic Surgery |  |  |  |  |  |  |  |
| --- | --- | --- | --- | --- | --- | --- | --- |
| Studies included: Bromnheim et al (2018), Bronheim et al (2019), Fu et al (2017), Lim et al (2017), McConaghy et al (2023), Ondeck et al (2018) |  | Studies<sup>1</sup> | Predictive accuracy score |  |  |  |  |
|  |  |  | Discrimination (C statistic) |  | Calibration (O/E ratio) |  | Accuracy (Brier score) |
|  |  |  | Range | Median | Range | Median | Range |
| COMPLICATION CATEGORIES | Any complication* | 3 | 0.567 to 0.706 | 0.607 | - |  | - |
|  | Catastrophic outcome | 1 | 0.793 |  | - |  | - |
|  | Severe complication | 3 | 0.571 to 0.615 | 0.5975 | - |  | - |
|  | Airway complication | 1 | 0.719 |  |  |  |  |
|  | Minor complication | 1 | 0.608 |  | - |  | - |
|  | Pulmonary complication | 1 | 0.760 |  | - |  | - |
|  | Cardiac complication | 1 | 0.750 |  | - |  | - |
|  |  | Studies<sup>1</sup> | Predictive accuracy score |  |  |  |  |
|  |  |  | Discrimination (C statistic) |  | Calibration (O/E ratio) |  | Accuracy (Brier score) |
|  |  |  | Range | Median | Range | Median | Range |
| HEALTHCARE UTILISATION AND RECOVERY CATEGORIES | Readmission | 1 | 0.906 |  | - |  | - |
|  | Reoperation | 1 | 0.866 |  |  |  |  |
|  | Extended length of stay | 3 | 0.561 to 0.630 | 0.572 | - |  | - |
|  | Length of stay | 1 | 0.536 |  | - |  | - |
|  | Discharge not to home | 3 | 0.590 to 0.640 | 0.631 | - |  |  |
| *includes composites such as 'all complications', 'morbidity' etc.<br>- No study provided a score<br><sup>1</sup> The number of studies reflects the number of studies reporting any of the predictive accuracy scores<br>Where a single study reported the outcome no range or median is available<br>**Outcomes in bold** denote the composite outcomes<br>Healthcare utilisation outcomes have been grouped e.g. discharge to a facility other than home could incorporate discharge to rehabilitation centre, or discharge to a nursing home etc. |  |  |  |  |  |  |  |

**Table 35.** ASA Classification system (3 studies) mixed surgery.

| Table 35 – ASA Classification system (3 studies) mixed surgery |  |  |  |  |  |  |
| --- | --- | --- | --- | --- | --- | --- |
| Studies included: Aceto et al (2021), Chrisant et al (2024), Sanker et al (2014) |  | Studies<sup>1</sup> | Predictive accuracy score |  |  |  |
|  |  |  | Discrimination (C statistic) |  | Calibration (O/E ratio) | Accuracy (Brier score) |
|  |  |  | Range | Median |  |  |
| COMPLICATION CATEGORIES | Pulmonary complication | 2 | 0.690 to 0.760 |  | - | - |
|  | Cardiac complication | 1 | 0.750 |  | - | - |
| *includes composites such as 'all complications', 'morbidity' etc.<br>- No study provided a score<br><sup>1</sup> The number of studies reflects the number of studies reporting any of the predictive accuracy scores<br>Where a single study reported the outcome no range or median is available<br>**Outcomes in bold** denote the composite outcomes |  |  |  |  |  |  |

**Table 36.** ASA Classification system (2 studies) Vascular Surgery.

| Table 36 – ASA Classification system (2 studies) Vascular Surgery |  |  |  |  |  |  |
| --- | --- | --- | --- | --- | --- | --- |
| Studies included: Wolters et al (2006), Feghali et al (2022) |  | Studies<sup>1</sup> | Predictive accuracy score |  |  |  |
|  |  |  | Discrimination (C statistic) |  | Calibration (O/E ratio) | Accuracy (Brier score) |
|  |  |  | Range | Median |  |  |
| COMPLICATION CATEGORIES | Any complication* | 1 | 0.518 |  | - | - |
|  | Severe complication | 1 | Range 0.541 to 0.606<br>Median 0.5735 |  | - | - |
| *includes composites such as 'all complications', 'morbidity' etc.<br>- No study provided a score<br><sup>1</sup> The number of studies reflects the number of studies reporting any of the predictive accuracy scores<br>Where a single study reported the outcome no range or median is available<br>**Outcomes in bold** denote the composite outcomes |  |  |  |  |  |  |

### 9.4 Appendix 4. Medline search strategies

- ((NSQIP or “National Surgical Quality Improvement Program”) adj4 (”Surgical Risk Calculator” or “Universal Surgical Risk Calculator”)).ti,ab. 166
- ((POSSUM and ((preop* or “pre-op*” or “postop*” or “post-op*” or risk or predict* or tool* or scor*) and (surg* or repair* or operat*))) or “P-POSSUM” or “Portsmouth-POSSUM” or “Physiological and Operative Severity Score”).ti,ab. 680
- (”Surgical Outcome Risk Tool” or (SORT adj3 ((preop* or “pre-op*” or risk or predict* or tool* or scor*) and surg*))).ti,ab. 137
- or/1-3 972
- (case reports or editorial or guideline or letter or meta analysis or patient education handout or practice guideline or “review” or “systematic review” or comment).pt. 7995825
- 4 not 5 874
- limit 6 to (english language and humans) 686
- exp animals/ not humans.sh. 5288190
- 7 not 8 686
- limit 9 to yr=”2019-Current” 198
- (ARISCAT or (”Postoperative Pulmonary Complications” adj2 scor*)).ti,ab. 69
- (”American Society of Anesthesiologists Classification System” or “ASA Classification System” or “American Society of Anesthesiologists Physical Status Classification System” or “ASA Physical Status Classification System”).ti,ab. 232
- ”Carlisle adj2 Calculator”.ti,ab. or (Carlisle.au,ax. and calculator.ti,ab.) 1
- ((”Clinical Frailty Scale” or CFS or “Clinical Frailty Scor*” or (Rockwood adj2 scor*) or (Rockwood adj2 scale*) or (Rockwood adj1 frailty)) and (preoperat* or “pre-operat*” or ((before or prior or advance or pre or prepar*) adj3 (surg* or operat* or anesthes* or anaesthes* or sedat*)))).ti,ab. 239
- ((”Cardiopulmonary Exercise Test*” or CPET) and (preoperat* or “pre-operat*” or ((before or prior or advance or pre or prepar*) adj3 (surg* or operat* or anesthes* or anaesthes* or sedat*)))).ti,ab. 459
- (DASI or “Duke Activity Status Index”).ti,ab. 406
- (”NELA PRS” or “NELA risk scor*” or “NELA Parsimonious Risk Scor*” or (Parsimonious adj2 “Risk Scor*”) or “National Emergency Laparotomy Audit Parsimonious Risk Score”).ti,ab. 13
- (”Nutrition Risk Screening” or (”NRS-2002” and (preoperat* or “pre-operat*” or ((before or prior or advance or pre or prepar*) adj3 (surg* or operat* or anesthes* or anaesthes* or sedat*))))).ti,ab. 364
- (”Apfel Score for Postoperative Nausea and Vomiting” or ((”Postoperative Nausea and Vomiting” or PONV or Apfel) adj2 (scor* or tool* or scale*))).ti,ab. 361
- ((”Revised Cardiac” adj2 Index) or RCRI).ti,ab. 402
- or/1-10 2519
- (”Children’s Fear Scale” or “Chronic Fatigue Syndrome” or “Corneal Fluorescein Staining” or “Diagnostic Autism Spectrum Interview*” or “Numeric Rating Scale*” or “Palin PRS” or “Personal Response System*” or “Pervasive Refusal Syndrome” or “Pierre Robin Sequence” or “Polygenic Risk Score*” or “R-CODOX-M/R-IVAC” or collision* or “rheumatoid factor cross reactive idiotype”).ti,ab. 69494
- 11 not 12 2460
- (case reports or editorial or guideline or letter or meta analysis or patient education handout or practice guideline or “review” or “systematic review” or comment).pt. 7995825
- 13 not 14 2188
- limit 15 to (english language and humans) 1639

## Abbreviations

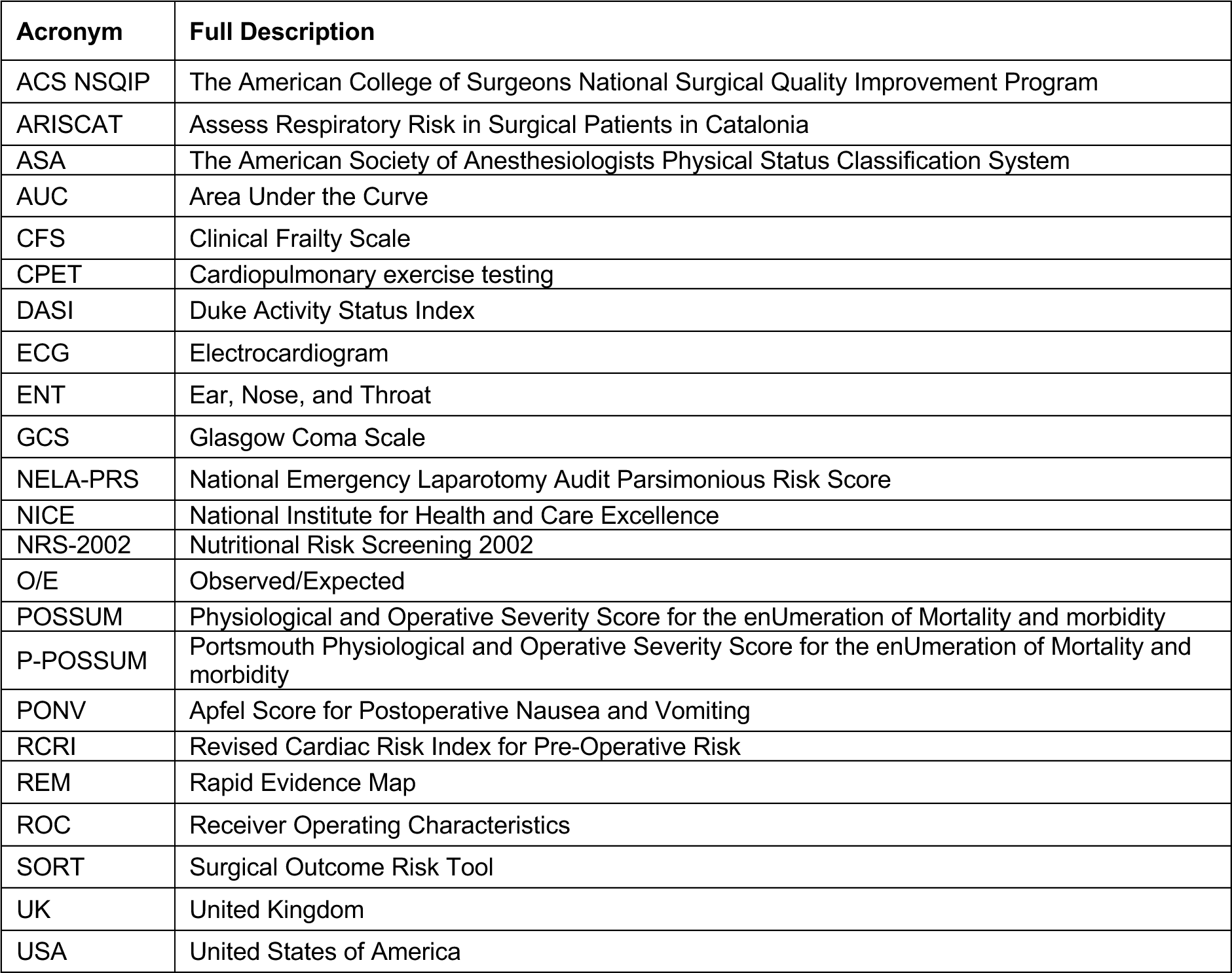

| Acronym | Full Description |
| --- | --- |
| ACS NSQIP | The American College of Surgeons National Surgical Quality Improvement Program |
| ARISCAT | Assess Respiratory Risk in Surgical Patients in Catalonia |
| ASA | The American Society of Anesthesiologists Physical Status Classification System |
| AUC | Area Under the Curve |
| CFS | Clinical Frailty Scale |
| CPET | Cardiopulmonary exercise testing |
| DASI | Duke Activity Status Index |
| ECG | Electrocardiogram |
| ENT | Ear, Nose, and Throat |
| GCS | Glasgow Coma Scale |
| NELA-PRS | National Emergency Laparotomy Audit Parsimonious Risk Score |
| NICE | National Institute for Health and Care Excellence |
| NRS-2002 | Nutritional Risk Screening 2002 |
| O/E | Observed/Expected |
| POSSUM | Physiological and Operative Severity Score for the enUmeration of Mortality and morbidity |
| P-POSSUM | Portsmouth Physiological and Operative Severity Score for the enUmeration of Mortality and morbidity |
| PONV | Apfel Score for Postoperative Nausea and Vomiting |
| RCRI | Revised Cardiac Risk Index for Pre-Operative Risk |
| REM | Rapid Evidence Map |
| ROC | Receiver Operating Characteristics |
| SORT | Surgical Outcome Risk Tool |
| UK | United Kingdom |
| USA | United States of America |

## Glossary

| Term | Definition |
| --- | --- |
| **Accuracy** | Accuracy of risk prediction tools is commonly assessed in the literature using the 'Brier Score'. This is a simultaneous measure of calibration and discrimination, reported as a score between 0 and 1. A score of 0 indicates no difference between the outcome predicted by the tool and actual outcome, thus indicating the best possible result. A score of 1 indicates that the test did not predict the outcome. |
| **Calibration** | Calibration assesses the agreement between the predicted and observed outcomes, typically presented graphically as observed risks versus predicted risks or an observed/expected (O/E) ratio (Collins et al., 2014). An O/E ratio of 1 implies the tool accurately predicted complications, a ratio below 1 suggests the tool overpredicted complications, whereas a ratio over 1 suggests the tool underpredicted complications (Hammond et al., 2021). Following the approach utilised by NICE, an O/E ratio of between 0.9 to 1.1 would be considered a fair level of calibration. |
| **Composite outcomes** | Rather than reporting individual outcomes, some studies combined two or more outcomes into a single measure. For example, 'All complications' could include, pneumonia, sepsis, infection etc. Where outcomes are combined, this is known as a composite outcome. |
| **Discrimination** | Discrimination measures how well a tool differentiates between patients who do and do not experience an event, quantified using the area under the receiver operating characteristic (ROC) curve (AUC) or c-statistic. Within the literature, the following c-statistic scores relate to a tool's performance:<br>• 90% or greater -considered an excellent level of discriminative ability,<br>• 80% or greater is considered a good level of discriminative ability,<br>• 70% or greater is considered a fair level of discriminative ability,<br>• 60% or greater is considered a poor level of discriminative ability,<br>• 50% or greater is considered a very poor level of discriminative ability (or no better than chance) (NICE 2020; Çorbacioğlu and Aksel, 2023). |
| **External validation** | In this context external validation refers to the evaluation of a risk prediction tool's predictive performance in a dataset that was not used to develop the tool, which is known as internal validation (Collins et al., 2014). |
| **Median** | The median is the middle value in a group of numbers ranked in order of size; in this way it is not sensitive to outliers or skewed data. |
| **Mixed surgery** | In this context mixed surgery refers to a study that included a dataset or sample with a range of surgeries across more than one surgical specialty |
| **Rapid Evidence Map** | Rapid Evidence Maps (REMs) use abbreviated systematic mapping or scoping review methods to provide a description of the nature, characteristics and volume of the available evidence for a particular policy domain or research question. The methods have been developed as part of the Health and Care Research Wales Evidence Centre Collaboration. |
| **Risk prediction tools** | Tools, usually a set of clinical or personal information that is used to calculate the estimated likelihood of adverse outcomes, including mortality, morbidity, and postoperative complications. |

